# Obesity Endotypes Unmask Heterogeneous Responses to Healthy Lifestyle Behaviors

**DOI:** 10.64898/2026.08.25.26361367

**Authors:** Devesh Malik, Min Seo Kim, Injeong Shim, Yang Sui, Roukoz Abou-Karam, Minku Song, Hong-Hee Won, Pradeep Natarajan, Patrick T. Ellinor, Akl C. Fahed

## Abstract

**Background:** Lifestyle interventions are central to obesity prevention and management, yet interindividual variability in response remains incompletely understood. Here, we leveraged genetically defined, distinct obesity endotypes to examine lifestyle–body mass index (BMI) associations across biological pathways.

**Methods:** In the UK Biobank, we analyzed 305,713 participants with partitioned polygenic scores (pPSs) representing 10 obesity endotypes. We evaluated interactions between endotype-specific genetic susceptibility and physical activity, diet, sedentary behavior, and sleep on BMI using multivariable linear regression. Primary findings were externally evaluated in the All of Us Research Program using Fitbit-derived lifestyle measures.

**Results:** Favorable lifestyle behaviors were associated with lower BMI for all obesity endotypes, but the magnitude of these associations varied significantly across endotypes. Higher endotype-specific pPSs strengthened the benefits of physical activity (7 endotypes), healthy diet (3 endotypes), nonsedentary behavior (5 endotypes), and adequate sleep (7 endotypes) on BMI. Distinct endotypes demonstrated the greatest responsiveness to different lifestyle domains, with the metabolically unhealthy endotype showing the strongest interaction with physical activity, metabolically healthy endotype with sedentary behavior, hypothalamic dysregulation endotype with diet, and hypoinsulin 2 endotype with sleep, corresponding to differences in BMI of 0.22–0.49 kg/m² between the highest and lowest pPS deciles. These interaction patterns were consistent in the All of Us cohort.

**Conclusions:** Obesity endotypes modify the association between lifestyle behaviors and BMI, demonstrating that responsiveness to lifestyle behaviors is heterogeneous and pathway dependent. These findings provide a framework for precision obesity prevention by identifying individuals who may derive greater benefit from specific lifestyle interventions.

## Introduction

The obesity pandemic represents one of the most pressing public health crises currently. Over the last three decades, its prevalence has more than doubled, with 1 in 8 adults worldwide and 2 in 5 adults in the U.S. now living with obesity.^1,2^ Lifestyle interventions, such as dietary modification and increased physical activity, are well-established as fundamental strategies for obesity prevention and management.^3–5^ However, marked heterogeneity in individual responses to these interventions has been consistently observed, underscoring the limitations of a standardized paradigm and highlighting the need for more biologically informed strategies to optimize disease prevention.^6–8^ Despite this variability, lifestyle interventions are largely applied uniformly, without consideration of the underlying biological heterogeneity of obesity.^9,10^

It is increasingly appreciated that obesity is a collection of biological subtypes or endotypes that converge on excess adiposity but diverge in pathways, risks, and treatment responses.^9–11^ In our recent study, we conducted the largest genome-wide association study (GWAS) for obesity and identified 11 distinct genetic endotypes representing different biological mechanisms driving obesity, each defined with a respective partitioned polygenic score (pPS).^11^ The existence of obesity subtypes has also been independently demonstrated in recent studies by Chami et al. and Coral et al., confirming the reproducibility and biological validity of obesity heterogeneity.^9,10^ Notably, these obesity subtypes and endotypes exhibit distinct associations with obesity-related comorbidities, raising the possibility that underlying biological heterogeneity may account for why certain individuals remain metabolically healthy despite similar lifestyle behaviors, while others progress toward obesity and cardiometabolic disease.^9–11^

Building on this foundation, our prior work introduced partitioned polygenic scores (pPSs) to operationalize obesity heterogeneity by decomposing genetic risk into mechanistically interpretable pathways. Before this pathway-specific framework was available, gene–lifestyle interactions were examined using single, composite polygenic risk scores.^12–16^ Using such a composite score for obesity, we previously showed that the combined effect of high genetic risk of obesity and unfavorable lifestyle was substantially greater than their individual effects, indicating meaningful gene–lifestyle interaction.^14^ However, a single aggregate polygenic score obscures the biological pathways driving obesity risk: two individuals with identical composite obesity polygenic scores may harbor entirely different obesity mechanisms and, consequently, different comorbidity profiles. Endotype-specific pPSs address this limitation by resolving which biological pathways underlie each individual’s genetic risk.^11,17–19^

Despite the advances in partitioned genetic risk scoring, no study has systematically examined how obesogenic lifestyle factors influence body mass index (BMI) across heterogeneous obesity endotypes. The present study addresses this gap by examining endotype-specific genetic risk and its relationship with BMI and lifestyle behaviors. By linking pathway-specific genetic architecture to modifiable behavioral exposures, this work moves beyond population-average models and toward a biologically informed, precision-prevention framework for obesity.

## Methods

### Study population

This study used the UK Biobank, a population-based biobank comprising over 500,000 participants aged 40-69 at recruitment, enrolled between 2006 and 2010 from across the United Kingdom.^20,21^ Participants underwent standardized baseline assessments that included detailed questionnaires on sociodemographic factors, lifestyle behaviors, and medical history, as well as physical measurements, biological sample collection, and genotype data. The UK Biobank was approved by the North West Multi-Centre Research Ethics Committee, and all participants provided written informed consent. This research was conducted using the UK Biobank Resource under application number 7089. All participants with available partitioned polygenic scores for all 10 endotypes and complete lifestyle behavior data were eligible for inclusion in this study. Participants with missing body mass index, key sociodemographic variables, or obesity-related covariate information were excluded.

To assess the generalizability of our primary findings beyond the UK Biobank, we performed external validation in the All of Us Research Program (AoU), a large and demographically diverse U.S. cohort enriched for populations historically underrepresented in biomedical research.^22,23^ The inclusion of AoU data in this study was approved under a data agreement between Massachusetts General Hospital and AoU.^23^ This resource integrates electronic health records with participant-reported data, with all participants providing informed consent under centralized ethical oversight. Phenotypic information is derived from multiple sources, including participant-reported data and linked electronic health records, which capture standardized clinical coding systems such as ICD-9/10, SNOMED, and CPT.^24^ All participants with available partitioned polygenic scores for all 10 endotypes and complete Fitbit data were eligible for inclusion in this study. Participants with missing body mass index, key sociodemographic variables, or obesity-related covariate information were excluded.

### Genotyping and imputation

Genotyping and imputation procedures in the UK Biobank have been described in detail previously.^21^ In brief, participants were genotyped using either the UK BiLEVE Axiom Array or the UK Biobank Axiom Array, each capturing ∼800,000 variants with substantial genome-wide coverage. Genetic imputation was performed utilizing reference panels from the Haplotype Reference Consortium (HRC), UK10K, and the 1000 Genomes Project Phase 3. All imputation procedures were carried out using IMPUTE 4 (https://jmarchini.org/software/). For the present analysis, we excluded samples that did not meet the quality control standards set by the UKB.^21^ From an initial cohort of 488,175 individuals, exclusions were made as follows: 968 individuals for excessive heterozygosity or missing data, 652 for aneuploidy, 378 for sex-gender mismatches, and 510 individuals who withdrew from the study. To ensure a maximally independent set of samples while minimizing the removal of individuals with valuable phenotype data, we employed the ukb_gen_samples_to_remove function from the ukbtools R package, applying a KING kinship coefficient cutoff of 0.0884.^25^

Genetic data methods and quality control within AoU has previously been reported.^23^ Briefly, whole-genome sequencing data were generated for approximately 414,000 participants using Illumina NovaSeq 6000 platforms. Sequencing libraries were prepared using a PCR-free Kapa HyperPrep protocol, and variants were identified using DRAGEN version 3.7.8 across participating genome centers. Quality control procedures were centrally implemented in accordance with the AoU Genomic Quality Report.^26^ Standard variant-level quality control was performed based on the following criteria: (1) monomorphic variants; (2) call rate < 90%; (3) presence in low complexity regions; and (4) population-specific Hardy-Weinberg equilibrium (HWE) P-value < 1 × 10^-15^. At the sample level, individuals flagged by the AoU program for known quality concerns were excluded. We retained only samples with inferred sex ploidy labeled as “XX” or “XY,” removed individuals with genotype missingness exceeding 5%, and addressed duplicate samples. Duplicate identification was performed using KING (v2.3.2), where pairs with heterozygous concordance greater than 0.8 were considered potential duplicates.^27^ From each such pair, one individual was removed, prioritizing retention of participants with linked electronic health record (EHR) data, followed by higher genotype call rate, with EHR availability given greater weight. After applying these filters, 18,752,405 autosomal variants and 410,400 samples remained. To characterize population structure, we conducted principal component analysis (PCA), rather than relying on AoU-provided components derived from external reference panels (HGDP and 1000 Genomes Project).^28^ A subset of autosomal variants was first curated for both relatedness estimation and PCA, requiring minor allele frequency > 0.01 and genotype missingness < 1%. Linkage disequilibrium pruning was performed using PLINK with parameters --indep-pairwise 500 200 0.1 and --indep-pairwise 2000 400 0.1, and variants within long-range LD regions were excluded.^29,30^ From the remaining pool, 100,000 variants were randomly selected. Using this pruned set, pairwise relatedness was estimated with KING, and unrelated individuals were defined by a kinship coefficient < 0.042.^27^ PCA was then performed with flashPCA (v2.0), using only unrelated individuals.^31^ The resulting principal components were subsequently projected onto the full dataset. Final analyses were restricted to individuals who were both unrelated and had linked EHR and genetic data.

### Endotype-specific partitioned polygenic scores

Our previous work demonstrated significant interactions between the obesity polygenic score and obesogenic lifestyle factors in the risk of obesity and obesity-related morbidities.^14^ Endotype-specific partitioned polygenic scores were derived from our previous work.^11^ These endotypes, also known as genetic clusters, were identified using a multi-trait, multi-ancestry genome-wide association study (GWAS) integrating anthropometric measures and downstream genetic clustering using Bayesian non-negative matrix factorization. This approach grouped significant obesity-associated variants into biologically coherent clusters, each representing a distinct genetic pathway contributing to obesity risk. Each obesity endotype was defined by discrete phenotypic, tissue-level, and single-cell regulatory features, and the resulting endotype-specific pPSs were validated in an independent cohort, the Mass General Brigham (MGB) Biobank. In the present study, these pPSs were used as measures of genetic predisposition to each obesity endotype, rather than direct measurements of the corresponding biological endophenotypes. Because the GWAS datasets used for clustering were derived from multiple large cohorts, some may have included participants from the UK Biobank. Consequently, the cohort used in the present study may not be fully independent from all GWAS datasets contributing to pPS construction. However, the obesity GWAS used for the primary score derivation excluded UK Biobank participants, and many of the traits used for clustering reflect cardiometabolic rather than lifestyle-related phenotypes. Due to instability from its reliance on a single contributing variant, the Hypo-LpA endotype was excluded. The remaining 10 endotypes, each driven by and named after its predominant biological feature, were as follows: Metabolically unhealthy, Strong beta cell, Hypoinsulin 1, Hypoinsulin 2, Proinsulin, Immune dysregulation, Hyperinsulin 1, Hyperinsulin 2, Hypothalamic dysregulation, and Metabolically healthy.

In the UK Biobank, 10 raw pPSs were calculated for each participant by summing the effect allele dosages across all variants assigned to a given endotype, weighted by their corresponding endotype-specific β coefficients. All PRS calculations were performed using the PLINK2 software via the --score function.^29^

We adjusted all polygenic scores for enrollment age, sex, genotyping array, and the first four principal components of genetic ancestry. Subsequently, all scores were residualized for the first four principal components of genetic ancestry and then scaled to a mean of 0 and a standard deviation of 1.

Scores were computed independently for each endotype using individual-level genetic data. To account for ancestry-specific effects, we regressed the top 10 principal components (PCs) from the raw pPS and standardized the residuals to a normal distribution.

### Harmonization of partitioned polygenic scores

To enable direct comparison of effect sizes across endotypes in the primary analysis, standardized pPSs were further harmonized to a similar association with BMI. Specifically, for each endotype, a multivariable linear regression model was fit with BMI as the outcome and the standardized pPS as the primary exposure, adjusting for relevant covariates. The median of the resulting adjusted regression β coefficients for the 10 endotypes was selected as a fixed reference value for harmonization. Each standardized pPS was subsequently rescaled by the ratio of its endotype-specific BMI coefficient to this reference value, as shown below:

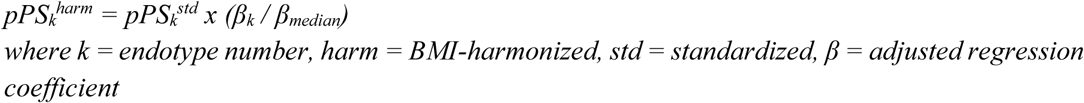

This harmonization process yielded pPSs with aligned effect estimates on BMI, altering score variance while preserving the underlying distributions and participant rank orderings.

### Obesity endotype membership and cumulative burden

For secondary analyses, individuals were classified as belonging to a certain endotype (membership to endotype) if their pPS value fell in the top decile of the corresponding pPS (approximately equivalent to a z-score of 1.28), a threshold commonly used in polygenic risk score studies to define individuals with high genetic susceptibility.^32,33^ Of note, obesity endotypes are not mutually exclusive; an individual may exhibit multiple concurrent endotypes, reflecting the coexistence of more than one biological pathway to obesity. A cumulative obesity endotype burden score was constructed by summing the number of endotypes for which each participant fell within the top decile.

### Lifestyle factors

Drawing from baseline self-reported questionnaires in the UK Biobank obtained at initial enrollment, we derived four well-established obesogenic lifestyle factors previously described in the literature: physical activity, diet, sedentary behavior, and sleep.^15,34^ Each was operationalized as a binary indicator reflecting favorable versus unfavorable behaviors, following the approach used in our prior gene–lifestyle interaction study.^6^ A healthy dietary pattern was defined using a composite score based on eight dietary components, with participants classified as having a healthy diet if they met at least four of the following criteria: higher intake of fruits, vegetables, whole grains, and fish, along with lower intake of refined grains, processed meats, unprocessed red meats, and sugar-sweetened foods or beverages. Physical activity was considered favorable if total activity exceeded 3,000 metabolic equivalents of task (MET) minutes per week, corresponding to approximately the highest 30% of activity levels in the UK Biobank population. Lower sedentary behavior was defined as spending no more than 2 hours per day watching television or using a computer outside of work-related activities. Adequate sleep duration was defined as an average of 6 to 8 hours per day (Supplementary Table 1).

### Primary and Secondary Outcomes

The primary outcome was body mass index (BMI), a clinically actionable measure of adiposity that predicts cardiometabolic morbidity, is routinely assessed in clinical practice, and reflects the integrated contributions of genetic and environmental factors.^14,35–42^ BMI was measured at initial evaluation in UK Biobank participants, calculated as weight in kilograms divided by height in meters squared according to the protocol of the UK Biobank.

Secondary outcomes included waist circumference (WC), hip circumference (HC), waist-to-hip ratio (WHR), and waist-to-height ratio (WHtR). These anthropometric measures are well-established correlates of obesity, cardiometabolic disease, and all-cause mortality and therefore serve as robust indicators of overall metabolic health.^43–53^ WC, HC, and height were measured directly at the initial evaluation, whereas WHR and WHtR were calculated during analysis by dividing WC by either HC or height, respectively.

### Covariates

All regression models were adjusted for a predefined set of demographic, socioeconomic, behavioral, and clinical covariates, selected *a priori* based on established associations with obesity. Demographic covariates included age at baseline and sex. Socioeconomic status was captured using educational attainment (college/university degree or professional qualifications = 1; other = 0) and Townsend Deprivation Index (categorized into 3 groups: bottom 20% [least deprived], middle 60% [intermediate], and top 20% [most deprived]). Behavioral covariates included smoking status (never, former, current) and the four binarized obesogenic lifestyle factors evaluated in this study (physical activity, diet, sedentary behavior, and sleep). Clinical covariates comprised self-reported or clinically ascertained diagnoses of type 2 diabetes mellitus, depression, hypothyroidism, Cushing’s syndrome, and polycystic ovary syndrome, as well as the use of weight-gaining medications, all of which are established risk factors for obesity (Supplementary Tables 2 and 3). To account for population structure, all models additionally included the first ten genetic principal components of ancestry.

### Statistical analyses

Multivariable linear regression models were used to evaluate associations between endotype-specific pPSs and BMI, lifestyle behaviors and BMI, and cumulative obesity endotype burden and BMI. Multiplicative interaction effects between genetic endotypes and lifestyle behaviors on BMI were assessed via multivariable linear regression models with interaction terms, using (i) continuous pPSs with binarized lifestyle variables and (ii) cumulative obesity endotype burden with binarized lifestyle variables; this analytical approach has been applied previously in gene-by-environment interaction studies.^6,54–56^ Each multivariable model included a single endotype-specific pPS (and the single corresponding interaction term in interaction analyses) unless otherwise specified, consistent with prior work leveraging partitioned polygenic scores.^32,33^ The linearity assumption between endotype-specific pPSs (exposure) and BMI (outcome) was assessed using restricted cubic splines.^56–58^ Predicted BMI across endotype percentiles was estimated from the interaction models by generating standardized predictions under favorable and unfavorable lifestyle conditions and averaging predictions over the observed covariate distribution.

Correlation among the continuous pPSs was performed using Pearson correlation; correlation coefficients are considered poor if |r| < 0.30.^59^ Correlation among lifestyle behaviors was assessed using the φ (phi) coefficient for binary variables; correlations were considered weak if |φ| < 0.30.^60^ Correlation between the continuous pPSs and binary lifestyle variables was assessed using the point-biserial correlation (r_pb_) coefficient; correlations were considered weak if |r_pb_| < 0.30.^61,62^ Analyses comparing BMI estimates across genetic strata were performed using estimated marginal means (a.k.a least-squares means) from multivariable models with interaction terms.^63^ Specifically, estimated marginal means were used for (i) evaluating lifestyle–BMI associations across levels of cumulative obesity endotype burden, and (ii) comparing lifestyle–BMI associations between top and bottom decile strata of each endotype. Significance was determined at a p-value of less than 0.05, utilizing a two-tailed test. Multiple testing was corrected using the Benjamini-Hochberg false detection rate (BH-FDR) method. All statistical analyses were performed using R (Version 4.5.1).

## Results

### Study participants

This study included 305,713 participants from the UK Biobank with endotype-specific pPSs, lifestyle data, and covariate information (Supplementary Fig. 1). The mean age (SD) was 56.8 (8.0) years, 52.3% were women, and the mean BMI (SD) was 27.4 (4.7) kg/m^2^ (Supplementary Table 4). Favorable lifestyle behaviors exhibited wide variation, with 11.1% of participants nonsedentary, 31.0% physically active, 35.8% adhering to a healthy diet, and 86.9% reporting adequate sleep. Women demonstrated a higher prevalence of favorable sedentary behavior and dietary patterns compared to men. When lifestyle factors were aggregated, 46.6% of participants adhered to ≤1 favorable lifestyle behavior, whereas only 15.6% demonstrated ≥3 favorable behaviors, highlighting marked variability in health behavior profiles across the cohort. Cumulative obesity endotype burden also showed a broad distribution: 33.8% of participants had exactly 1 composite endotype top decile, 17.3% had exactly 2 top deciles, and 9.4% had ≥3 top deciles, with similar proportions in men and women. Finally, 39.8% received higher education and 18.8% resided in the lowest socioeconomic quintile.

### Obesity endotypes

We first characterized the distribution and overlap of obesity endotypes within the cohort. All 10 endotype-specific pPSs, both standardized and BMI-harmonized, demonstrated a normal distribution centered at zero, with symmetrical tails and no evidence of outlier inflation (Supplementary Fig. 2). Correlations between endotype-specific pPSs were generally weak or absent (|r| < 0.30), indicating minimal overlap between genetic endotype scores (Supplementary Fig. 3). When participants with at least one endotype-specific top decile membership were grouped by unique endotype membership, the majority of individuals belonged to only one endotype (Figure 1a/b). Combinations involving two or more endotypes occurred progressively less frequently. Mean BMI values were calculated for participants in the top decile of each endotype-specific pPS, irrespective of membership to other endotypes (Supplementary Fig. 4). Across the ten endotypes evaluated, mean BMI values showed some variability between endotype groups, but the range of differences was relatively small (27.55 to 28.55 kg/m^2^). The lowest mean BMI was observed in the Proinsulin endotype, and the highest mean BMI was in the Metabolically unhealthy endotype.

**Figure 1.**
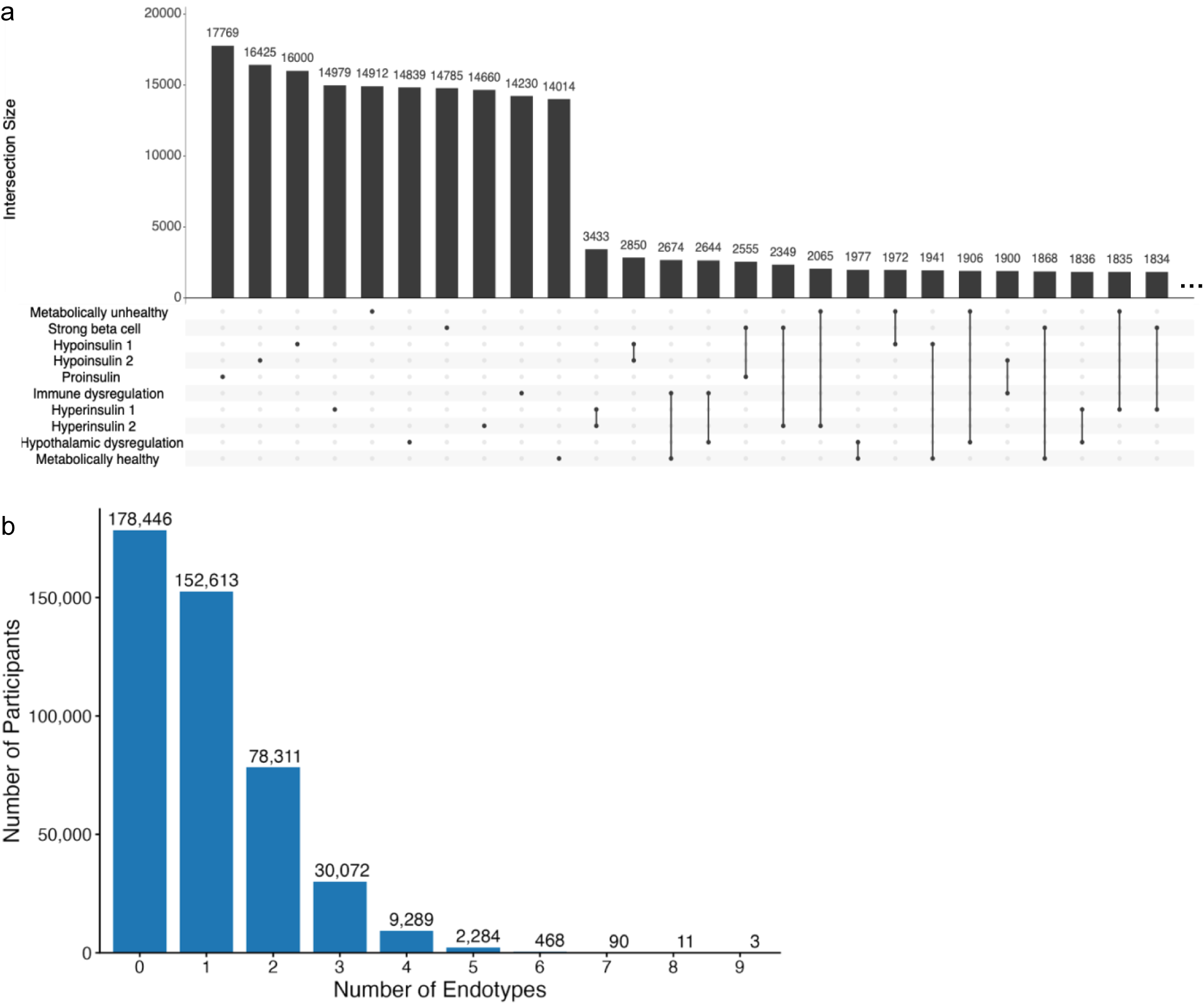
Combinatorial classification and cumulative distribution of obesity endotypes. (a) UpSet Plot showing all observed combinations of top decile status across the 10 obesity endotypes. Interaction size denotes the number of participants exhibiting each combination in the intersection matrix. The set size indicates how many participants are in the top decile for each endotype, regardless of membership to other endotype top deciles. All set sizes are equivalent as they are defined as 10% of the total cohort. “…” indicates truncation of less frequent combinations not shown. Of the 1,024 possible top decile combinations (including membership to zero endotype top deciles), our cohort has 878 combinations. (b) Distribution of participants by the number of obesity endotypes for which they are classified in the top decile.

### Relationship between obesity endotypes and BMI

Having characterized the distribution and population structure of endotype-specific pPSs, we next examined how these scores related to BMI. In multivariable models adjusting for demographic and clinical covariates, each standardized pPS was positively associated with BMI (Figure 2a). The estimated increase in BMI per 1-SD increment in pPS differed by endotype, indicating that endotypes contribute variably to BMI. To enable direct comparison of effect modification across endotypes on a common scale for future analyses, we harmonized pPSs to a shared BMI effect. After harmonization, the estimated associations aligned to a shared effect size for all endotypes as expected (Supplementary Fig. 5).

**Figure 2.**
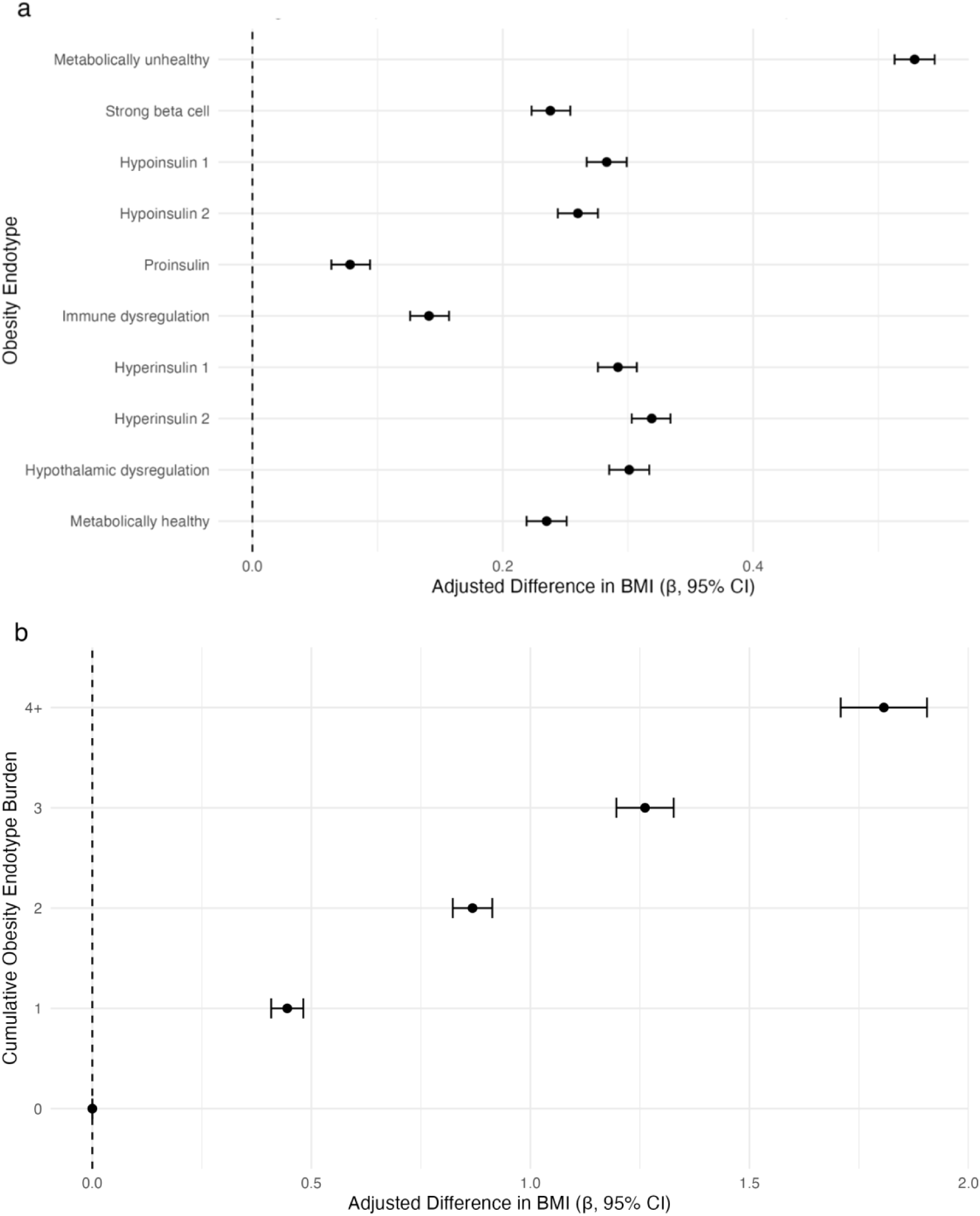
Associations of individual obesity endotypes and cumulative endotype burden with BMI. (a) Forest plot showing the change in BMI per 1-standard-deviation increase in each standardized partitioned polygenic score (pPS). Effect estimates were derived from separate multivariable linear regression models, each with the respective pPS as the exposure, BMI as the outcome, and adjustment for covariates. (b) Forest plot showing the difference in BMI associated with increasing cumulative obesity endotype burden, defined as the total number of endotypes for which an individual is classified in the top decile. Effect estimates were derived from a multivariable linear regression model with cumulative endotype burden as the exposure, BMI as the outcome, and adjustment for covariates. CI: confidence interval.

Cumulative obesity endotype burden, defined as the number of endotypes for which a participant was in the top decile, was associated with graded increases in BMI (Figure 2b). Relative to individuals with no top decile classifications, those with one, two, three, and ≥4 endotypes in the top decile exhibited progressively larger BMI differences, consistent with a dose-dependent pattern of multi-endotype genetic susceptibility.

### Baseline associations of lifestyle behaviors and endotypes with BMI

To contextualize potential effect modification of obesity endotypes on lifestyle’s impact on BMI in subsequent analyses, we first quantified the independent associations of each lifestyle factor with BMI. In adjusted models, favorable sedentary behavior, physical activity, diet, and sleep were each associated with lower BMI, with varying effect estimates across the behaviors; these findings have been shown previously in the literature (Supplementary Fig. 6).^6,34^ Pairwise correlations between lifestyle behaviors were minimal (|φ| ≤ 0.07), indicating that the four lifestyle variables were largely independent within the cohort (Supplementary Fig. 7). Point-biserial correlations between lifestyle factors and obesity endotype scores were negligible (|r_pb_| ≤ 0.02), indicating minimal association between lifestyle behaviors and genetic endotype scores (Supplementary Fig. 8).The linear relationship assumption between continuous pPSs and BMI for multivariable linear regression was satisfied using restricted cubic spline models which allowed for flexibility in the functional form (Supplementary Fig. 9). Across all ten endotypes, predicted BMI increased approximately linearly with increasing pPS.

### Endotype-specific modification of lifestyle–BMI associations

Having established the significant associations between endotype-specific pPSs and BMI, as well as between lifestyle behaviors and BMI, we next evaluated whether the obesity endotypes modified the association between lifestyle behaviors and BMI. Endotype–lifestyle interaction models showed that the association between favorable lifestyle behaviors and BMI differed according to BMI-harmonized continuous pPS (Figure 3; Supplementary Tables 5-44). Significant negative interactions were observed for several endotype–lifestyle combinations, indicating that individuals with higher endotype scores benefit more from a healthy lifestyle in terms of BMI reduction (i.e. greater reduction in BMI from favorable lifestyle compared to those with average endotype score). Additionally, interaction magnitudes varied across endotypes; for example, among individuals with Strong beta cell scores 1 standard deviation above the mean, the interaction with favorable physical activity was approximately twofold stronger than that observed for Metabolically unhealthy, despite both showing significant interactions with physical activity. Linear interaction plots that show predicted BMI trajectories stratified by lifestyle status further illustrate these differences (Supplementary Fig. 10, 11, 12, and 13).

**Figure 3.**
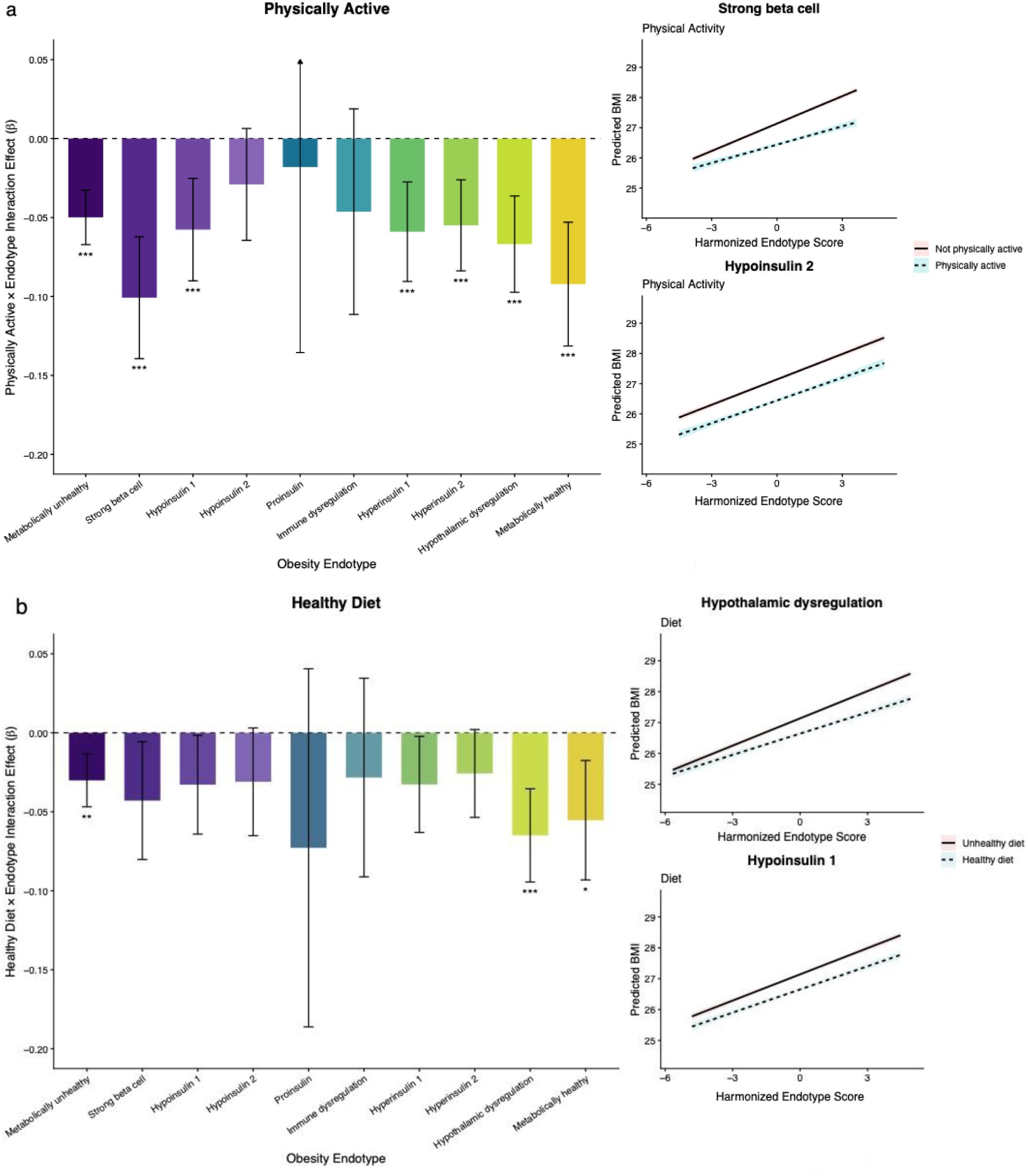
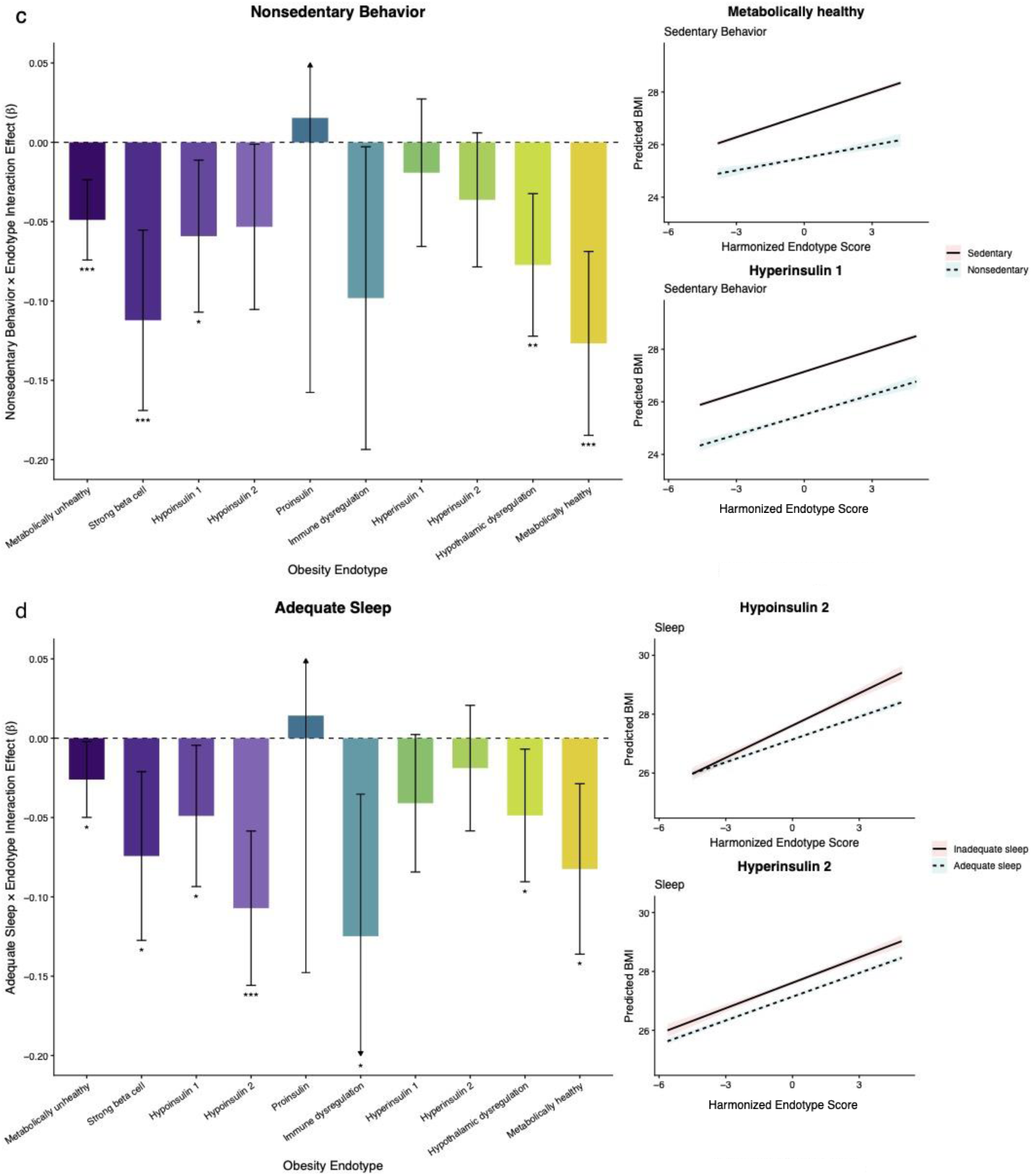
Interaction effects between favorable lifestyle behaviors and obesity endotypes on BMI. Bar plots showing lifestyle–endotype interaction effect estimates from multivariable linear regression models evaluating whether associations between lifestyle behaviors and BMI differ by BMI-harmonized obesity endotype scores. Since favorable lifestyle behaviors are associated with lower BMI (negative main-effect estimates), a significant negative lifestyle–endotype interaction estimate indicates that higher endotype scores are associated with a greater BMI reduction from the favorable lifestyle option compared to lower scores. As harmonization was performed to a pre-selected BMI effect, the absolute values are inconclusive; however, conclusions regarding relative effects are appropriate. Separate models were fitted for each combination of partitioned polygenic score (continuous) and lifestyle behavior (binarized). Error bars represent the 95% confidence interval. Linear interaction plots show predicted BMI across the range of BMI-harmonized endotype scores, stratified by lifestyle favorability, illustrating that for endotypes with significant interaction effects, the lines for favorable and unfavorable lifestyle groups diverge as endotype score increases, whereas for non-significant interactions, the lines remain largely parallel, indicating minimal differential effect. The examples shown represent one significant and one non- significant endotype for each lifestyle behavior, selected for illustration from the corresponding bar plots. Shaded bands represent the 95% confidence interval. Panels correspond to (a) physically active, (b) healthy diet, (c) nonsedentary behavior, and (d) adequate sleep. BH-FDR correction was applied to all p- values within each lifestyle-specific set of models. * = p-value < 0.05; ** = p-value < 0.01; *** = p-value < 0.001.

To further characterize the nature of these interactions across the full distribution of genetic risk, we examined predicted BMI differences between favorable and unfavorable lifestyle behaviors across percentiles of each endotype-specific polygenic score (Figure 4; Supplementary Tables 5-44; Supplemental Fig. 14-18). Across all lifestyle domains, the magnitude of BMI reduction associated with favorable behaviors increased progressively with higher endotype-specific genetic risk, consistent with effect modification observed in the interaction models. However, both the magnitude and shape of these gradients varied across endotypes, indicating heterogeneity in how genetic pathways modulate lifestyle responsiveness. Notably, the endotypes demonstrating the strongest gradients differed by lifestyle domain. For example, the metabolically unhealthy endotype showed the most pronounced gradient for physical activity, hypothalamic dysregulation for diet, hypoinsulin 2 for sleep, and metabolically healthy for sedentary behavior, highlighting biologically distinct patterns of lifestyle responsiveness across endotypes.

**Figure 4.**
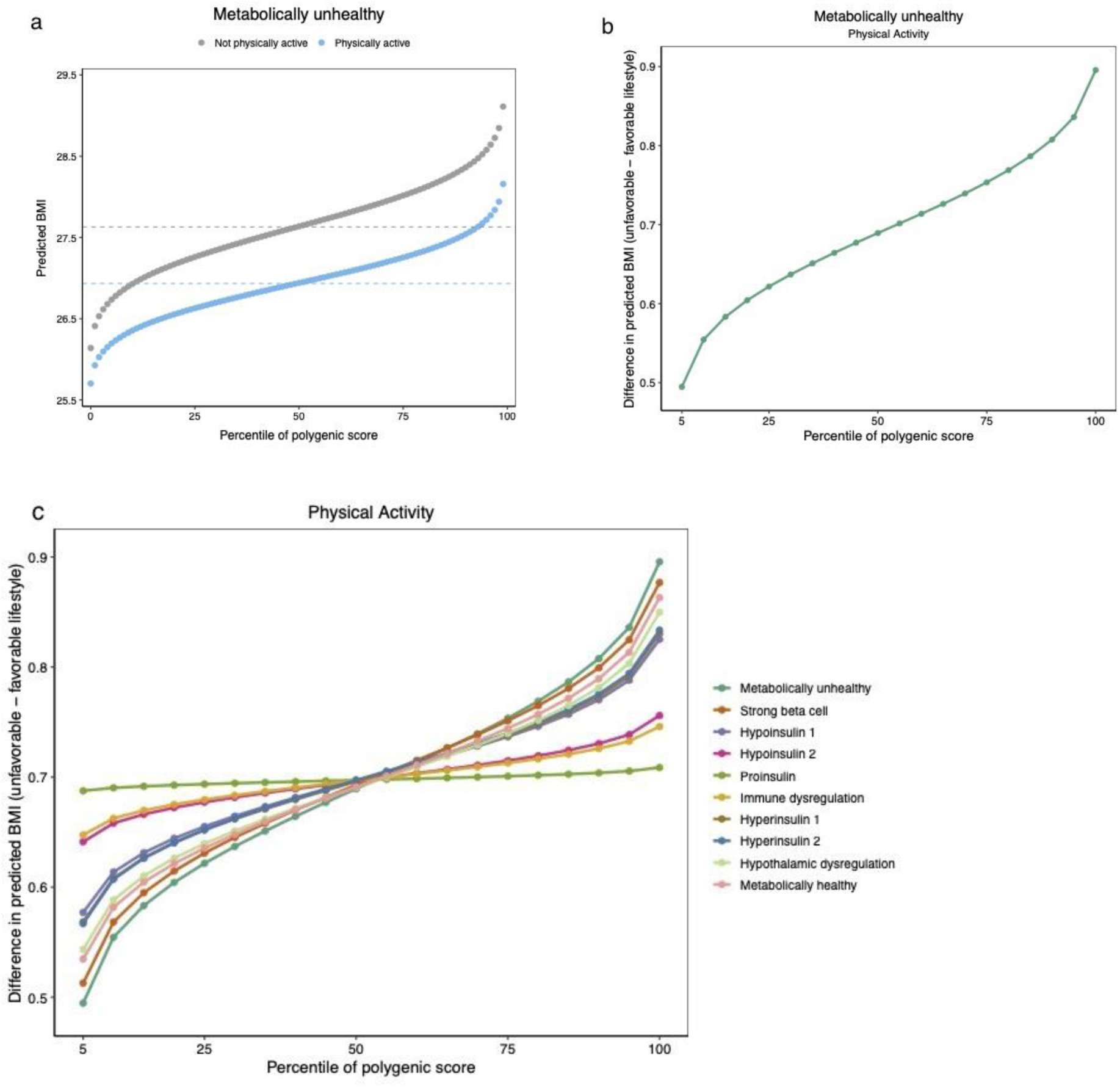
Difference in predicted BMI between unfavorable and favorable lifestyle across genetic risk percentiles by obesity endotype. Conditional and contrast plots illustrating the estimated difference in predicted BMI (unfavorable minus favorable lifestyle) across percentiles of genetic risk. (a) Conditional plot showing the predicted BMI for unfavorable and favorable physical activity at every percentile of the Metabolically unhealthy endotype score. (b) Contrast plot showing the difference between predicted BMI values for unfavorable and favorable physical activity at every vigintile of the Metabolically unhealthy endotype score. (c) Contrast plot showing the difference between predicted BMI values for unfavorable and favorable physical activity at every vigintile of all 10 endotype scores. Positive values indicate a greater BMI reduction associated with favorable lifestyle behaviors compared to unfavorable behaviors. All estimates were derived from multivariable linear regression models including an interaction term between harmonized polygenic score (continuous) and lifestyle behavior (binarized), adjusted for covariates.

To contextualize these continuous gradients, we next examined estimated BMI differences between favorable and unfavorable lifestyle behaviors at the extremes of genetic risk, comparing individuals in the highest versus lowest deciles of each obesity endotype (Supplementary Fig. 19). Consistent with the patterns observed across the full distribution in Figure 4, individuals in the top decile of endotype-specific genetic risk generally exhibited greater BMI reductions associated with favorable behaviors than those in the bottom decile, varying across endotypes and across lifestyle domains. For example, individuals in the highest decile of the Metabolically unhealthy endotype experienced an additional 0.41 kg/m² reduction in BMI with favorable physical activity compared to those in the lowest decile (−0.97 vs −0.56 kg/m²). These differences highlight that the magnitude of behavioral benefit can vary meaningfully at the extremes of genetic susceptibility, providing an interpretable scale for the potential clinical impact of gene–lifestyle interaction.

### Lifestyle–BMI associations across cumulative obesity endotype burden

We next examined effect modification across cumulative genetic susceptibility. Across cumulative obesity endotype burden, favorable lifestyle behaviors were consistently associated with lower BMI, with the magnitude of these associations increased by burden level (Supplementary Fig. 20). For physical activity, the estimated BMI difference comparing favorable versus unfavorable behavior was greatest among individuals with three or more obesity endotypes (β = -1.00, [95% CI: -1.12, −0.90]), and progressively smaller at lower burden levels (β = −0.72 [-0.80, −0.64] , −0.69 [-0.74, −0.63], and −0.61 [-0.67, −0.56] for burden counts of 2, 1, and 0, respectively). Diet and sleep showed similar patterns, with larger BMI differences between favorable and unfavorable behaviors among those with higher endotype burden (diet: β = −0.69 [-0.80, −0.58] for burden ≥3 vs β = −0.42 [-0.47, −0.37] for burden 0; sleep: β = −0.71 [-0.86, −0.56] for burden ≥3 vs β = −0.37 [-0.45, −0.30] for burden 0). The strongest associations were observed for sedentary behavior, for which favorable status corresponded to β estimates ranging from –1.89 [-2.06, - 1.73] (burden ≥3) to –1.50 [-1.58, -1.43] (burden 0). Although confidence intervals overlapped across strata for several behaviors, the pattern of increasingly negative estimates with higher burden suggests that favorable lifestyle may confer greater BMI reduction among individuals harboring multifaceted pathways to obesity risk, consistent with effect modification.

### Sensitivity analyses and external replication

Several sensitivity analyses were conducted to assess the robustness of the primary findings. First, BMI- harmonized endotype–lifestyle interactions were estimated using continuous measures of lifestyle behaviors where available (physical activity defined as met-minutes per week, and sedentary behavior defined as -[time spent in front of screen]); the resulting interaction estimates were consistent with those observed using binarized lifestyle definitions (Supplementary Fig. 21). Second, using longitudinal Fitbit data available for a subset of participants in AoU (n = 25,661), we replicated our primary interaction findings across key lifestyle domains, including physical activity (defined as favorable if moderate-to-vigorous activity, calculated as fairly active minutes + 2 × very active minutes, was in the top 30% of the All of Us distribution), sedentary behavior (modeled as –[continuous sedentary minutes]), and sleep duration (defined as an average of 6–8 hours per day). Endotype–lifestyle interaction patterns were directionally consistent and preserved the relative pattern across endotypes observed in the UK Biobank (Supplementary Fig. 22). Due to reduced sample size and limited power to detect interaction effects, replication was evaluated based on concordance in effect direction and relative magnitude across endotypes rather than statistical significance alone.^64^

## Discussion

In this large, population-based analysis of more than 300,000 individuals, we demonstrate that obesity endotypes variably modify the magnitude of BMI reduction associated with favorable lifestyle behaviors. While prior studies have leveraged genome-wide polygenic risk scores to quantify overall genetic susceptibility to obesity, such scores aggregate risk across diverse biological processes into a single summary measure.^12–14^ Consequently, individuals with comparable global genetic risk may develop obesity through distinct underlying mechanisms. Endotype-specific partitioned polygenic scores, in contrast, retain pathway-level resolution, enabling genetic susceptibility to be interpreted in mechanistic rather than purely quantitative terms.^11,17,19,32,65^ This distinction is particularly relevant for obesity, a condition driven by diverse biological processes with variable downstream health implications. Within this framework, our findings indicate that the impact of lifestyle behaviors on BMI depends on the dominant genetic pathway underlying obesity risk and varies across the genetic risk distributions.

Importantly, interaction patterns were coherent across multiple behaviors and robust to alternative lifestyle definitions and cohorts.

The endotype-specific patterns observed in this study may suggest biologically plausible specificity in how distinct obesity pathways interact with particular lifestyle behaviors. Notably, the metabolically unhealthy endotype consistently exhibited sensitivity across multiple favorable lifestyle domains, including physical activity, sedentary behavior, diet, and sleep. This pattern aligns with the biological features of this endotype, which is characterized by insulin resistance, dyslipidemia, and systemic metabolic dysfunction, pathways that are broadly responsive to behavioral modulation. In contrast, several endotypes demonstrated more selective lifestyle sensitivity, suggesting pathway-specific interactions rather than generalized responsiveness. For example, the hyperinsulin endotypes showed pronounced modification by favorable physical activity but comparatively weaker or absent effects for favorable sedentary behavior, diet, or sleep. Given the established role of skeletal muscle glucose uptake and insulin sensitivity in mediating the metabolic benefits of physical activity, this could suggest that exercise-centered interventions may be particularly effective for individuals whose genetic risk is dominated by hyperinsulinemic mechanisms. Similarly, the hypoinsulin 2 endotype exhibited stronger modification by sleep relative to other behaviors, consistent with evidence linking favorable sleep with beta-cell function, insulin secretion, and neuroendocrine control of energy balance.^66^ These findings are speculative, however, and do not currently support prescribing endotype-specific lifestyle recommendations. Nevertheless, they raise the possibility that prioritizing specific behavioral targets, such as sleep optimization for hypoinsulinemic pathways or physical activity for hyperinsulinemic pathways, could enhance the effectiveness and personalization of obesity prevention strategies.

Clinically, our findings have important implications for obesity prevention and counseling. Current lifestyle recommendations are largely delivered as uniform guidance, despite wide inter-individual variability in response.^7,8,14^ However, our results suggest that prevention efforts may be more effective when aligned with underlying biological susceptibility. Furthermore, these targeted intervention approaches may be clinically meaningful when contextualized against established lifestyle intervention benchmarks. A recent large meta-analysis reported average BMI reductions of approximately 0.3 to 1.0 kg/m^2^ across dietary and physical activity interventions, effect sizes that are considered clinically relevant at both individual and population levels.^67^ In the present study, favorable sedentary behavior, physical activity, or sleep among those one standard deviation above the mean endotype score was associated with BMI differences of ∼0.1 kg/m^2^; although modest in isolation, differences across multiple endotypes may cumulatively approach the magnitude of average BMI reductions reported for lifestyle interventions. In addition, BMI differences of comparable magnitude were observed even between individuals at the top versus bottom extremes of a single endotype, such as between those in the highest versus lowest decile of the Metabolically unhealthy endotype engaging in favorable sedentary behavior. Additionally, lifestyle-associated BMI differences among individuals with higher cumulative genetic burden frequently fell within or exceeded this range, particularly for sedentary behavior and physical activity. Importantly, BMI differences of the magnitudes seen in our study have also been associated with meaningful downstream health consequences. Prior population-based studies have demonstrated that each 0.1-unit increase in BMI is associated with approximately a 0.5-1.3% higher risk of cardiovascular multimorbidity, with evidence of curvilinear risk acceleration at higher BMI levels.^68,69^ In this context, the genetically amplified lifestyle-associated BMI differences observed here may translate into clinically relevant differences in long-term cardiometabolic risk.

To translate these findings into clinical practice, prospective validation will be essential. Randomized clinical trials that incorporate endotype-specific polygenic scores into study design could assess whether genetically informed prioritization of lifestyle interventions improves weight-related and cardiometabolic outcomes compared with standard, uniform counseling strategies. For example, trials could test whether prioritizing physical activity among individuals with hyperinsulinemic endotypes or sleep optimization among hypoinsulinemic pathways yields greater BMI reduction than standard advice. In parallel, longitudinal cohort studies with repeated lifestyle measurements would clarify whether endotype-specific lifestyle responsiveness persists over time and predicts incident cardiometabolic disease. Finally, integration of genetic sub-phenotyping into implementation research frameworks will be necessary to determine feasibility, cost-effectiveness, and scalability in real-world clinical settings. Together, these steps would move the field from observational gene–lifestyle interaction toward actionable precision-prevention strategies.

Our study has limitations. First, lifestyle behaviors were self-reported at baseline and subsequently dichotomized, which may introduce misclassification and attenuate power. Survey-based measures are also inherently subject to recall bias, social desirability bias, and measurement error, and may not fully capture the dynamic or multidimensional nature of behaviors such as physical activity, sedentary time, diet, and sleep. Although UK Biobank includes validated instruments, alternative approaches such as repeated longitudinal assessments, objective accelerometry, wearable-derived sleep metrics, or dietary biomarkers may provide more precise characterization of lifestyle exposures. Nevertheless, the consistency of findings across multiple behaviors and sensitivity analyses mitigates concerns regarding systematic bias. Second, we acknowledge that our understanding of distinct obesity endotypes is still in its early stages, and comprehensive interpretation remains limited by the current state of knowledge.

Nevertheless, by moving beyond a uniform definition of obesity and embracing a framework grounded in heterogeneous risk pathways, we aim to contribute to a more nuanced understanding of this complex condition. While the effort to sub-classify obesity has gained increasing clinical recognition, our study is among the first to operationalize these endotypes and assess how they shape the impact of healthy lifestyle factors, a key component of obesity prevention and management.^9–11^ Third, the UK Biobank cohort is predominantly of European ancestry, and extending this framework to more diverse populations will be essential to ensure generalizability. Finally, the observational design precludes causal inference, and prospective or interventional studies will be required to determine whether genetically informed lifestyle stratification improves long-term outcomes.

This study demonstrates that obesity endotypes modify the relationship between lifestyle behaviors and obesity risk. By integrating genetic pathway information with modifiable exposures, these findings support a shift from uniform lifestyle recommendations toward precision-prevention strategies aligned with individual dominant biological drivers of obesity. As genomic data become increasingly integrated into clinical practice, this framework provides a foundation for translating genetic insight into personalized approaches to obesity prevention.

## Data Availability

All data produced in the present study are available upon reasonable request to the authors.

---

**Supplementary Fig. 1.**
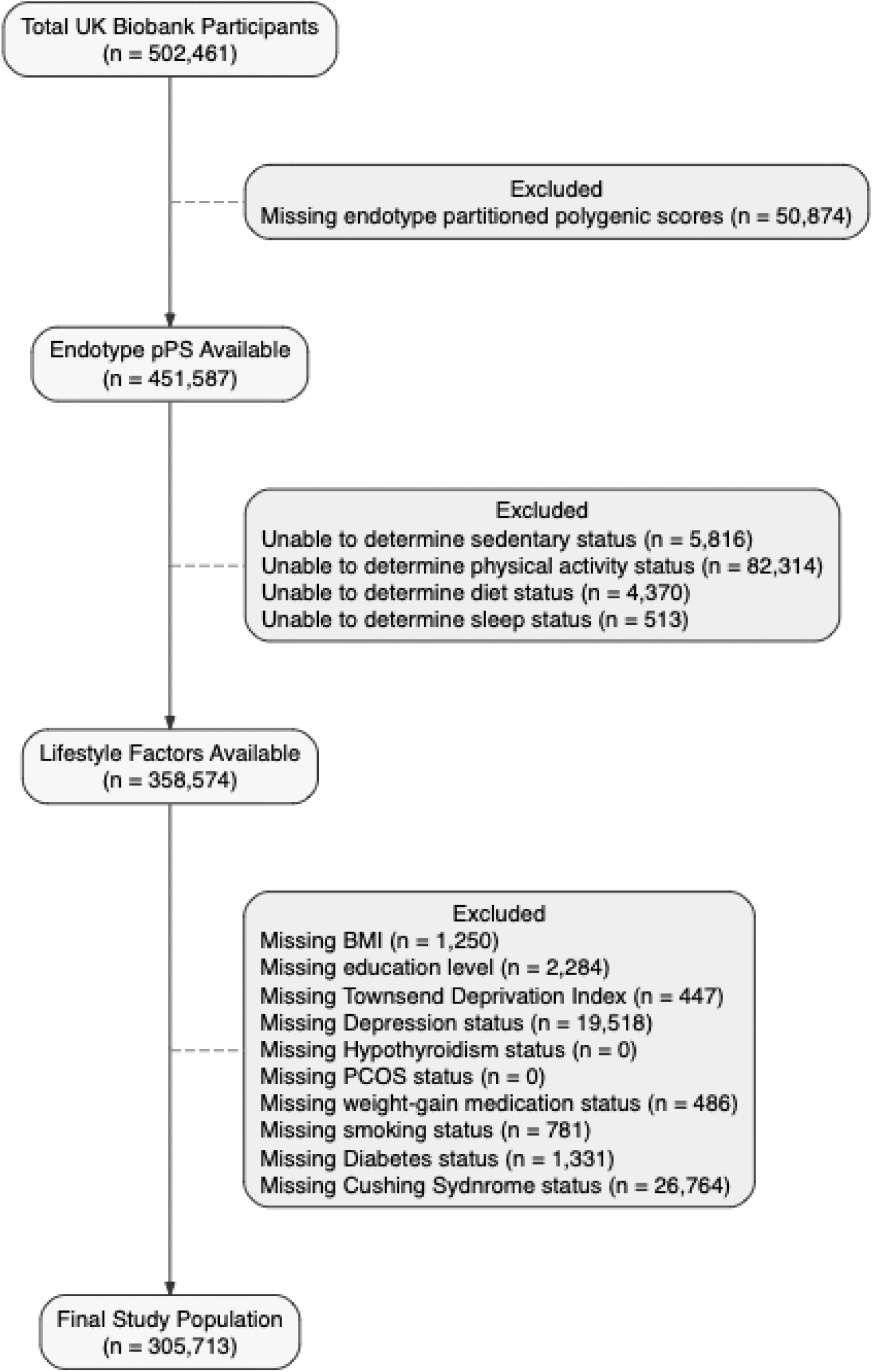
Study Flow Diagram.

**Supplementary Fig. 2.**
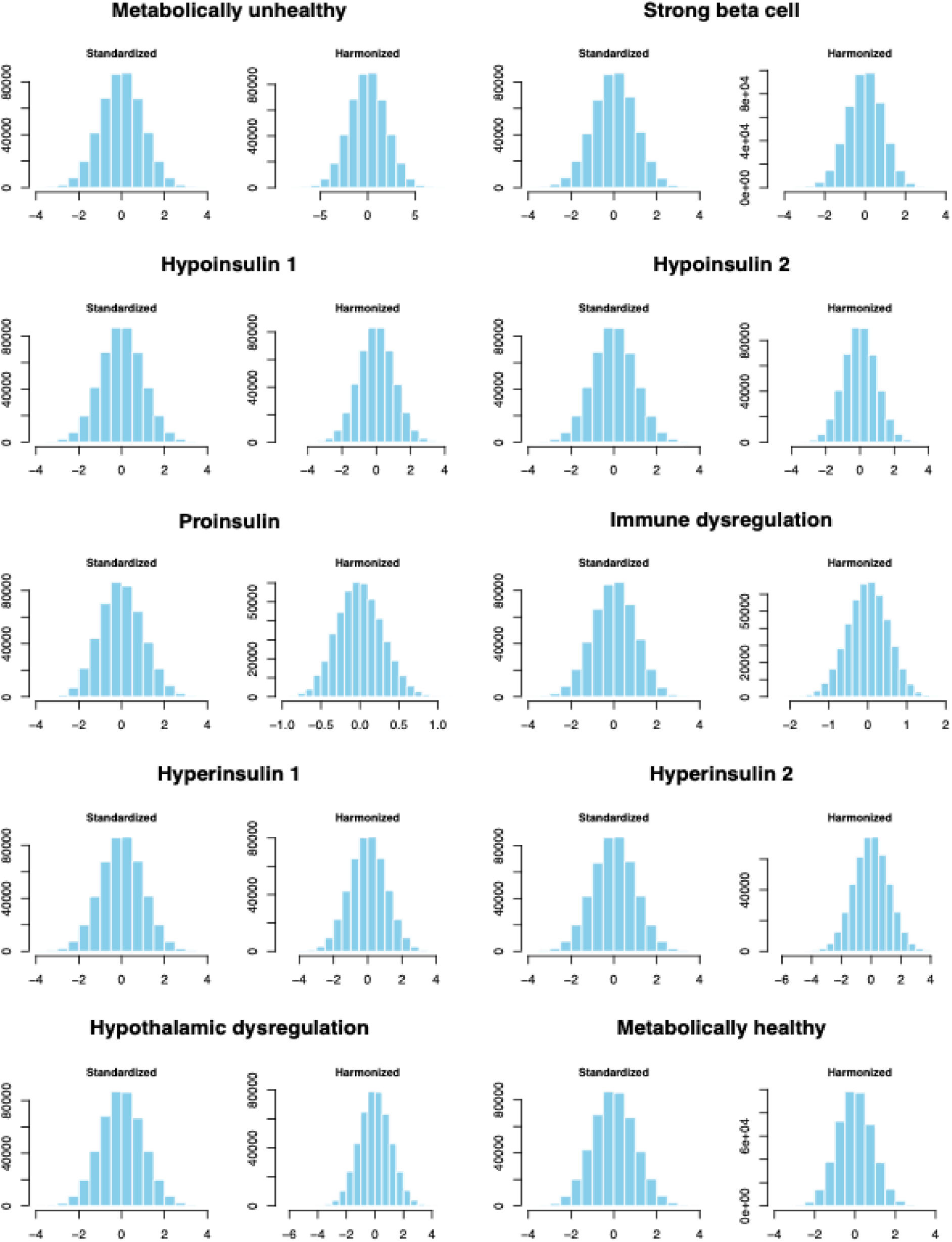
Histograms of standardized and BMI-harmonized pPSs.

**Supplementary Fig. 3.**
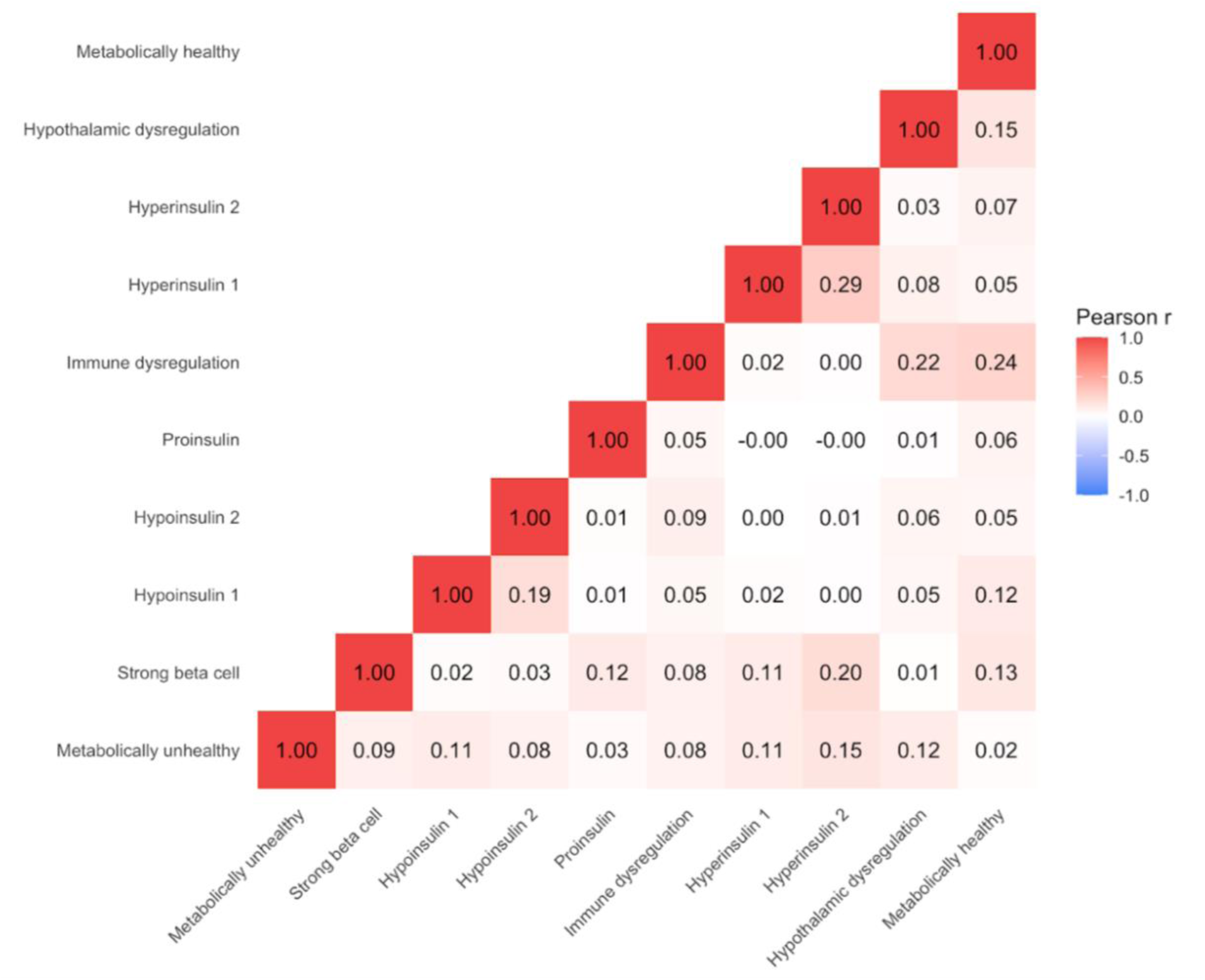
Pearson correlation among endotype-specific pPSs.

**Supplementary Fig. 4.**
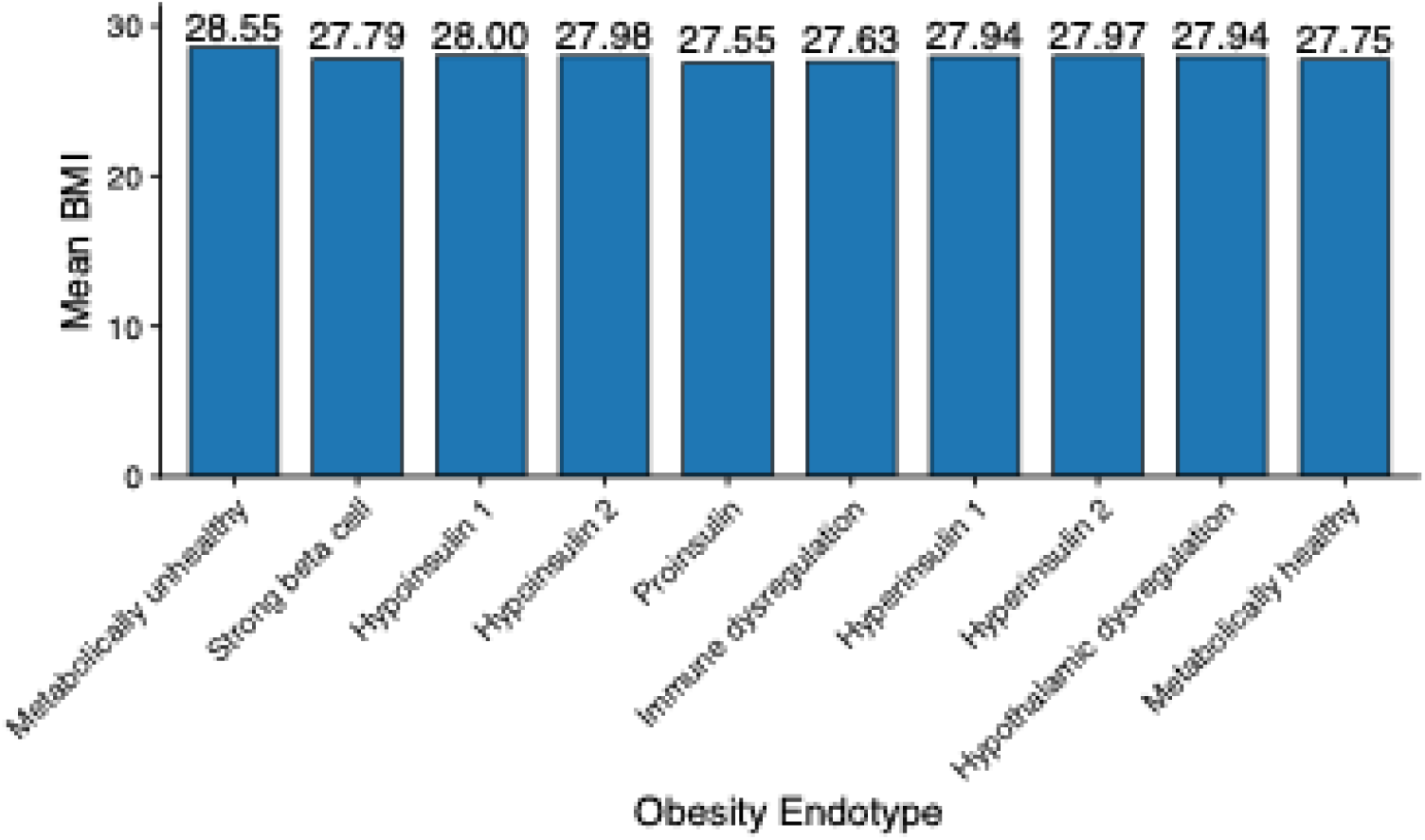
Mean BMI among participants in the top decile of each endotype.

**Supplementary Fig. 5.**
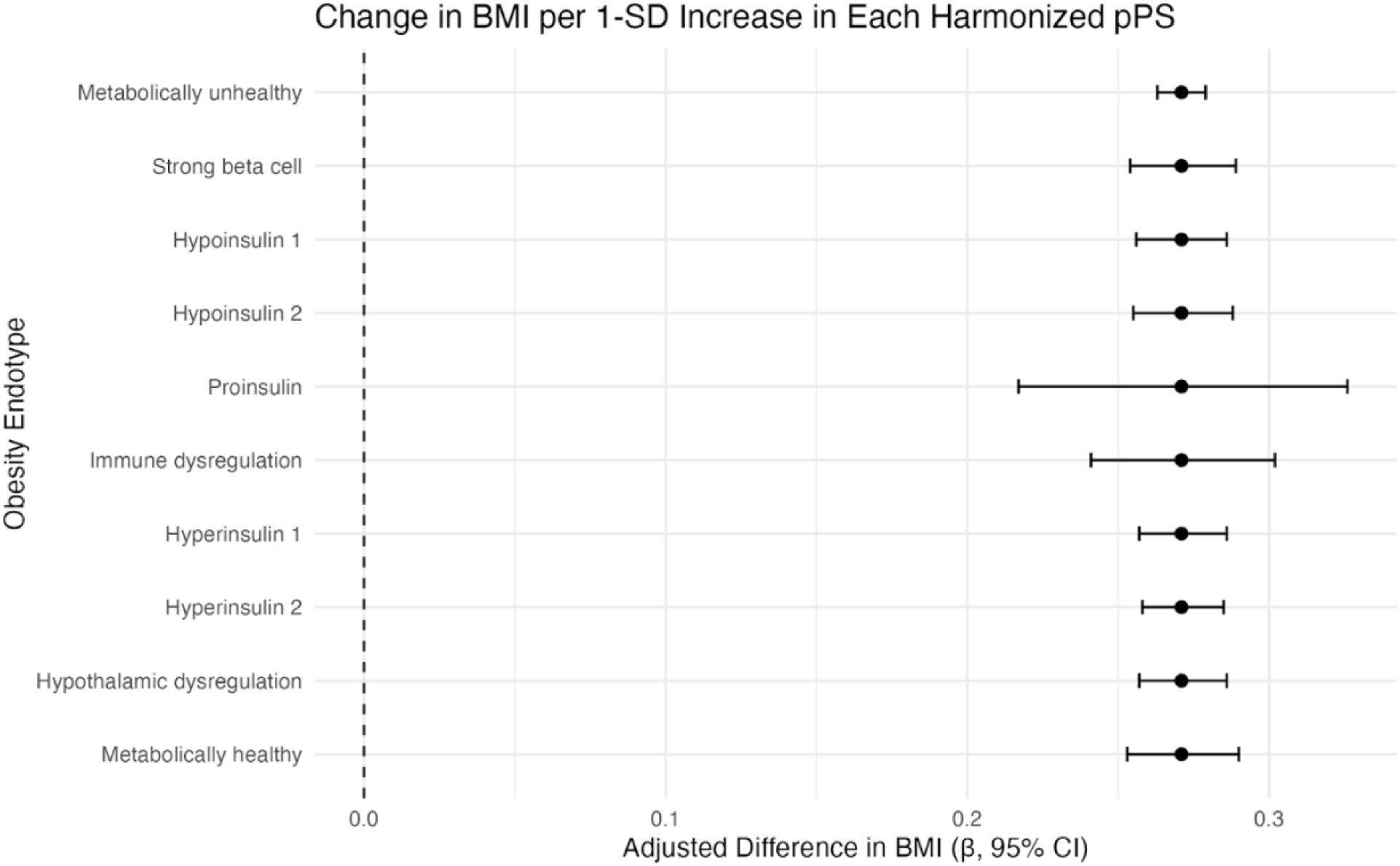
Associations of BMI-harmonized endotype-specific pPSs with BMI.

**Supplementary Fig. 6.**
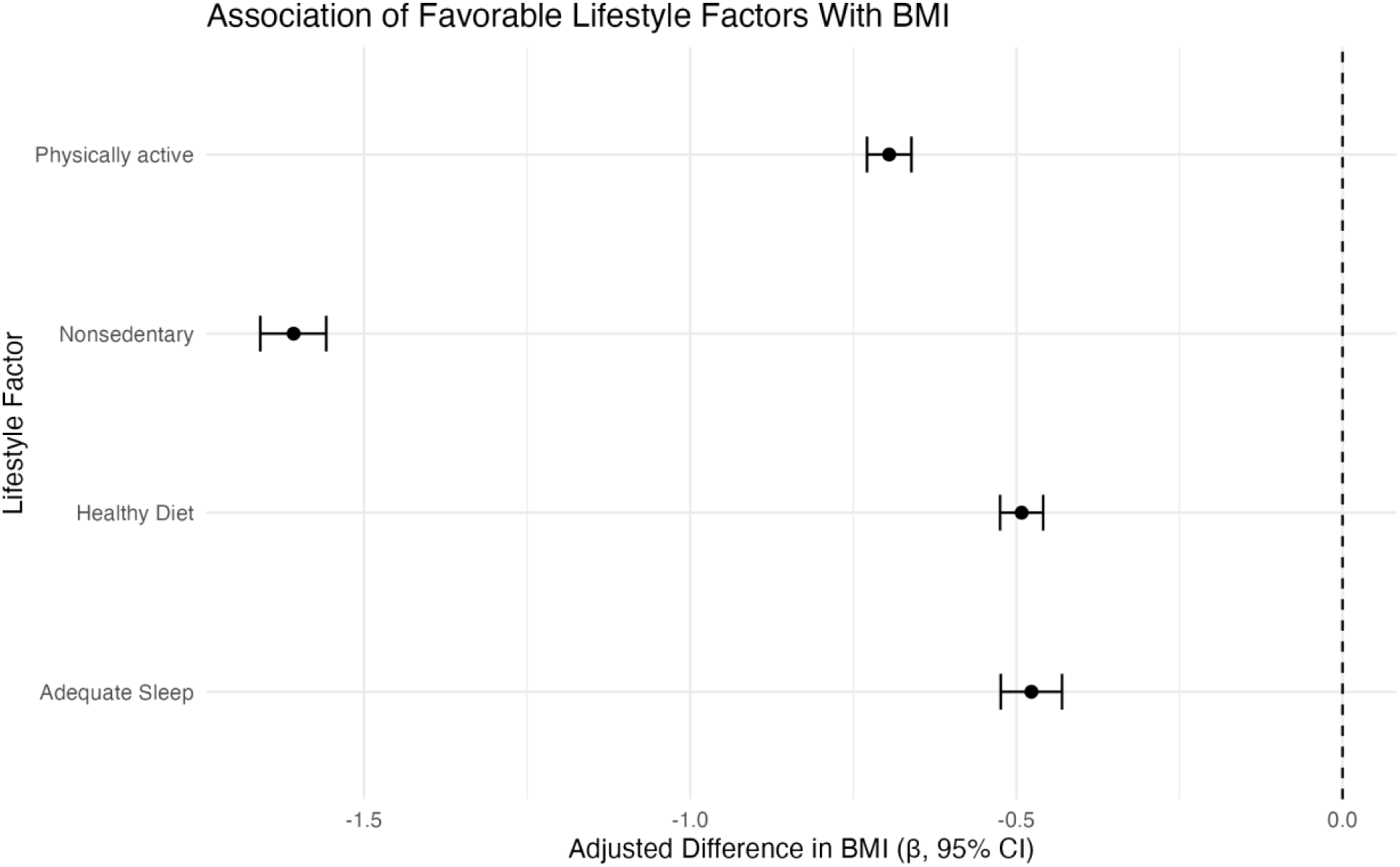
Association between lifestyle factors and BMI.

**Supplementary Fig. 7.**
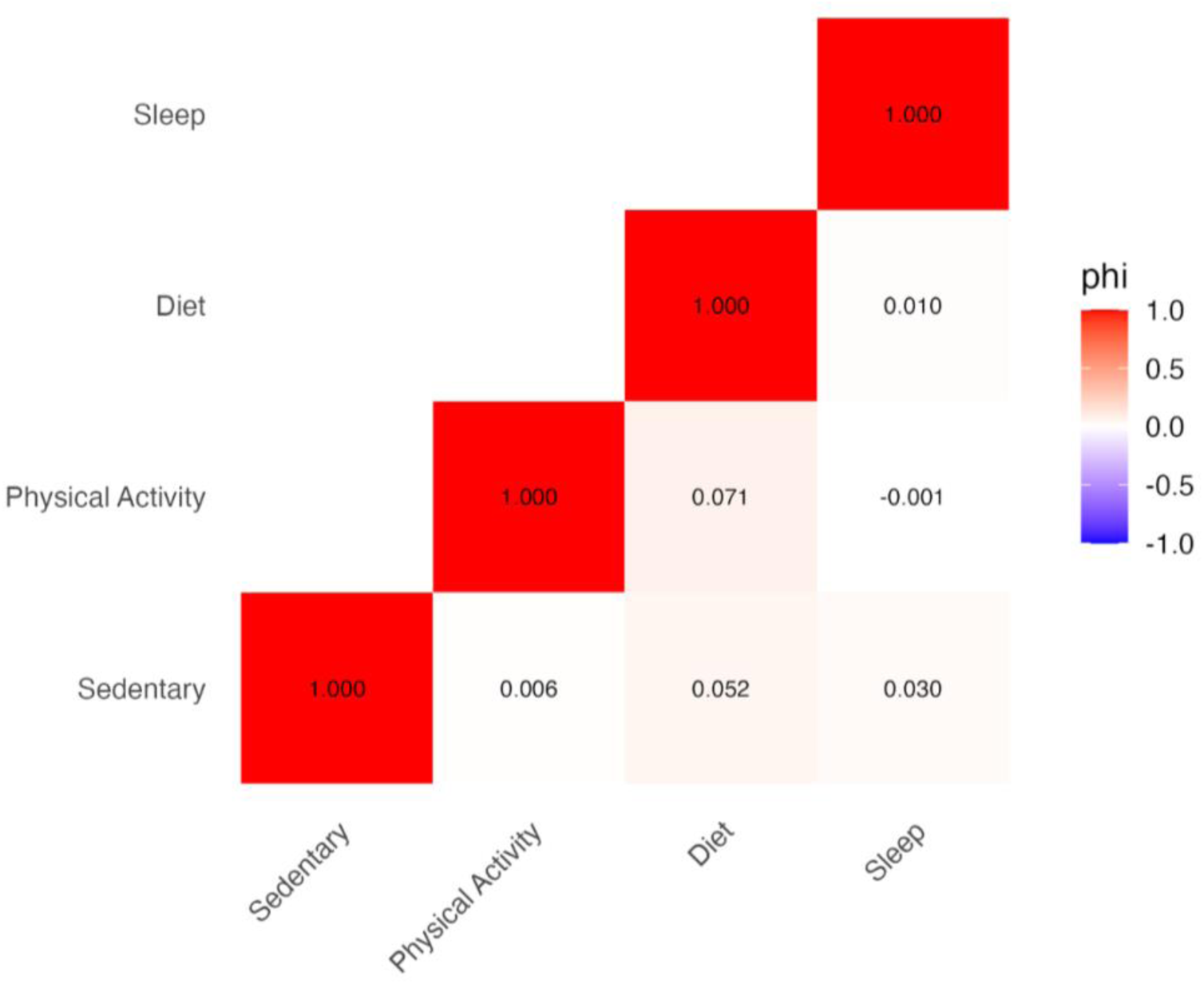
Phi correlation among lifestyle factors.

**Supplementary Fig. 8.**
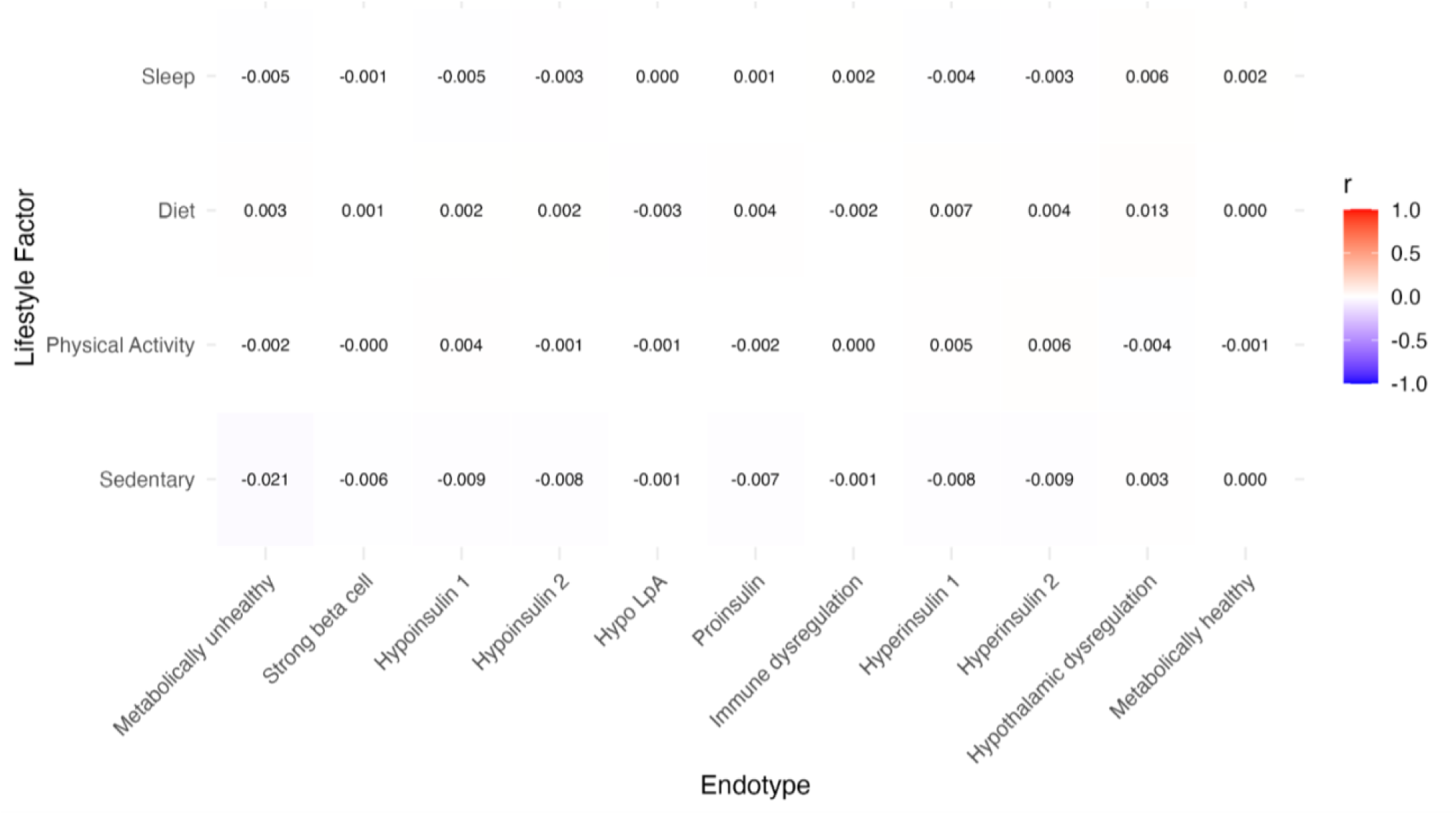
Point-biserial correlation among lifestyle factors and endotype scores.

**Supplementary Fig. 9.**
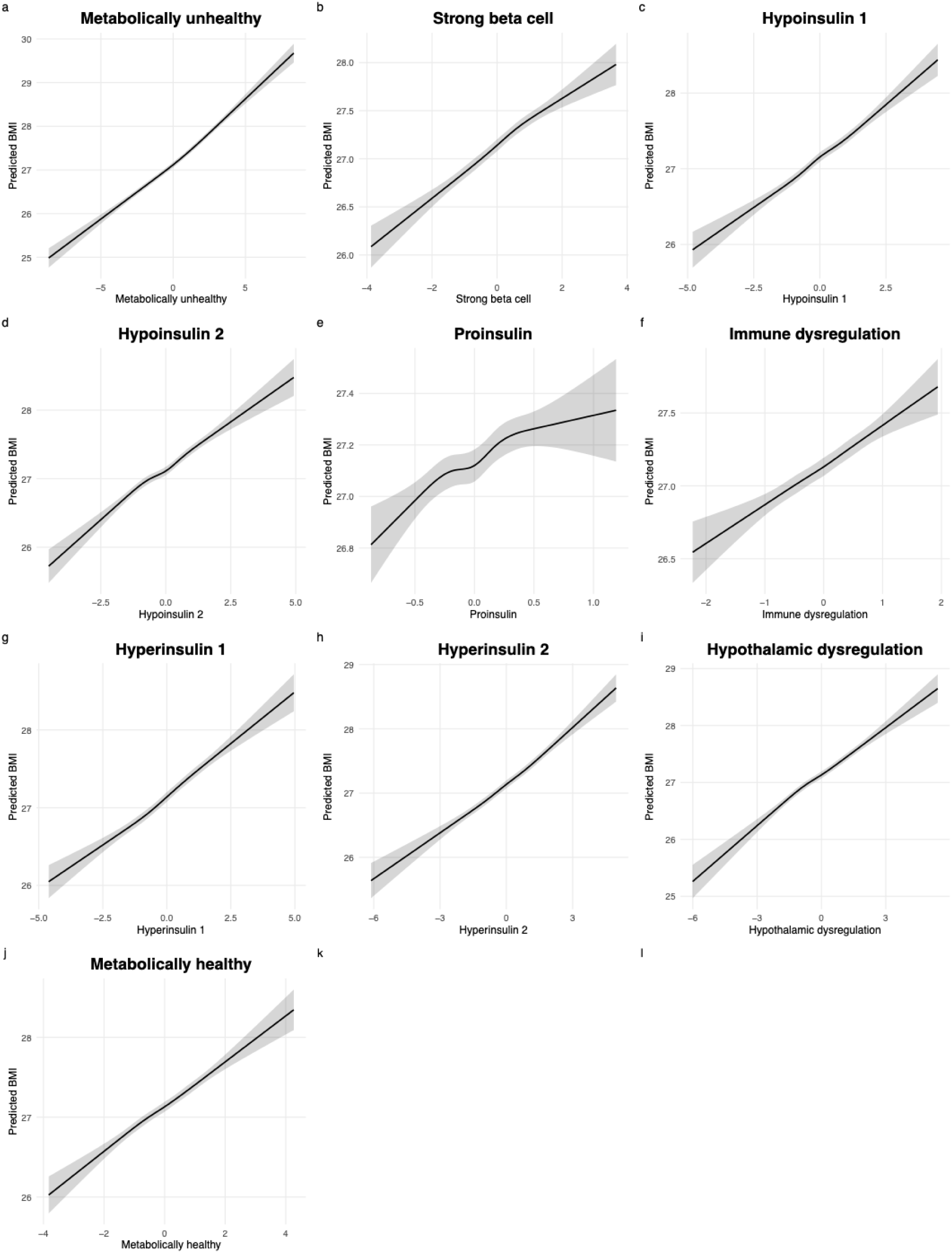
Restricted Cubic Splines to test assumption of linear relationship between continuous partitioned polygenic scores and BMI. Shaded bands represent the 95% confidence interval. Degrees of freedom = 6; 5 knots set at 5%, 27.5%, 50%, 72.5%, and 95%.

**Supplementary Fig. 10.**
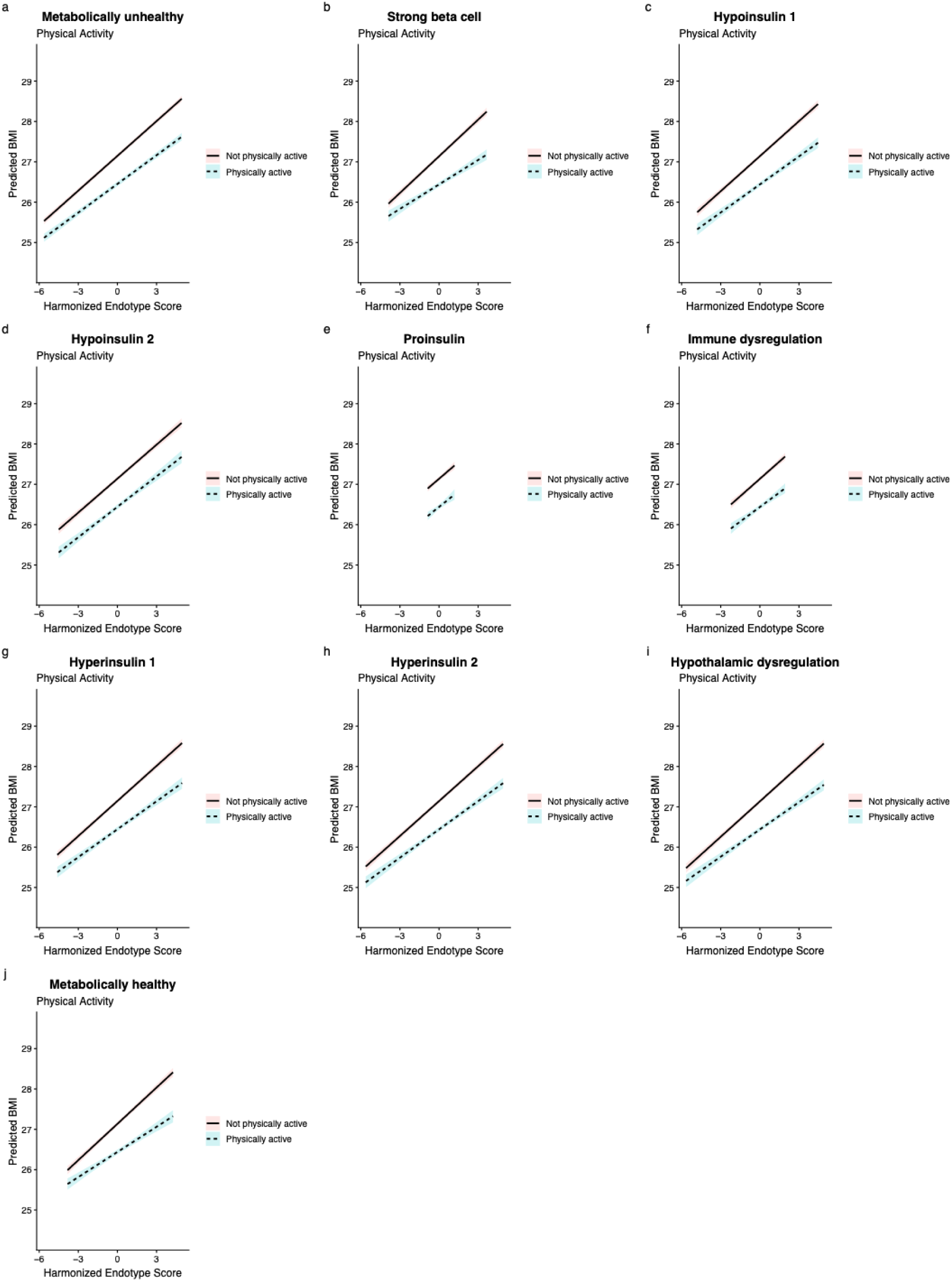
Interaction plots with predicted BMI trajectories stratified by physical activity status for all endotypes. Shaded bands represent the 95% confidence interval.

**Supplementary Fig. 11.**
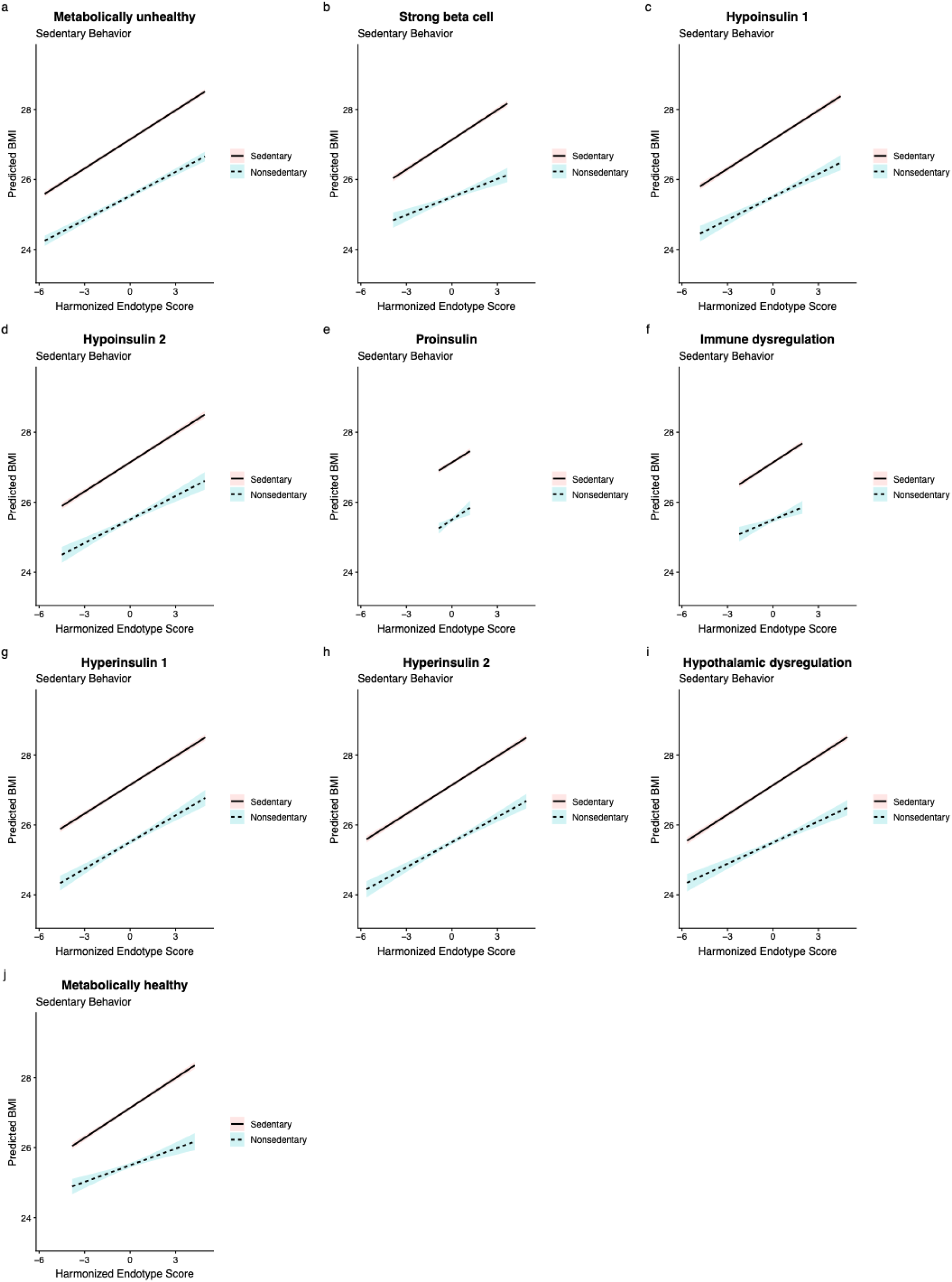
Interaction plots with predicted BMI trajectories stratified by sedentary behavior status for all endotypes. Shaded bands represent the 95% confidence interval.

**Supplementary Fig. 12.**
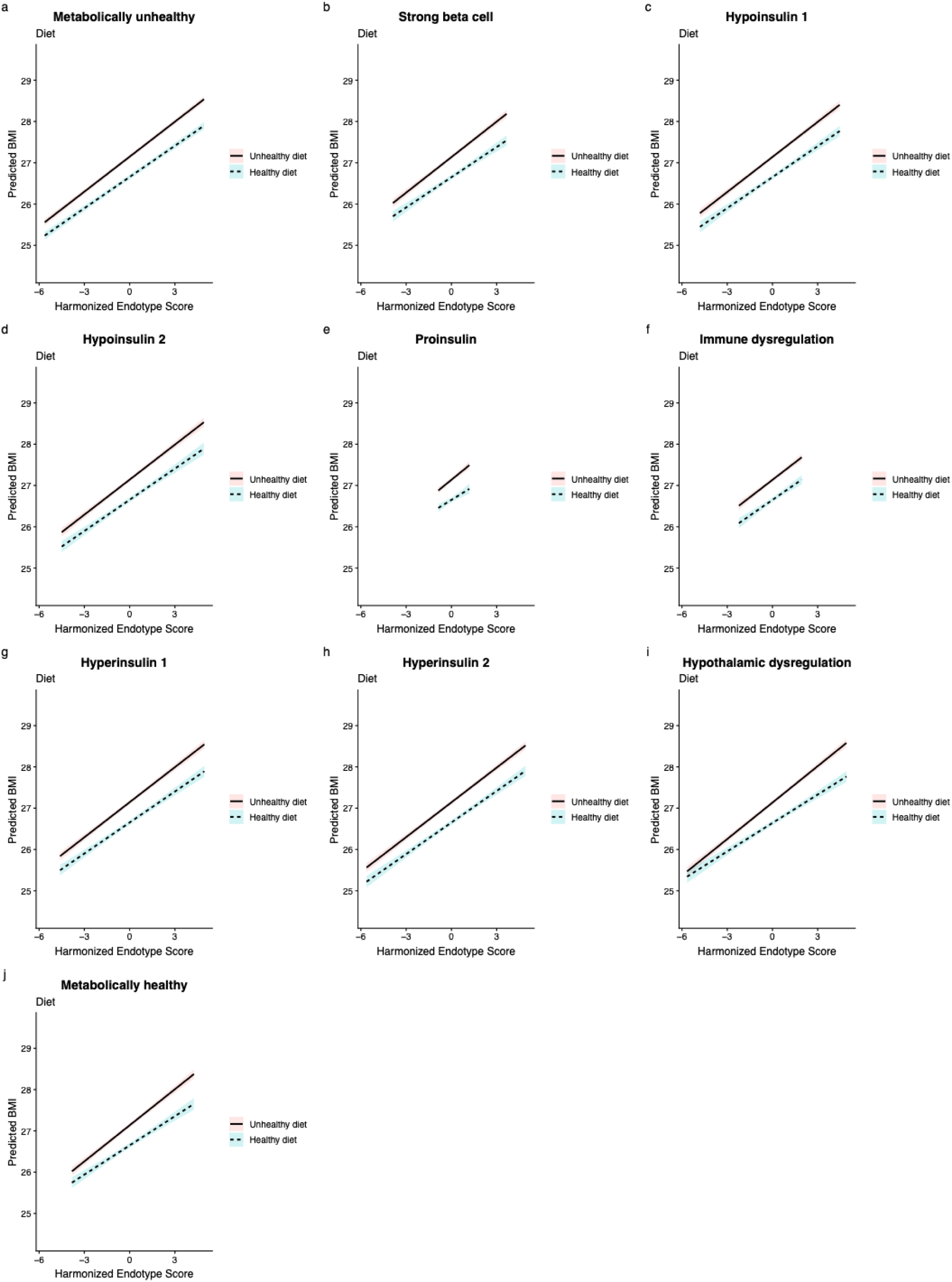
Interaction plots with predicted BMI trajectories stratified by diet status for all endotypes. Shaded bands represent the 95% confidence interval.

**Supplementary Fig. 13.**
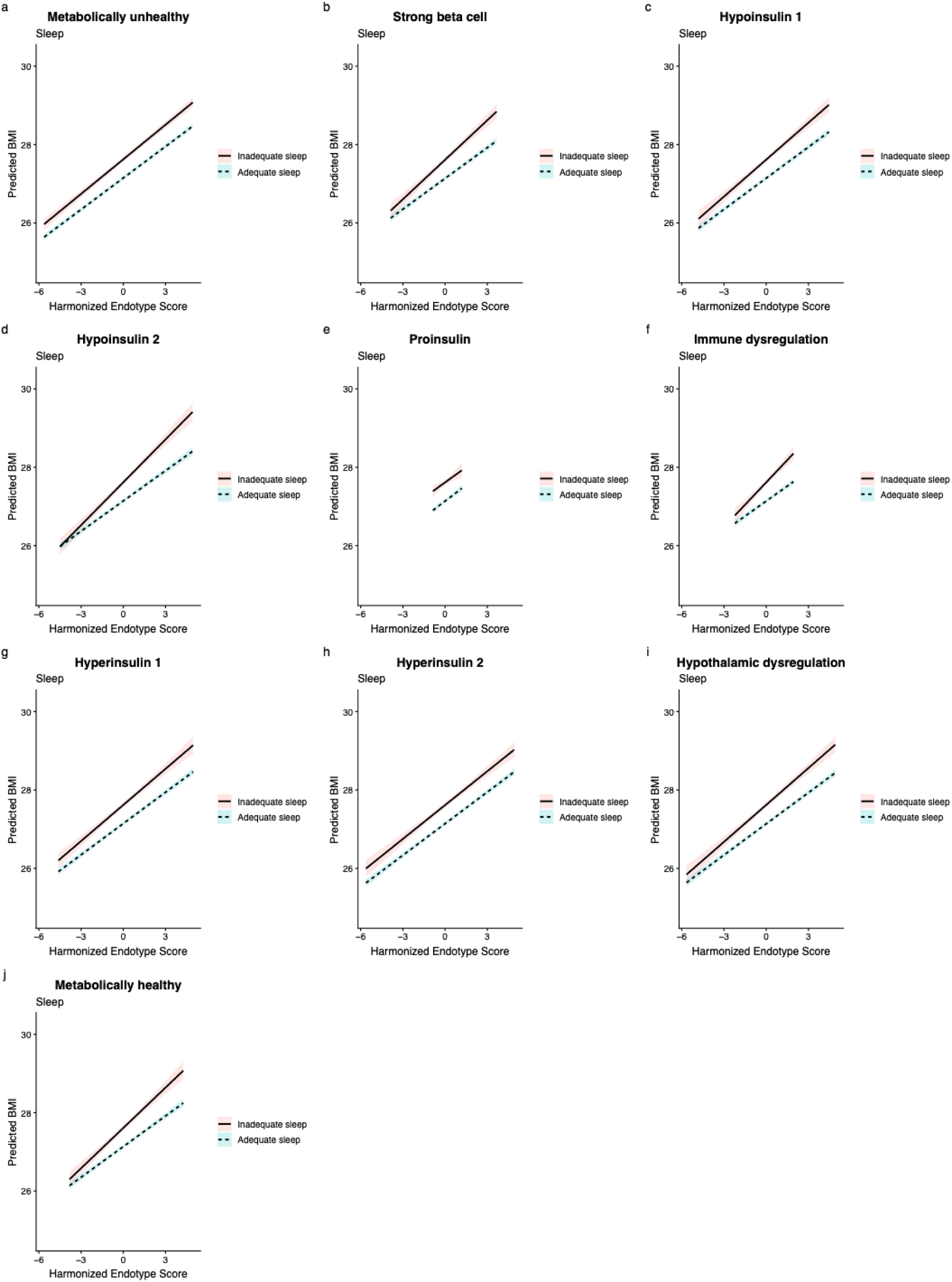
Interaction plots with predicted BMI trajectories stratified by sleep status for all endotypes. Shaded bands represent the 95% confidence interval.

**Supplementary Fig. 14.**
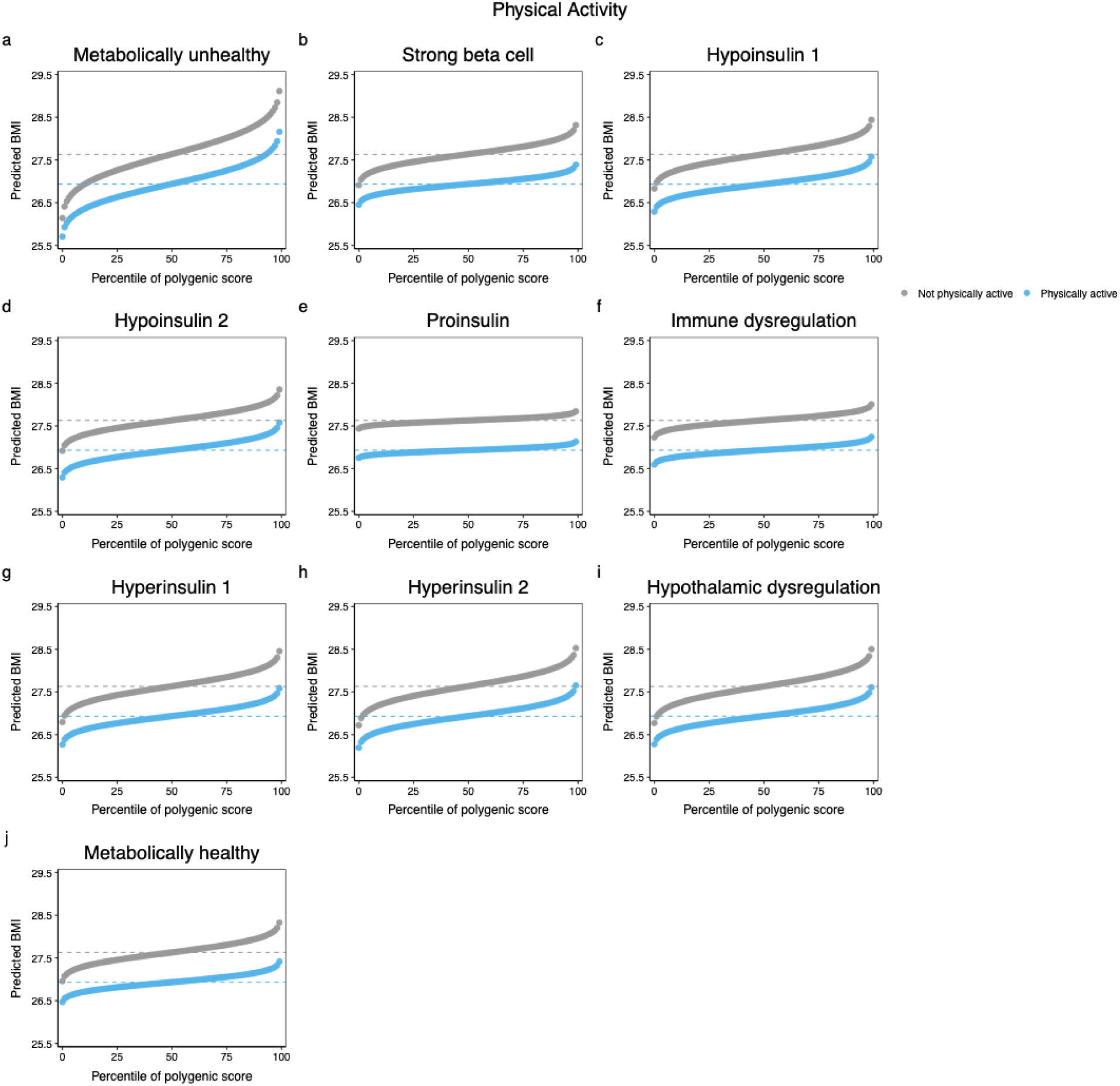
Predicted BMI across genetic risk percentiles for favorable and unfavorable physical activity by obesity endotype.

**Supplementary Fig. 15.**
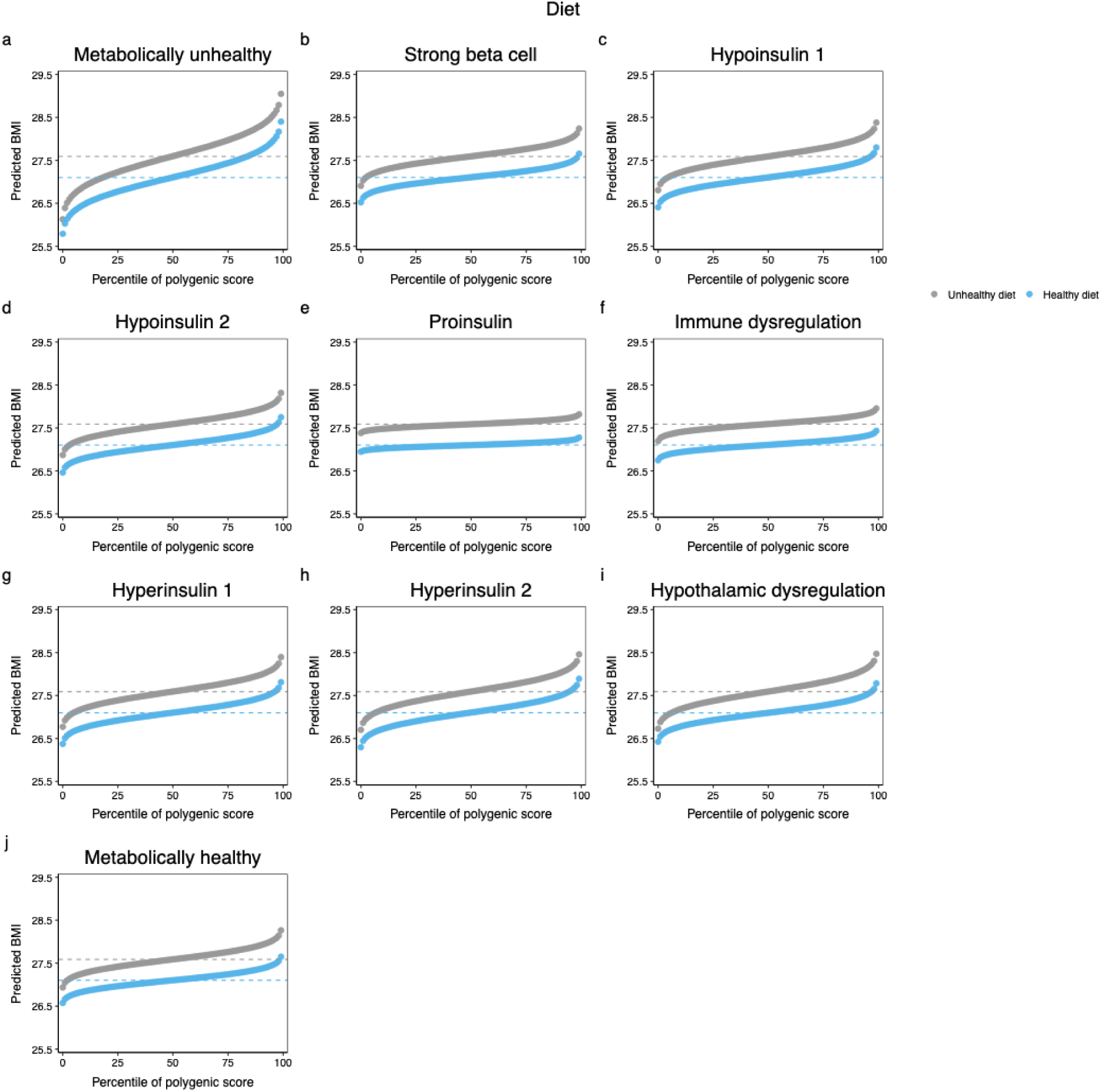
Predicted BMI across genetic risk percentiles for favorable and unfavorable diet by obesity endotype.

**Supplementary Fig. 16.**
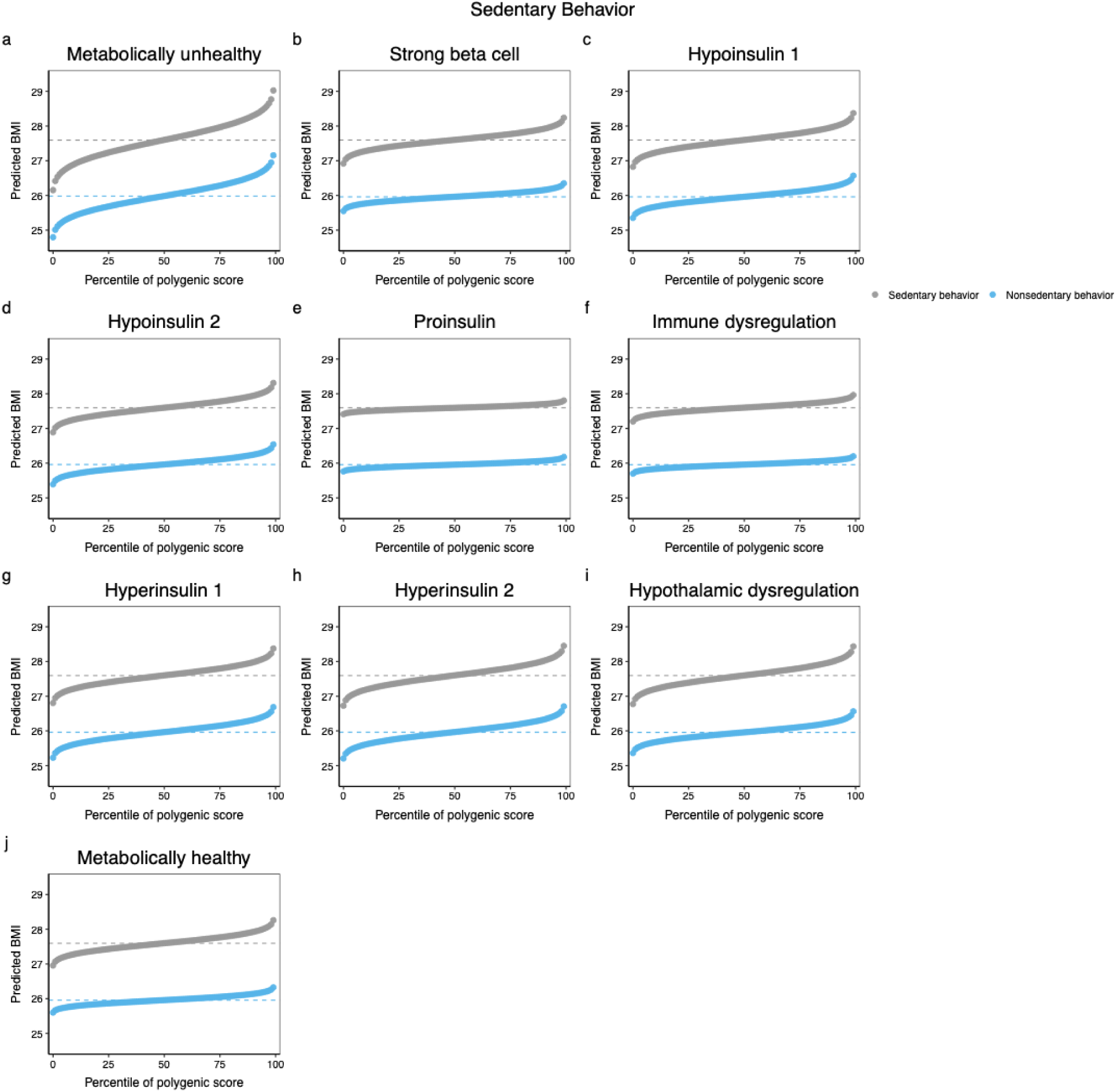
Predicted BMI across genetic risk percentiles for favorable and unfavorable sedentary behavior by obesity endotype.

**Supplementary Fig. 17.**
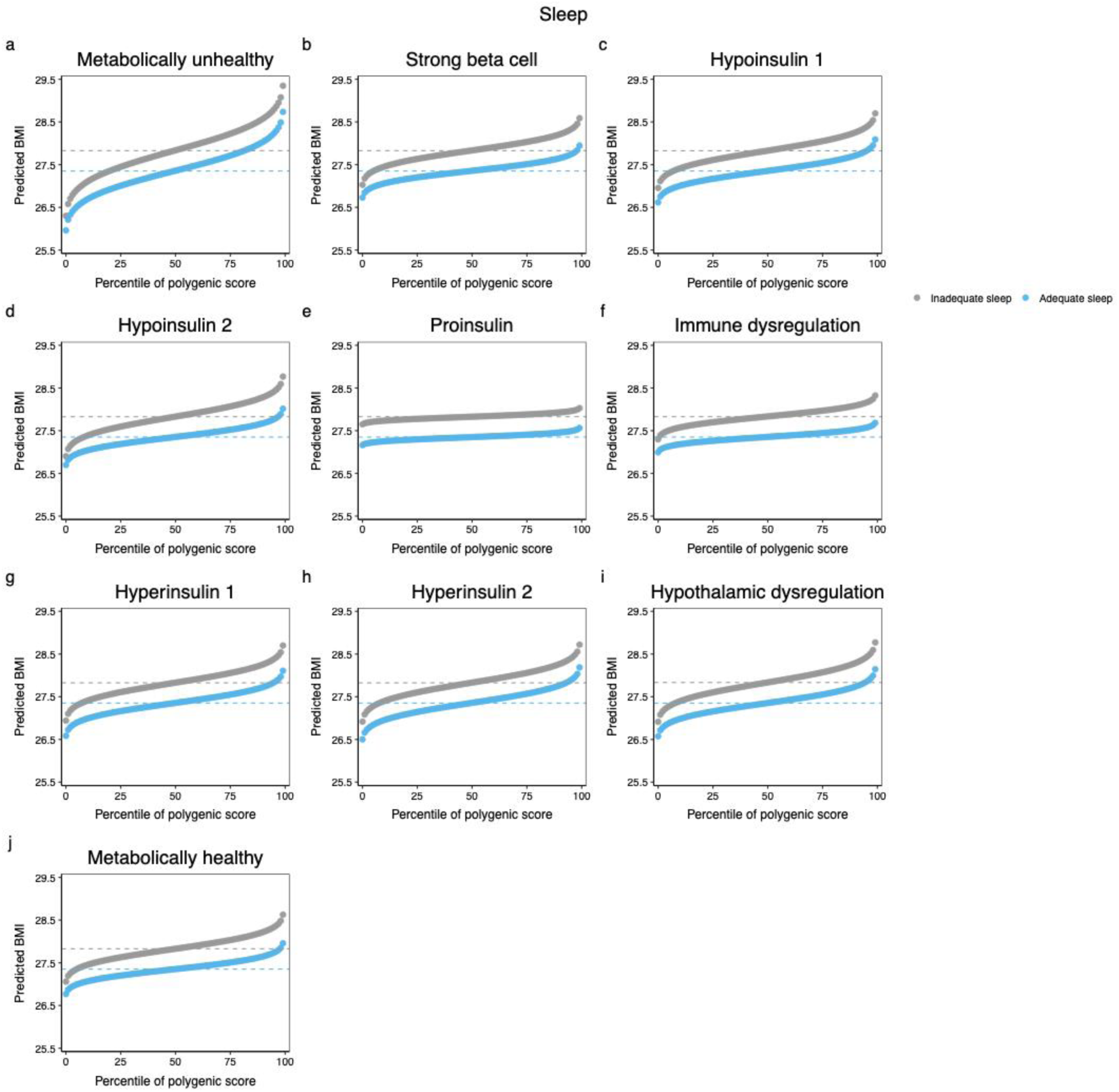
Predicted BMI across genetic risk percentiles for favorable and unfavorable sleep by obesity endotype.

**Supplementary Fig. 18.**
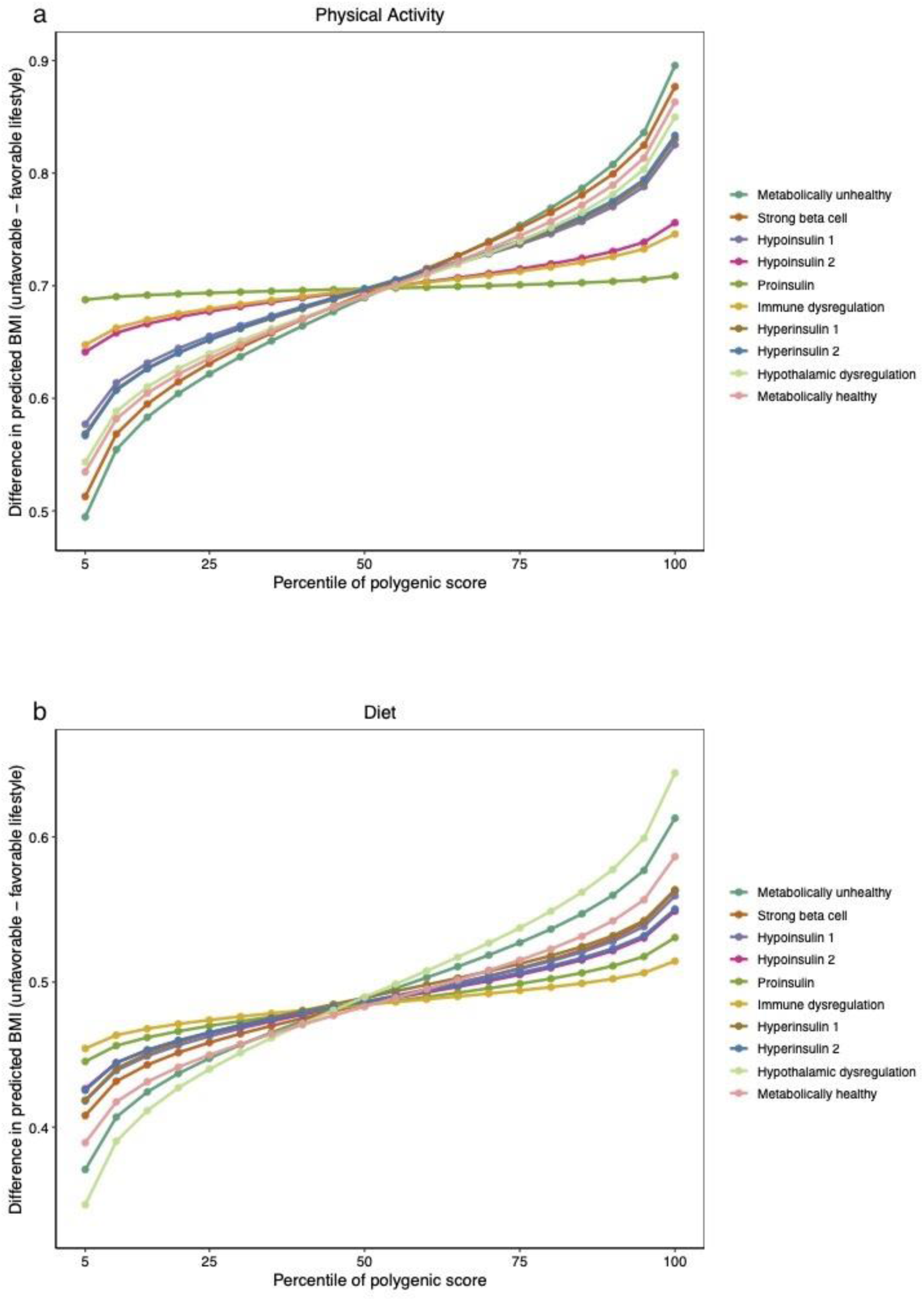
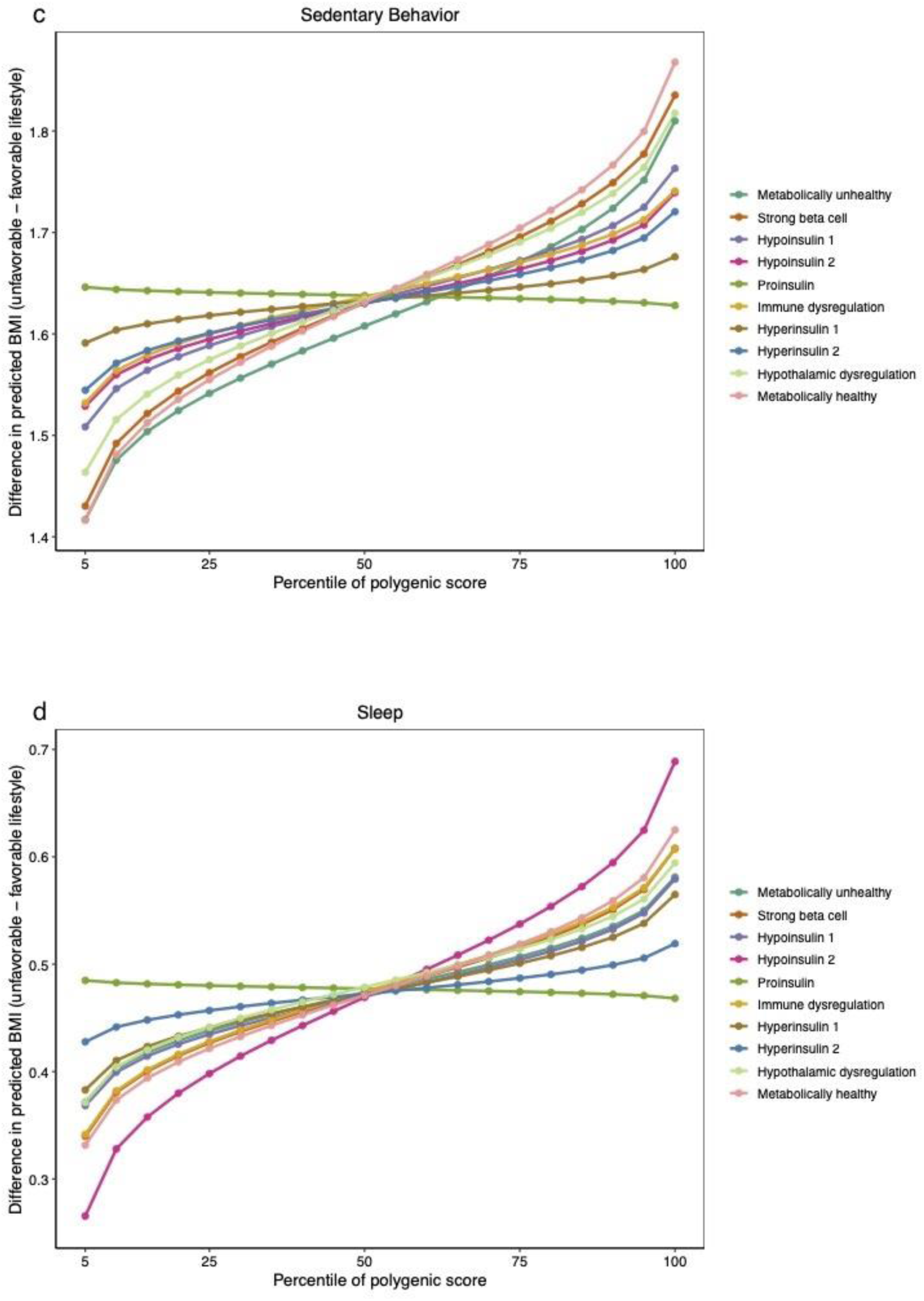
Difference in predicted BMI between favorable and unfavorable (a) physical activity, (b) sedentary behavior, (c) diet, and (d) sleep across genetic risk percentiles by obesity endotype.

**Supplementary Fig. 19.**
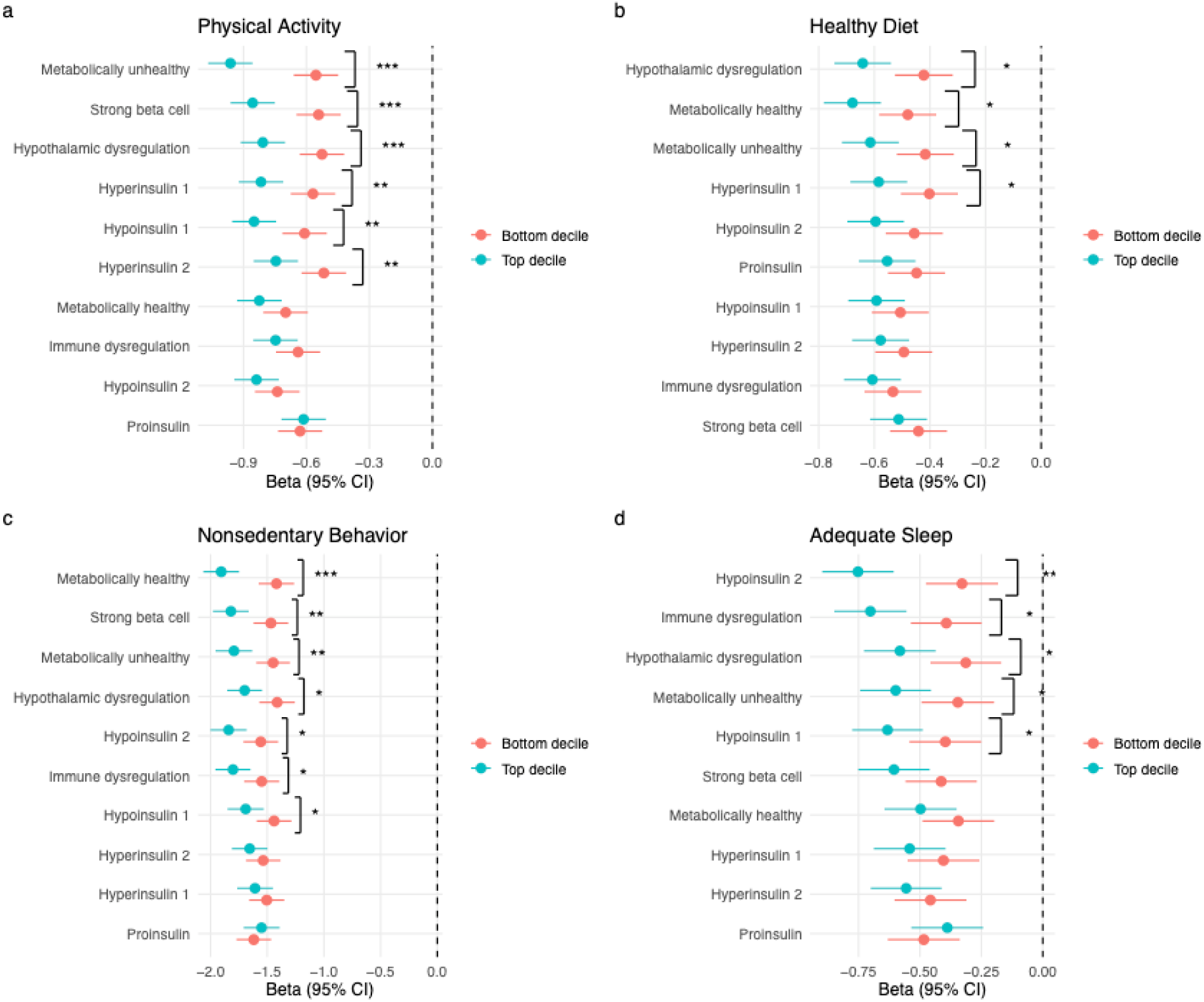
Estimated associations between lifestyle behaviors and BMI for top versus bottom decile of each endotype, ordered by greatest to least interaction term coefficient. Error bars represent the 95% confidence interval. * = p-value < 0.05; ** = p-value < 0.01; *** = p-value < 0.001.

**Supplementary Fig. 20.**
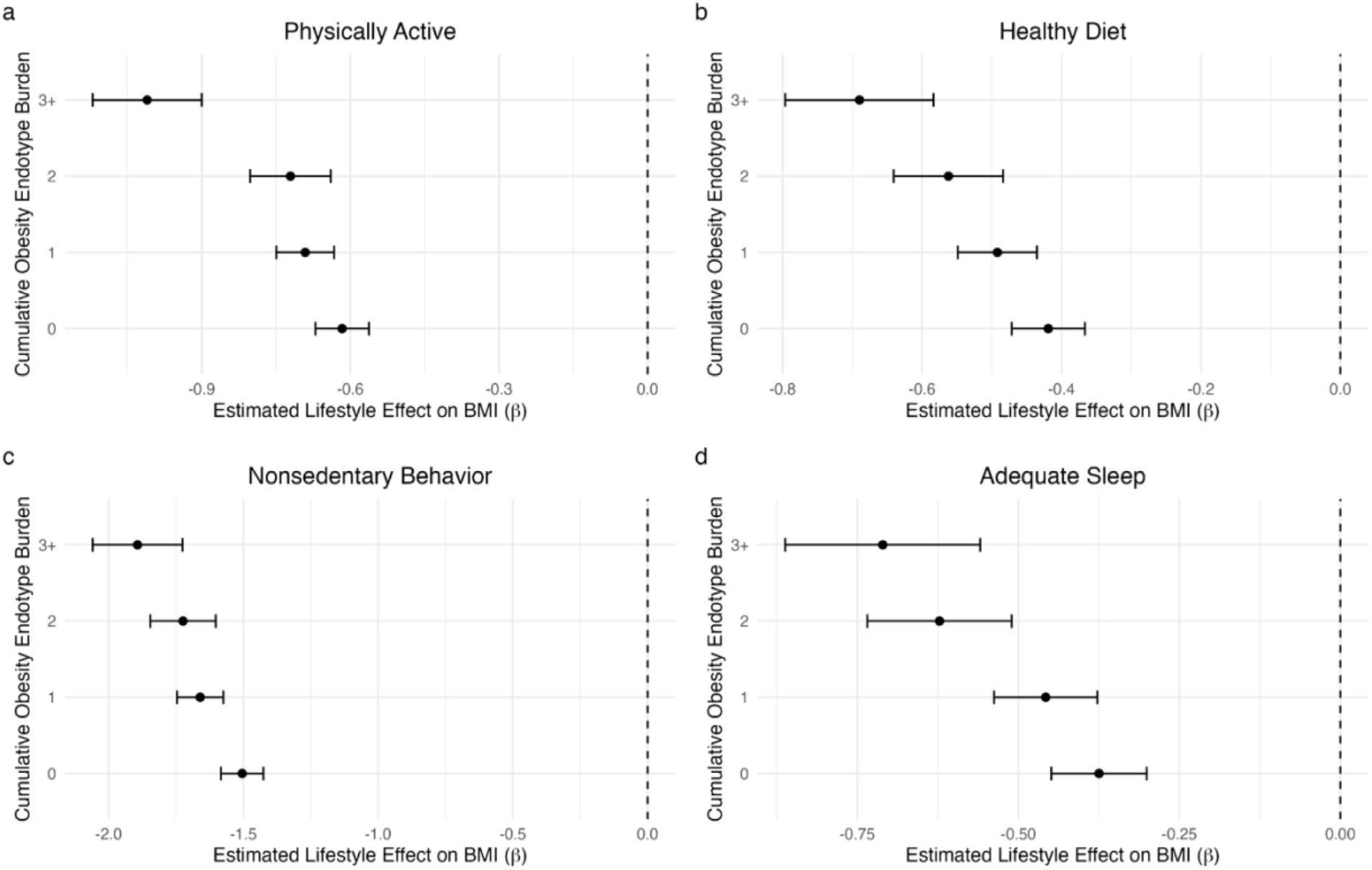
Estimated associations between healthy lifestyle behaviors and BMI conditional on cumulative obesity endotype burden.

**Supplementary Fig. 21.**
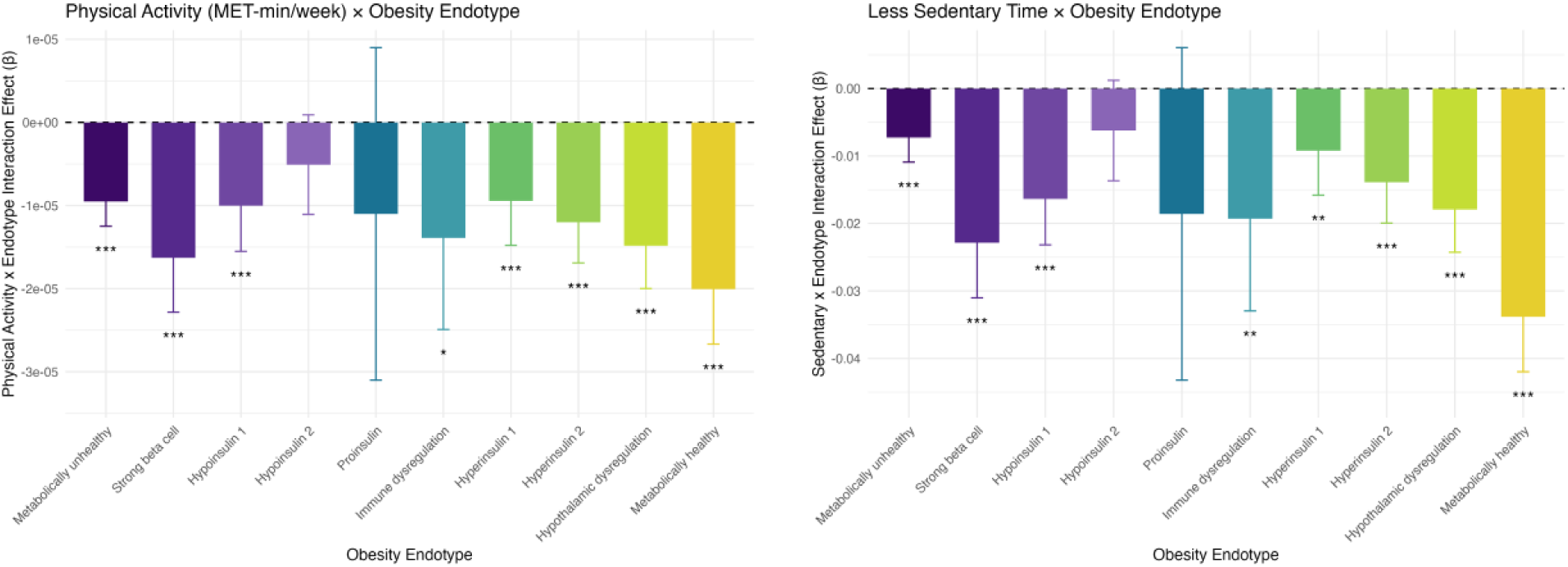
Interaction effects between lifestyle behaviors and harmonized endotype-specific partitioned polygenic scores on BMI, using continuous physical activity and nonsedentary time. Error bars represent the 95% confidence interval. * = p-value < 0.05; ** = p-value < 0.01; *** = p-value < 0.001.

**Supplementary Fig. 22.**
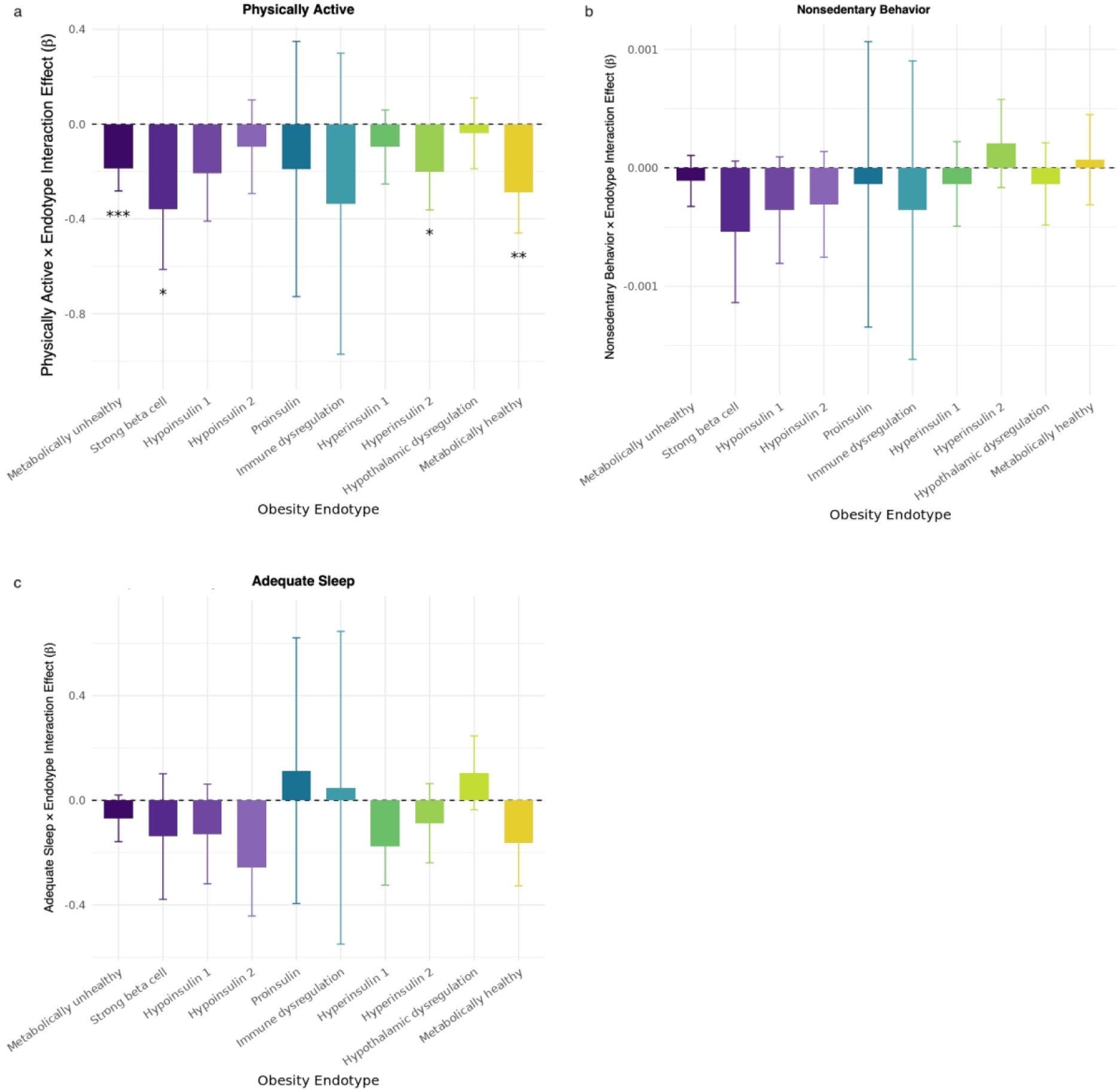
Interaction effects between lifestyle behaviors and obesity endotypes on BMI in the All of Us cohort. * = p-value < 0.05; ** = p-value < 0.01; *** = p-value < 0.001.

**ST1:** Definitions of lifestyle behavior variables.

| Lifestyle Factor | Definition (1 = favorable; 0 = unfavorable) |
| --- | --- |
| Physical Activity | Score 1 if the total MET minutes per week for all activities (Data-Field 22040) are greater than or equal to 3000 minutes per week, otherwise 0. |
| Diet | Score 1 if more than 4 of the 8 criteria below are met, otherwise 0.<br>1: Fruits $\geq 3$ servings/day (Data-Field 1309, 1319)<br>2: Vegetables $\geq 3$ servings/day (Data-Field 1289, 1299)<br>3: Whole grains $\geq 3$ servings/day (Data-Field 1438, 1448, 1458, 1468)<br>4: Refined grains $\leq 1.5$ servings/day (Data-Field 1438, 1448, 1458, 1468)<br>5: Fish $\geq 2$ servings/week (Data-Field 1329, 1339)<br>6: Processed meats $\leq 1$ serving/week (Data-Field 1349)<br>7: Unprocessed red meats $\leq 1.5$ servings/week (Data-Field 1369, 1379, 1389)<br>8: Never eat sugar-sweetened foods or beverages (Data-Field 6144) |
| Sedentary behavior | Score 1 if the sum of the hours below is less than 2, otherwise 0.<br>1: Time spent watching television (TV) (Data-Field 1070)<br>2: Time spent using computer (not including computer use at work) (Data-Field 1080) |
| Sleep | Score 1 if sleep duration is 6 to 8 hours/day, otherwise 0. (Data-Field 1160) |

**ST1: Definitions of lifestyle behavior variables.**

**ST2:** Definitions of covariate phenotypes.

| Phenotype | ICD-9 | ICD-10 | Self-reported data-fields (codes) |
| --- | --- | --- | --- |
| Depression | 311 | F32, F32.0, F32.1, F32.2, F32.3, F32.8, F32.9, F33, F33.0, F33.1, F33.2, F33.3, F33.4, F33.8, F33.9 | 20002 (1286) |
| Hypothyroidism | 244, 2442, 2449, 24499 | E03, E03.0, E03.1, E03.2, E03.3, E03.5, E03.8, E03.9 | 20002 (1226) |
| PCOS | 2564 OR [(626, 6260, 62601, 62602, 62609, 6261, 6263, 6264, 6265, 6266, 6267, 6268, 6269) & (7041 or ICD-10 L68.0)] | E28.2 | 20002 (1350) |
| Type 2 Diabetes Mellitus | – | E11, E11.0, E11.1, E11.2, E11.3, E11.4, E11.5, E11.6, E11.7, E11.8, E11.9 | – |
| Cushing Syndrome | – | E24, E24.0, E24.1, E24.2, E24.3, E24.4, E24.8, E24.9 | – |

**ST2: Definitions of covariate phenotypes.**

**ST3:**
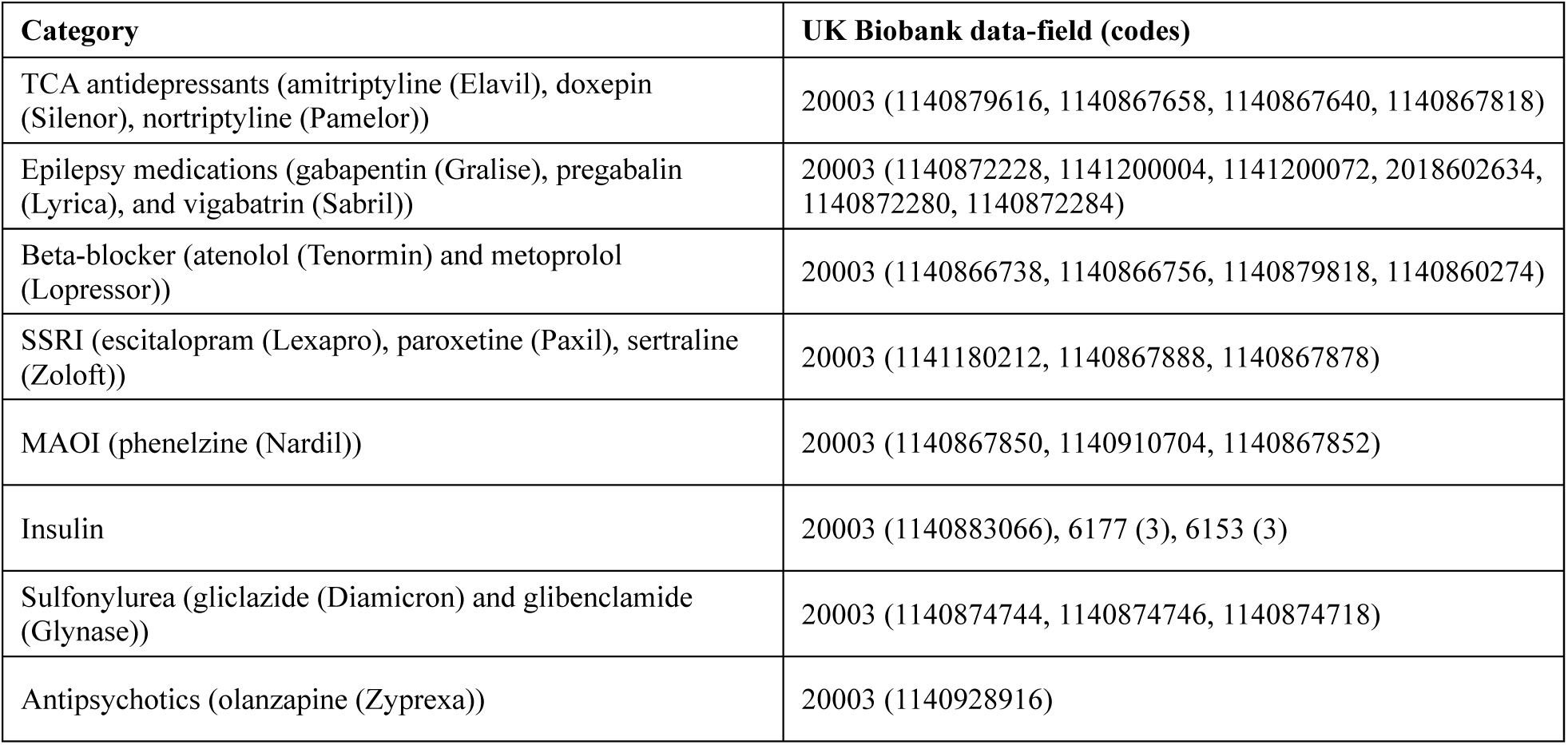
Definition of weight-gaining medications.

**ST4:** Cohort characteristics.

ST3: Definition of weight-gaining medications
|  | **Total**<br>**Population** | **Men** | **Women** |
| --- | --- | --- | --- |
|  | **n= 305,713** | **n = 145,713** | **n = 160,000** |
| **Age in years, mean (SD)** | 56.8 (8.0) | 57.3 (8.0) | 56.3 (8.0) |
| **BMI in kg/m2, mean (SD)** | 27.4 (4.7) | 27.9 (4.2) | 27.0 (5.1) |
| **Lifestyle Factors, no. (%)** |  |  |  |
| Sedentary (favorable) | 34,035 (11.1%) | 12,958 (8.9%) | 21,077 (13.2%) |
| Physical Activity (favorable) | 94,817 (31.0%) | 47,448 (32.6%) | 47,369 (29.6%) |
| Diet (favorable) | 109,569 (35.8%) | 42,762 (29.3%) | 66,807 (41.8%) |
| Sleep (favorable) | 265,733 (86.9%) | 127,317 (87.4%) | 138,416 (86.5%) |
| **Composite Top Deciles, no. (%)** |  |  |  |
| 0 | 120,582 (39.4%) | 57,113 (39.2%) | 63,469 (39.7%) |
| 1 | 103,466 (33.8%) | 49,194 (33.8%) | 54,272 (33.9%) |
| 2 | 52,990 (17.3%) | 25,507 (17.5%) | 27,483 (17.2%) |
| 3+ | 28,675 (9.4%) | 13,899 (9.5%) | 14,776 (9.2%) |
| **No. of healthy lifestyle factors, no. (%)** |  |  |  |
| 0 | 17,295 (5.7%) | 8,750 (6.0%) | 8,545 (5.3%) |
| 1 | 125,090 (40.9%) | 64,059 (44.0%) | 61,031 (38.1%) |
| 2 | 115,622 (37.8%) | 53,886 (37.0%) | 61,736 (38.6%) |
| 3 | 43,004 (14.1%) | 17,418 (12.0%) | 25,586 (16.0%) |
| 4 | 4,702 (1.5%) | 1,600 (1.1%) | 3,102 (1.9%) |
| **Education, no. (%)** |  |  |  |
| Higher education | 121,638 (39.8%) | 58,339 (40.0%) | 63,299 (39.6%) |
| Remainder | 184,075 (60.2%) | 87,374 (60.0%) | 96,701 (60.4%) |
| **Socioeconomic status quintile, no. (%)** |  |  |  |
| 1 (least deprived) | 62,914 (20.6%) | 30,592 (21.0%) | 32,322 (20.2%) |
| 2 to 4 | 185,383 (60.6%) | 87,412 (60.0%) | 97,971 (61.2%) |
| 5 (most deprived) | 57,416 (18.8%) | 27,709 (19.0%) | 29,707 (18.6%) |
| **Other, no. (%)** |  |  |  |
| Never Smoker | 204,740 (67.0%) | 89,362 (61.3%) | 115,378 (72.1%) |
| Previous Smoker | 77,739 (25.4%) | 43,774 (30.0%) | 33,965 (21.2%) |
| Current Smoker | 23,234 (7.6%) | 12,577 (8.6%) | 10,657 (6.7%) |
| Diabetes | 27,428 (9.0%) | 17,217 (11.8%) | 10,211 (6.4%) |
| Depression | 29,685 (9.7%) | 11,295 (7.8%) | 18,390 (11.5%) |
| Cushing's syndrome | 128 (0.0%) | 33 (0.0%) | 95 (0.1%) |
| Hypothyroidism | 22,287 (7.3%) | 4,659 (3.2%) | 17,628 (11.0%) |
| Polycystic ovary syndrome (PCOS) | 601 (0.2%) | 0 (0.0%) | 599 (0.4%) |
| Weight-gain medication use | 27,835 (9.1%) | 14,632 (10.0%) | 13,203 (8.3%) |

**ST4: Cohort characteristics.**

**ST5:** Linear regression. BMI ∼ age + sex + pc’s 1-10 + clinical covariates + Metabolically unhealthy pPS * physical_activity.

| term | Beta | SE | t-statistic | p-value | q-value (BH-FDR) |
| --- | --- | --- | --- | --- | --- |
| (Intercept) | 26.52712199 | 0.076266781 | 347.8201339 | 0 | .. |
| age | -0.008081902 | 0.001042156 | -7.754982129 | 8.86263E-15 | .. |
| sex | 0.641437164 | 0.016587635 | 38.66959731 | 0 | .. |
| harmonized_PRS_1 | 0.28705241 | 0.004932002 | 58.20200071 | 0 | .. |
| pc1 | 0.002657023 | 0.000163744 | 16.22666897 | 3.45912E-59 | .. |
| pc2 | 0.010915479 | 0.000311811 | 35.00670675 | 6.0668E-268 | .. |
| pc3 | 0.000723283 | 0.000578052 | 1.251243234 | 0.210846707 | .. |
| pc4 | 0.002532719 | 0.000776721 | 3.260783371 | 0.00111117 | .. |
| pc5 | 0.013841133 | 0.001055374 | 13.11491029 | 2.77202E-39 | .. |
| pc6 | 0.009168993 | 0.001611419 | 5.69001354 | 1.27145E-08 | .. |
| pc7 | 0.004795687 | 0.00162676 | 2.947999419 | 0.003198618 | .. |
| pc8 | 0.006947357 | 0.001674947 | 4.147806168 | 3.35767E-05 | .. |
| pc9 | -0.01403205 | 0.001804252 | -7.777210536 | 7.43706E-15 | .. |
| pc10 | 0.007267266 | 0.002018714 | 3.599947556 | 0.000318332 | .. |
| physical_activity | -0.695074461 | 0.017369044 | -40.01800313 | 0 | .. |
| sedentary | -1.608715144 | 0.025723739 | -62.53815271 | 0 | .. |
| diet | -0.491869155 | 0.017085935 | -28.78795672 | 5.3247E-182 | .. |
| sleep | -0.477106611 | 0.023927411 | -19.93975109 | 2.09482E-88 | .. |
| higher_ed | -0.705420927 | 0.016748993 | -42.11721371 | 0 | .. |
| TDI_group | 0.35084677 | 0.013101373 | 26.77938881 | 8.5584E-158 | .. |
| depression | 0.655563123 | 0.027480244 | 23.85579708 | 1.1497E-125 | .. |
| hypothyroidism | 0.862741407 | 0.031153004 | 27.69368235 | 1.3465E-168 | .. |
| PCOS | 2.812976141 | 0.180099363 | 15.6190233 | 5.67427E-55 | .. |
| weightgainMeds | 0.90905442 | 0.02926467 | 31.06320371 | 1.6205E-211 | .. |
| smoke_statusFormer | 1.59459808 | 0.033381989 | 47.76821703 | 0 | .. |
| smoke_statusNever | 1.007812894 | 0.031187759 | 32.31437339 | 1.0717E-228 | .. |
| t2dm | 3.490712153 | 0.029604842 | 117.9101763 | 0 | .. |
| cushing | 2.164918152 | 0.389227028 | 5.562096142 | 2.66775E-08 | .. |
| harmonized_PRS_1:physical_activity | -0.049910012 | 0.008811606 | -5.664121976 | 1.47911E-08 | 1.47911E-07 |

**ST6:** Linear regression. BMI ∼ age + sex + pc’s 1-10 + clinical covariates + Strong beta cell pPS * physical_activity.

| term | Beta | SE | t-statistic | p-value | q-value (BH-FDR) |
| --- | --- | --- | --- | --- | --- |
| (Intercept) | 26.58403617 | 0.076699744 | 346.5987585 | 0 | .. |
| age | -0.008880419 | 0.001048065 | -8.473156523 | 2.39868E-17 | .. |
| sex | 0.64187242 | 0.016682738 | 38.47524317 | 0 | .. |
| harmonized PRS 2 | 0.302851873 | 0.010999673 | 27.53280714 | 1.1381E-166 | .. |
| pc1 | 0.002614663 | 0.000164682 | 15.8770886 | 9.6228E-57 | .. |
| pc2 | 0.010914595 | 0.000313598 | 34.80437317 | 6.9266E-265 | .. |
| pc3 | 0.000581263 | 0.000581364 | 0.999827611 | 0.317394733 | .. |
| pc4 | 0.00247424 | 0.000781174 | 3.167334386 | 0.001538584 | .. |
| pc5 | 0.013856276 | 0.001061425 | 13.05440364 | 6.1436E-39 | .. |
| pc6 | 0.009131686 | 0.001620639 | 5.634618978 | 1.75599E-08 | .. |
| pc7 | 0.004537385 | 0.001636077 | 2.773332012 | 0.005548884 | .. |
| pc8 | 0.006997019 | 0.001684548 | 4.153648464 | 3.27304E-05 | .. |
| pc9 | -0.014116127 | 0.00181459 | -7.779236707 | 7.31895E-15 | .. |
| pc10 | 0.00710876 | 0.002030286 | 3.501358838 | 0.000462958 | .. |
| physical activity | -0.69759876 | 0.017468565 | -39.93451954 | 0 | .. |
| sedentary | -1.634476397 | 0.025867476 | -63.18654449 | 0 | .. |
| diet | -0.486711165 | 0.017183629 | -28.32411932 | 2.9539E-176 | .. |
| sleep | -0.475951918 | 0.024064579 | -19.77811104 | 5.21132E-87 | .. |
| higher ed | -0.721931646 | 0.016842932 | -42.86258819 | 0 | .. |
| TDI group | 0.353313002 | 0.013176405 | 26.81406673 | 3.3823E-158 | .. |
| depression | 0.655484042 | 0.027637885 | 23.71686693 | 3.1308E-124 | .. |
| hypothyroidism | 0.877979183 | 0.031330684 | 28.02298209 | 1.4128E-172 | .. |
| PCOS | 2.85800751 | 0.181130215 | 15.77874522 | 4.58599E-56 | .. |
| weightgainMeds | 0.923203429 | 0.029431628 | 31.36773198 | 1.2307E-215 | .. |
| smoke_statusFormer | 1.579706834 | 0.033572741 | 47.05325726 | 0 | .. |
| smoke_statusNever | 0.985341217 | 0.031364718 | 31.41559329 | 2.7485E-216 | .. |
| t2dm | 3.582985408 | 0.029764277 | 120.3787129 | 0 | .. |
| cushing | 2.130802376 | 0.391455042 | 5.443287593 | 5.23462E-08 | .. |
| harmonized PRS 2:physical activity | -0.100806154 | 0.019702318 | -5.116461666 | 3.11508E-07 | 1.55754E-06 |

**ST7:** Linear regression. BMI ∼ age + sex + pc’s 1-10 + clinical covariates + Hypoinsulin 1 pPS * physical_activity.

| term | Beta | SE | t-statistic | p-value | q-value (BH-FDR) |
| --- | --- | --- | --- | --- | --- |
| (Intercept) | 26.55600626 | 0.076657986 | 346.4219152 | 0 | .. |
| age | -0.008455695 | 0.001047513 | -8.072163971 | 6.93106E-16 | .. |
| sex | 0.641507232 | 0.01667308 | 38.47562777 | 0 | .. |
| harmonized PRS 3 | 0.289376899 | 0.009231959 | 31.34512295 | 2.4967E-215 | .. |
| pc1 | 0.002622858 | 0.000164587 | 15.93600064 | 3.75899E-57 | .. |
| pc2 | 0.010902011 | 0.000313417 | 34.78438275 | 1.3856E-264 | .. |
| pc3 | 0.000680229 | 0.000581029 | 1.170732412 | 0.241707265 | .. |
| pc4 | 0.002453843 | 0.00078072 | 3.143051996 | 0.001672121 | .. |
| pc5 | 0.013906013 | 0.00106081 | 13.10886366 | 3.00197E-39 | .. |
| pc6 | 0.009024085 | 0.001619699 | 5.571456886 | 2.5283E-08 | .. |
| pc7 | 0.004433564 | 0.001635128 | 2.711448302 | 0.006699373 | .. |
| pc8 | 0.006940192 | 0.001683572 | 4.122302543 | 3.75203E-05 | .. |
| pc9 | -0.014026595 | 0.001813544 | -7.734357672 | 1.04241E-14 | .. |
| pc10 | 0.007186278 | 0.002029115 | 3.541582444 | 0.000397794 | .. |
| physical activity | -0.700663023 | 0.017458671 | -40.13266583 | 0 | .. |
| sedentary | -1.634320956 | 0.025852284 | -63.21766218 | 0 | .. |
| diet | -0.488627625 | 0.017173856 | -28.45182888 | 7.9085E-178 | .. |
| sleep | -0.474192614 | 0.024050738 | -19.71634373 | 1.76744E-86 | .. |
| higher ed | -0.716577958 | 0.016834181 | -42.56684428 | 0 | .. |
| TDI group | 0.352263306 | 0.013168997 | 26.74944175 | 1.9061E-157 | .. |
| depression | 0.656794435 | 0.027621751 | 23.77816067 | 7.3038E-125 | .. |
| hypothyroidism | 0.876759635 | 0.031312682 | 28.00014488 | 2.6763E-172 | .. |
| PCOS | 2.863760596 | 0.181024988 | 15.81969774 | 2.39635E-56 | .. |
| weightgainMeds | 0.919699517 | 0.029414724 | 31.26663738 | 2.8982E-214 | .. |
| smoke_statusFormer | 1.585525168 | 0.033553447 | 47.25371954 | 0 | .. |
| smoke_statusNever | 0.994260841 | 0.031347557 | 31.71733048 | 2.0504E-220 | .. |
| t2dm | 3.547954602 | 0.029740564 | 119.2968147 | 0 | .. |
| cushing | 2.155218825 | 0.391228429 | 5.508850241 | 3.61475E-08 | .. |
| harmonized PRS 3:physical activity | -0.05765911 | 0.016550824 | -3.483760678 | 0.000494491 | 0.000706415 |

**ST8:**
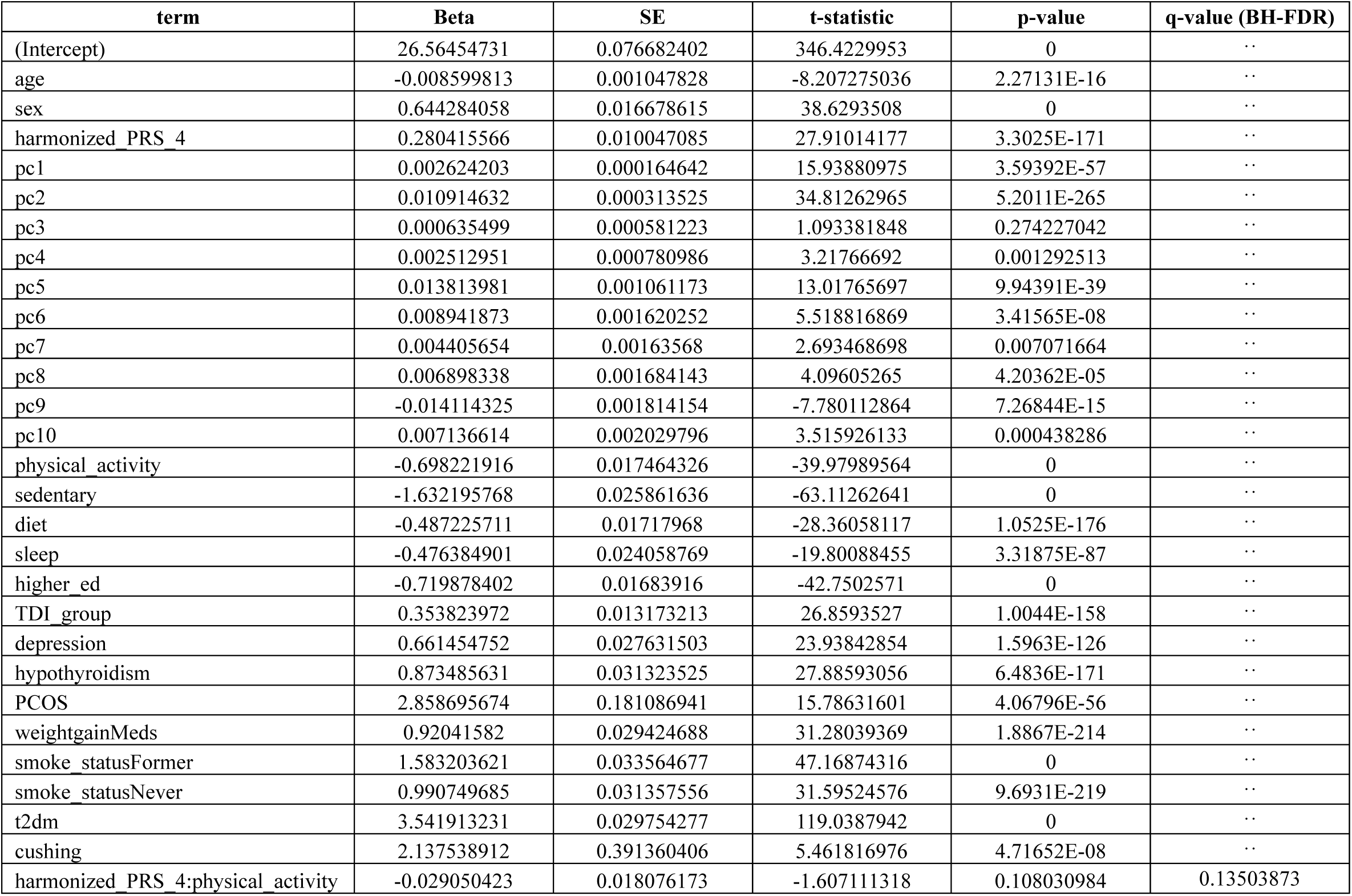
Linear regression. BMI ∼ age + sex + pc’s 1-10 + clinical covariates + Hypoinsulin 2 pPS * physical_activity.

**ST9:** Linear regression. BMI ∼ age + sex + pc’s 1-10 + clinical covariates + Proinsulin pPS * physical_activity.

| term | Beta | SE | t-statistic | p-value | q-value (BH-FDR) |
| --- | --- | --- | --- | --- | --- |
| (Intercept) | 26.57648329 | 0.076802095 | 346.0385213 | 0 | .. |
| age | -0.008764567 | 0.001049466 | -8.351457022 | 6.77024E-17 | .. |
| sex | 0.64257967 | 0.016704982 | 38.46634946 | 0 | .. |
| harmonized PRS 6 | 0.27703277 | 0.03338221 | 8.298814548 | 1.05576E-16 | .. |
| pc1 | 0.002614974 | 0.000164901 | 15.8578467 | 1.30711E-56 | .. |
| pc2 | 0.01091843 | 0.000314017 | 34.77022972 | 2.2633E-264 | .. |
| pc3 | 0.000617801 | 0.000582137 | 1.061264328 | 0.288570631 | .. |
| pc4 | 0.002475381 | 0.000782213 | 3.164587601 | 0.001553181 | .. |
| pc5 | 0.013818366 | 0.001062836 | 13.00141095 | 1.22979E-38 | .. |
| pc6 | 0.009090373 | 0.001622797 | 5.601670586 | 2.12477E-08 | .. |
| pc7 | 0.004466952 | 0.001638263 | 2.726638372 | 0.006398677 | .. |
| pc8 | 0.006962831 | 0.001686794 | 4.127849234 | 3.66269E-05 | .. |
| pc9 | -0.014118502 | 0.001817006 | -7.770200952 | 7.86033E-15 | .. |
| pc10 | 0.007133609 | 0.002032994 | 3.508917301 | 0.000449999 | .. |
| physical activity | -0.69765935 | 0.017492029 | -39.88441547 | 0 | .. |
| sedentary | -1.637534134 | 0.025902121 | -63.22007815 | 0 | .. |
| diet | -0.486280563 | 0.017206855 | -28.26086321 | 1.7643E-175 | .. |
| sleep | -0.476903203 | 0.024096653 | -19.79126357 | 4.016E-87 | .. |
| higher ed | -0.722696733 | 0.016865362 | -42.85094666 | 0 | .. |
| TDI group | 0.354706462 | 0.013193933 | 26.88405832 | 5.1746E-159 | .. |
| depression | 0.65759829 | 0.027674703 | 23.76171085 | 1.0798E-124 | .. |
| hypothyroidism | 0.876950328 | 0.031372913 | 27.9524673 | 1.0141E-171 | .. |
| PCOS | 2.869296462 | 0.181371657 | 15.81998266 | 2.38554E-56 | .. |
| weightgainMeds | 0.924062303 | 0.029470943 | 31.35502962 | 1.8314E-215 | .. |
| smoke_statusFormer | 1.580789657 | 0.033617418 | 47.02293549 | 0 | .. |
| smoke_statusNever | 0.985938853 | 0.031406504 | 31.39282396 | 5.6098E-216 | .. |
| t2dm | 3.561541429 | 0.029795075 | 119.5345696 | 0 | .. |
| cushing | 2.152574612 | 0.391976569 | 5.491590018 | 3.98647E-08 | .. |
| harmonized PRS 6:physical activity | -0.018027546 | 0.059957209 | -0.300673538 | 0.763663653 | 0.763663653 |

**ST10:** Linear regression. BMI ∼ age + sex + pc’s 1-10 + clinical covariates + Immune dysregulation pPS * physical_activity.

| term | Beta | SE | t-statistic | p-value | q-value (BH-FDR) |
| --- | --- | --- | --- | --- | --- |
| (Intercept) | 26.57369139 | 0.076774905 | 346.1247037 | 0 | .. |
| age | -0.008762596 | 0.001049086 | -8.352599347 | 6.70508E-17 | .. |
| sex | 0.643832564 | 0.016698982 | 38.55519902 | 0 | .. |
| harmonized PRS 7 | 0.285856217 | 0.018531645 | 15.42530207 | 1.15922E-53 | .. |
| pc1 | 0.002621405 | 0.000164843 | 15.90247757 | 6.42026E-57 | .. |
| pc2 | 0.010914698 | 0.000313904 | 34.77077889 | 2.2207E-264 | .. |
| pc3 | 0.000591209 | 0.000581935 | 1.015937431 | 0.309659987 | .. |
| pc4 | 0.002475606 | 0.000781935 | 3.165998686 | 0.001545667 | .. |
| pc5 | 0.013827513 | 0.001062458 | 13.01464393 | 1.03438E-38 | .. |
| pc6 | 0.009095368 | 0.001622219 | 5.606746414 | 2.06343E-08 | .. |
| pc7 | 0.004468713 | 0.001637673 | 2.728696547 | 0.006358882 | .. |
| pc8 | 0.006992671 | 0.001686196 | 4.147008475 | 3.36938E-05 | .. |
| pc9 | -0.014147722 | 0.001816362 | -7.789045258 | 6.77276E-15 | .. |
| pc10 | 0.007066965 | 0.002032271 | 3.477374342 | 0.000506421 | .. |
| physical activity | -0.697890391 | 0.017485625 | -39.91223556 | 0 | .. |
| sedentary | -1.638710703 | 0.025892105 | -63.28997617 | 0 | .. |
| diet | -0.485009853 | 0.017200323 | -28.1977174 | 1.0464E-174 | .. |
| sleep | -0.477179508 | 0.024088111 | -19.80975201 | 2.7836E-87 | .. |
| higher ed | -0.721780082 | 0.016859514 | -42.81144059 | 0 | .. |
| TDI group | 0.354130581 | 0.013189215 | 26.85001211 | 1.2905E-158 | .. |
| depression | 0.656798283 | 0.027664814 | 23.74128655 | 1.7539E-124 | .. |
| hypothyroidism | 0.885205177 | 0.031363271 | 28.22426204 | 4.9534E-175 | .. |
| PCOS | 2.865876503 | 0.181306864 | 15.80677335 | 2.94163E-56 | .. |
| weightgainMeds | 0.928659623 | 0.029461574 | 31.52104588 | 1.0026E-217 | .. |
| smoke_statusFormer | 1.584092699 | 0.03360583 | 47.1374377 | 0 | .. |
| smoke_statusNever | 0.987947093 | 0.03139569 | 31.46760285 | 5.3762E-217 | .. |
| t2dm | 3.560167355 | 0.029784554 | 119.5306585 | 0 | .. |
| cushing | 2.132691232 | 0.391837462 | 5.442795652 | 5.2491E-08 | .. |
| harmonized PRS 7:physical activity | -0.046295204 | 0.033212474 | -1.393910111 | 0.163345764 | 0.181495293 |

**ST11:** Linear regression. BMI ∼ age + sex + pc’s 1-10 + clinical covariates + Hyperinsulin 1 pPS * physical_activity.

| term | Beta | SE | t-statistic | p-value | q-value (BH-FDR) |
| --- | --- | --- | --- | --- | --- |
| (Intercept) | 26.57134881 | 0.076645342 | 346.6792374 | 0 | .. |
| age | -0.008694701 | 0.001047326 | -8.301807476 | 1.0295E-16 | .. |
| sex | 0.641817121 | 0.016671085 | 38.49882045 | 0 | .. |
| harmonized PRS 8 | 0.289714127 | 0.008946524 | 32.38287177 | 1.1752E-229 | .. |
| pc1 | 0.002611425 | 0.000164565 | 15.86868773 | 1.1E-56 | .. |
| pc2 | 0.010934911 | 0.000313376 | 34.89391932 | 3.0868E-266 | .. |
| pc3 | 0.00061988 | 0.000580951 | 1.067010074 | 0.28596813 | .. |
| pc4 | 0.002500829 | 0.000780618 | 3.20365169 | 0.001357104 | .. |
| pc5 | 0.013882993 | 0.00106067 | 13.08888615 | 3.90522E-39 | .. |
| pc6 | 0.009133016 | 0.001619488 | 5.639447058 | 1.70747E-08 | .. |
| pc7 | 0.004405867 | 0.001634921 | 2.694850608 | 0.007042404 | .. |
| pc8 | 0.0069787 | 0.00168335 | 4.145720993 | 3.38837E-05 | .. |
| pc9 | -0.014132328 | 0.001813301 | -7.793700497 | 6.52778E-15 | .. |
| pc10 | 0.00729937 | 0.002028848 | 3.597791312 | 0.000320982 | .. |
| physical activity | -0.700037294 | 0.017456231 | -40.10243171 | 0 | .. |
| sedentary | -1.633449223 | 0.025849043 | -63.19186535 | 0 | .. |
| diet | -0.491621186 | 0.017172244 | -28.62882598 | 5.0978E-180 | .. |
| sleep | -0.474445015 | 0.024047545 | -19.72945755 | 1.36419E-86 | .. |
| higher ed | -0.715798491 | 0.016832134 | -42.52571186 | 0 | .. |
| TDI group | 0.351289919 | 0.013167266 | 26.67903169 | 1.2482E-156 | .. |
| depression | 0.659393291 | 0.027618368 | 23.87517185 | 7.2411E-126 | .. |
| hypothyroidism | 0.874126826 | 0.031308645 | 27.91966302 | 2.5326E-171 | .. |
| PCOS | 2.851340824 | 0.181001833 | 15.7531047 | 6.87943E-56 | .. |
| weightgainMeds | 0.923438946 | 0.029410682 | 31.39808029 | 4.7581E-216 | .. |
| smoke_statusFormer | 1.584143915 | 0.033548953 | 47.21887797 | 0 | .. |
| smoke_statusNever | 0.993624981 | 0.031343203 | 31.70144971 | 3.3887E-220 | .. |
| t2dm | 3.566849911 | 0.029734713 | 119.9557531 | 0 | .. |
| cushing | 2.152174684 | 0.391176494 | 5.501799609 | 3.76234E-08 | .. |
| harmonized PRS 8:physical activity | -0.059007135 | 0.016076466 | -3.670404596 | 0.000242208 | 0.00040368 |

**ST12:** Linear regression. BMI ∼ age + sex + pc’s 1-10 + clinical covariates + Hyperinsulin 2 pPS * physical_activity.

| term | Beta | SE | t-statistic | p-value | q-value (BH-FDR) |
| --- | --- | --- | --- | --- | --- |
| (Intercept) | 26.56328091 | 0.076613498 | 346.7180282 | 0 | .. |
| age | -0.008584941 | 0.001046887 | -8.20044747 | 2.40407E-16 | .. |
| sex | 0.64162462 | 0.016663937 | 38.50378226 | 0 | .. |
| harmonized PRS 9 | 0.288480854 | 0.008185096 | 35.24464993 | 1.4616E-271 | .. |
| pc1 | 0.002611143 | 0.000164495 | 15.87367303 | 1.01607E-56 | .. |
| pc2 | 0.010925027 | 0.000313242 | 34.87723128 | 5.5151E-266 | .. |
| pc3 | 0.000585682 | 0.000580704 | 1.008572165 | 0.313180661 | .. |
| pc4 | 0.002533314 | 0.000780292 | 3.246621711 | 0.001167961 | .. |
| pc5 | 0.013856578 | 0.00106022 | 13.06953523 | 5.03659E-39 | .. |
| pc6 | 0.00918985 | 0.001618806 | 5.676930478 | 1.37257E-08 | .. |
| pc7 | 0.004353807 | 0.001634228 | 2.664137393 | 0.007719008 | .. |
| pc8 | 0.006980314 | 0.001682635 | 4.148441239 | 3.34837E-05 | .. |
| pc9 | -0.014084748 | 0.001812533 | -7.770753027 | 7.82615E-15 | .. |
| pc10 | 0.007012123 | 0.002027983 | 3.457682687 | 0.000544917 | .. |
| physical activity | -0.70099245 | 0.017448947 | -40.17391081 | 0 | .. |
| sedentary | -1.631948036 | 0.025838161 | -63.16037908 | 0 | .. |
| diet | -0.48814372 | 0.01716423 | -28.43959294 | 1.1195E-177 | .. |
| sleep | -0.473649765 | 0.024037406 | -19.70469586 | 2.22424E-86 | .. |
| higher ed | -0.715761523 | 0.016824774 | -42.54211835 | 0 | .. |
| TDI group | 0.352422093 | 0.013161504 | 26.77673327 | 9.1885E-158 | .. |
| depression | 0.657434595 | 0.027606411 | 23.81456271 | 3.0717E-125 | .. |
| hypothyroidism | 0.869724289 | 0.031296108 | 27.79017444 | 9.2909E-170 | .. |
| PCOS | 2.865535522 | 0.180924805 | 15.83826782 | 1.78439E-56 | .. |
| weightgainMeds | 0.919153489 | 0.029398543 | 31.26527406 | 3.0241E-214 | .. |
| smoke_statusFormer | 1.582497609 | 0.033534867 | 47.18961915 | 0 | .. |
| smoke_statusNever | 0.991547371 | 0.031329637 | 31.64886242 | 1.7857E-219 | .. |
| t2dm | 3.570147763 | 0.02972265 | 120.1153934 | 0 | .. |
| cushing | 2.106988732 | 0.391011894 | 5.388554072 | 7.10791E-08 | .. |
| harmonized PRS 9:physical activity | -0.054964524 | 0.014700279 | -3.739012346 | 0.000184778 | 0.000369557 |

**ST13:**
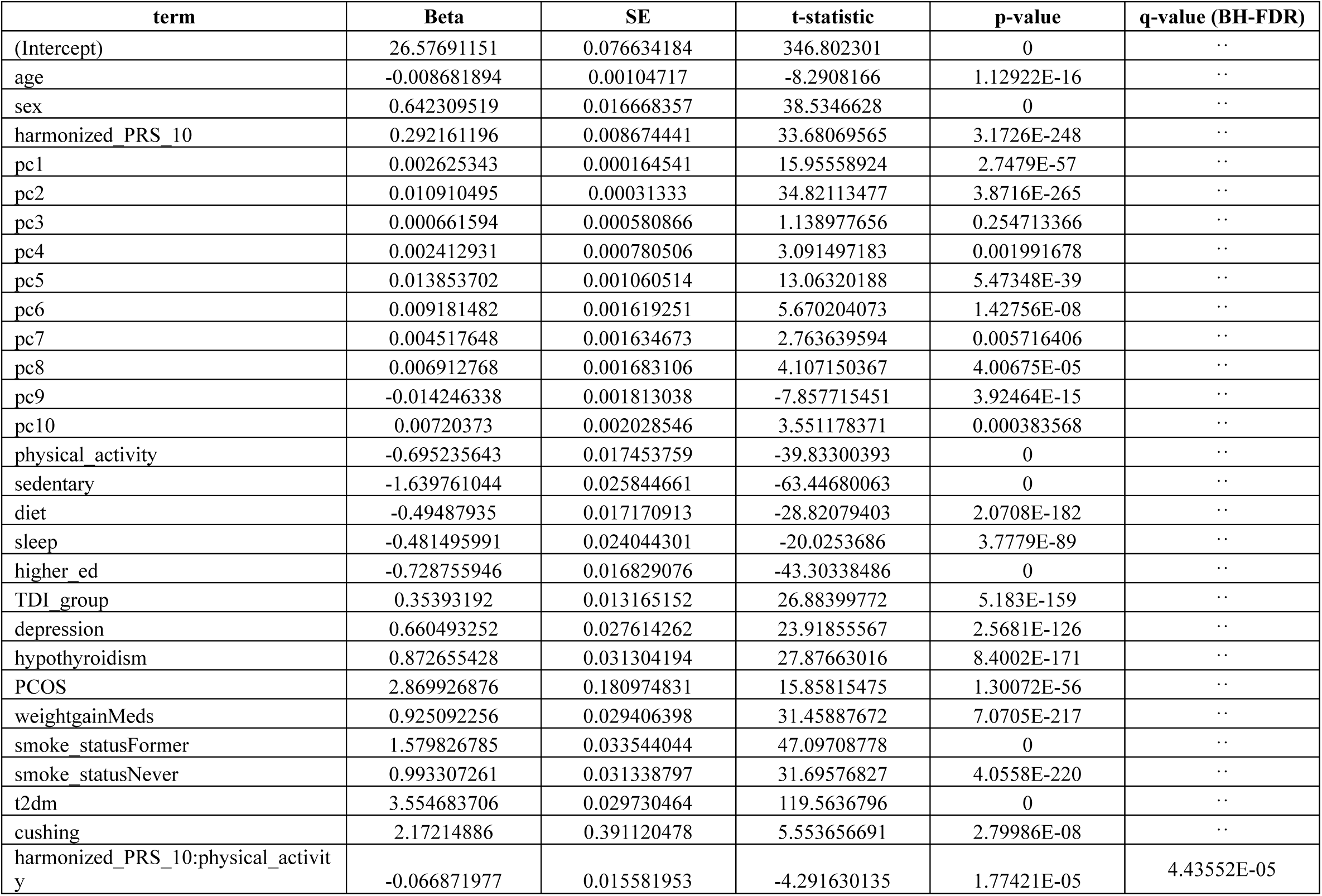
Linear regression. BMI ∼ age + sex + pc’s 1-10 + clinical covariates + Hypothalamic dysregulation pPS * physical_activity.

| term | Beta | SE | t-statistic | p-value | q-value (BH-FDR) |
| --- | --- | --- | --- | --- | --- |
| (Intercept) | 26.57691151 | 0.076634184 | 346.802301 | 0 | .. |
| age | -0.008681894 | 0.00104717 | -8.2908166 | 1.12922E-16 | .. |
| sex | 0.642309519 | 0.016668357 | 38.5346628 | 0 | .. |
| harmonized PRS 10 | 0.292161196 | 0.008674441 | 33.68069565 | 3.1726E-248 | .. |
| pc1 | 0.002625343 | 0.000164541 | 15.95558924 | 2.7479E-57 | .. |
| pc2 | 0.010910495 | 0.00031333 | 34.82113477 | 3.8716E-265 | .. |
| pc3 | 0.000661594 | 0.000580866 | 1.138977656 | 0.254713366 | .. |
| pc4 | 0.002412931 | 0.000780506 | 3.091497183 | 0.001991678 | .. |
| pc5 | 0.013853702 | 0.001060514 | 13.06320188 | 5.47348E-39 | .. |
| pc6 | 0.009181482 | 0.001619251 | 5.670204073 | 1.42756E-08 | .. |
| pc7 | 0.004517648 | 0.001634673 | 2.763639594 | 0.005716406 | .. |
| pc8 | 0.006912768 | 0.001683106 | 4.107150367 | 4.00675E-05 | .. |
| pc9 | -0.014246338 | 0.001813038 | -7.857715451 | 3.92464E-15 | .. |
| pc10 | 0.00720373 | 0.002028546 | 3.551178371 | 0.000383568 | .. |
| physical activity | -0.695235643 | 0.017453759 | -39.83300393 | 0 | .. |
| sedentary | -1.639761044 | 0.025844661 | -63.44680063 | 0 | .. |
| diet | -0.49487935 | 0.017170913 | -28.82079403 | 2.0708E-182 | .. |
| sleep | -0.481495991 | 0.024044301 | -20.0253686 | 3.7779E-89 | .. |
| higher ed | -0.728755946 | 0.016829076 | -43.30338486 | 0 | .. |
| TDI group | 0.35393192 | 0.013165152 | 26.88399772 | 5.183E-159 | .. |
| depression | 0.660493252 | 0.027614262 | 23.91855567 | 2.5681E-126 | .. |
| hypothyroidism | 0.872655428 | 0.031304194 | 27.87663016 | 8.4002E-171 | .. |
| PCOS | 2.869926876 | 0.180974831 | 15.85815475 | 1.30072E-56 | .. |
| weightgainMeds | 0.925092256 | 0.029406398 | 31.45887672 | 7.0705E-217 | .. |
| smoke_statusFormer | 1.579826785 | 0.033544044 | 47.09708778 | 0 | .. |
| smoke_statusNever | 0.993307261 | 0.031338797 | 31.69576827 | 4.0558E-220 | .. |
| t2dm | 3.554683706 | 0.029730464 | 119.5636796 | 0 | .. |
| cushing | 2.17214886 | 0.391120478 | 5.553656691 | 2.79986E-08 | .. |
| harmonized_PRS_10:physical_activity | -0.066871977 | 0.015581953 | -4.291630135 | 1.77421E-05 | 4.43552E-05 |

**ST14:** Linear regression. BMI ∼ age + sex + pc’s 1-10 + clinical covariates + Metabolically healthy pPS * physical_activity.

| term | Beta | SE | t-statistic | p-value | q-value (BH-FDR) |
| --- | --- | --- | --- | --- | --- |
| (Intercept) | 26.57812929 | 0.07670298 | 346.5071298 | 0 | .. |
| age | -0.008857893 | 0.001048109 | -8.451305917 | 2.893E-17 | .. |
| sex | 0.639314911 | 0.016683914 | 38.31924127 | 0 | .. |
| harmonized PRS 11 | 0.299870931 | 0.011111738 | 26.98686237 | 3.2544E-160 | .. |
| pc1 | 0.002608565 | 0.000164689 | 15.83934995 | 1.75398E-56 | .. |
| pc2 | 0.010928511 | 0.000313612 | 34.8472248 | 1.5647E-265 | .. |
| pc3 | 0.000567166 | 0.00058139 | 0.975534833 | 0.329295804 | .. |
| pc4 | 0.00244267 | 0.000781208 | 3.126785238 | 0.001767455 | .. |
| pc5 | 0.013823461 | 0.001061471 | 13.02292903 | 9.28087E-39 | .. |
| pc6 | 0.009036917 | 0.001620709 | 5.575903049 | 2.46456E-08 | .. |
| pc7 | 0.00450924 | 0.001636148 | 2.75600995 | 0.005851468 | .. |
| pc8 | 0.006857199 | 0.001684629 | 4.070451605 | 4.69339E-05 | .. |
| pc9 | -0.014131075 | 0.00181467 | -7.787132084 | 6.87605E-15 | .. |
| pc10 | 0.007066975 | 0.002030375 | 3.480625002 | 0.000500315 | .. |
| physical activity | -0.696756549 | 0.017469556 | -39.88404373 | 0 | .. |
| sedentary | -1.638998587 | 0.02586798 | -63.36012992 | 0 | .. |
| diet | -0.486566174 | 0.017184371 | -28.3144593 | 3.8819E-176 | .. |
| sleep | -0.476002737 | 0.024065624 | -19.77936392 | 5.08361E-87 | .. |
| higher ed | -0.722534738 | 0.016843667 | -42.89652224 | 0 | .. |
| TDI_group | 0.354243351 | 0.013176937 | 26.88358946 | 5.2401E-159 | .. |
| depression | 0.654559855 | 0.027639124 | 23.68236624 | 7.0914E-124 | .. |
| hypothyroidism | 0.875375682 | 0.031332252 | 27.93848598 | 1.4981E-171 | .. |
| PCOS | 2.877697322 | 0.181138492 | 15.88672452 | 8.25338E-57 | .. |
| weightgainMeds | 0.927728872 | 0.029433368 | 31.51962989 | 1.0482E-217 | .. |
| smoke_statusFormer | 1.582320617 | 0.033574348 | 47.12885662 | 0 | .. |
| smoke_statusNever | 0.98963556 | 0.03136646 | 31.55075758 | 3.9365E-218 | .. |
| t2dm | 3.594369769 | 0.029777624 | 120.7070717 | 0 | .. |
| cushing | 2.135709984 | 0.391472214 | 5.455585116 | 4.88496E-08 | .. |
| harmonized_PRS_11:physical_activity | -0.092184925 | 0.019999575 | -4.609344205 | 4.04104E-06 | 1.34701E-05 |

**ST15:** Linear regression. BMI ∼ age + sex + pc’s 1-10 + clinical covariates + Metabolically unhealthy pPS * physical_activity.

| term | Beta | SE | t-statistic | p-value | q-value (BH-FDR) |
| --- | --- | --- | --- | --- | --- |
| (Intercept) | 26.52694329 | 0.076269136 | 347.8070512 | 0 | .. |
| age | -0.008066967 | 0.00104218 | -7.740470186 | 9.93507E-15 | .. |
| sex | 0.64114722 | 0.016588031 | 38.6511953 | 0 | .. |
| harmonized PRS 1 | 0.276985184 | 0.00434356 | 63.76915694 | 0 | .. |
| pc1 | 0.002659279 | 0.000163749 | 16.23994088 | 2.78694E-59 | .. |
| pc2 | 0.010913889 | 0.000311822 | 35.0004215 | 7.5545E-268 | .. |
| pc3 | 0.000730933 | 0.00057807 | 1.264436436 | 0.206074386 | .. |
| pc4 | 0.002539737 | 0.000776744 | 3.2697223 | 0.00107665 | .. |
| pc5 | 0.013837463 | 0.001055404 | 13.11105761 | 2.91642E-39 | .. |
| pc6 | 0.009120043 | 0.001611447 | 5.659535118 | 1.51919E-08 | .. |
| pc7 | 0.004797209 | 0.001626808 | 2.948846941 | 0.00318986 | .. |
| pc8 | 0.006963249 | 0.001674994 | 4.157179946 | 3.22288E-05 | .. |
| pc9 | -0.014011805 | 0.0018043 | -7.765783372 | 8.13918E-15 | .. |
| pc10 | 0.007269725 | 0.002018775 | 3.601057992 | 0.000316975 | .. |
| physical activity | -0.695100535 | 0.017369562 | -40.01831003 | 0 | .. |
| sedentary | -1.613441384 | 0.025758977 | -62.63608206 | 0 | .. |
| diet | -0.492174234 | 0.017086468 | -28.80491283 | 3.2701E-182 | .. |
| sleep | -0.477203687 | 0.023928178 | -19.94316853 | 1.95665E-88 | .. |
| higher ed | -0.705701102 | 0.016749592 | -42.13243501 | 0 | .. |
| TDI group | 0.351025741 | 0.013101736 | 26.79230773 | 6.0569E-158 | .. |
| depression | 0.655847056 | 0.02748115 | 23.86534208 | 9.1557E-126 | .. |
| hypothyroidism | 0.862832273 | 0.031153916 | 27.69578901 | 1.2702E-168 | .. |
| PCOS | 2.812825034 | 0.180104647 | 15.61772604 | 5.79079E-55 | .. |
| weightgainMeds | 0.908971664 | 0.029265639 | 31.05934834 | 1.8262E-211 | .. |
| smoke_statusFormer | 1.593798085 | 0.033382995 | 47.74281225 | 0 | .. |
| smoke_statusNever | 1.007237012 | 0.031188592 | 32.29504575 | 1.9978E-228 | .. |
| t2dm | 3.490640841 | 0.029605961 | 117.90331 | 0 | .. |
| cushing | 2.173841223 | 0.389234861 | 5.584908863 | 2.34021E-08 | .. |
| harmonized PRS 1:sedentary | -0.048913312 | 0.012896075 | -3.792883644 | 0.000148937 | 0.000496455 |

**ST16:**
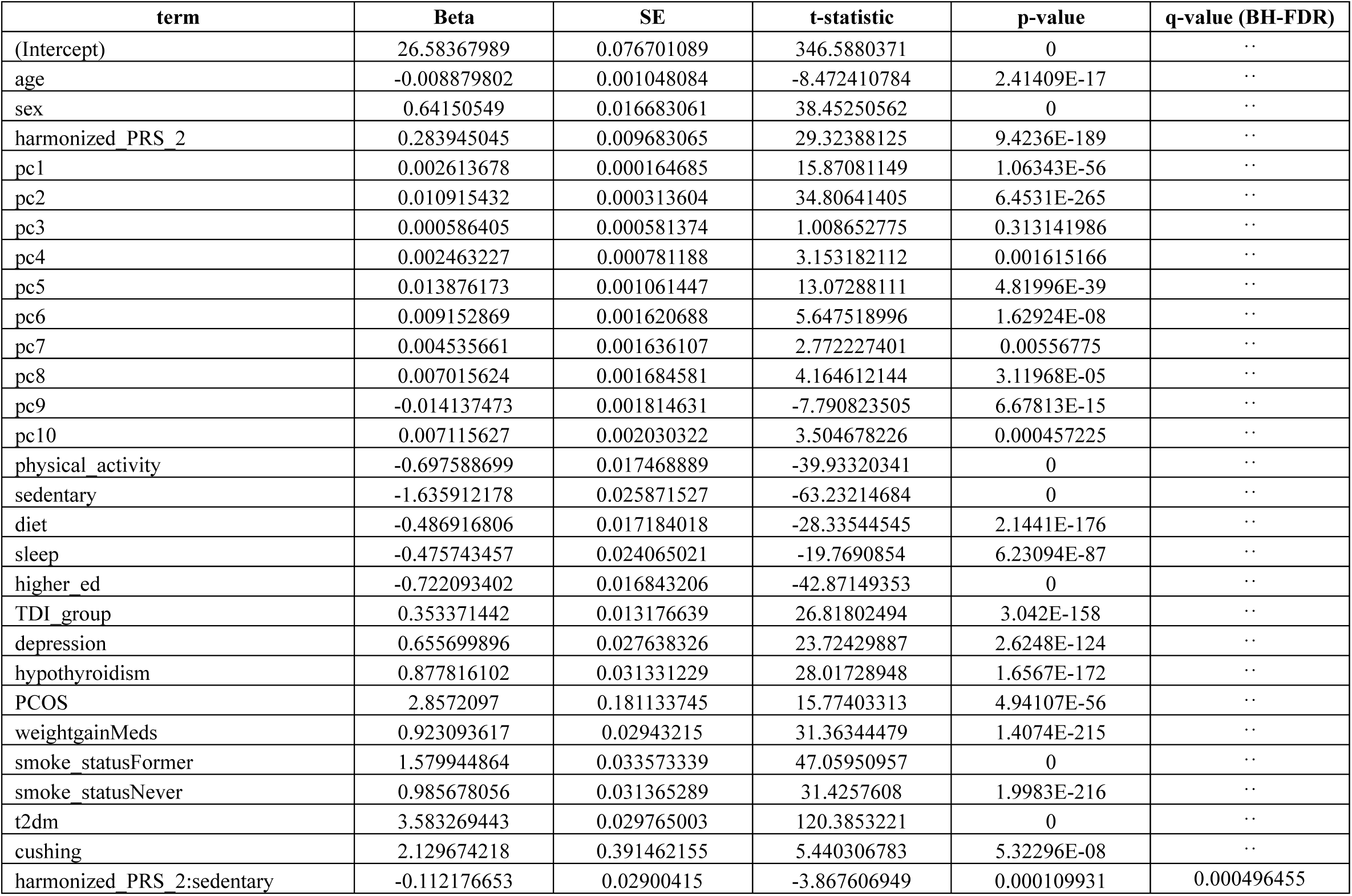
Linear regression. BMI ∼ age + sex + pc’s 1-10 + clinical covariates + Strong beta cell pPS * sedentary.

**ST17:** Linear regression. BMI ∼ age + sex + pc’s 1-10 + clinical covariates + Hypoinsulin 1 pPS * sedentary.

| term | Beta | SE | t-statistic | p-value | q-value (BH-FDR) |
| --- | --- | --- | --- | --- | --- |
| (Intercept) | 26.55530945 | 0.076658733 | 346.4094503 | 0 | .. |
| age | -0.008429822 | 0.001047523 | -8.047386412 | 8.48803E-16 | .. |
| sex | 0.641340523 | 0.01667319 | 38.46537548 | 0 | .. |
| harmonized PRS 3 | 0.277985157 | 0.008126146 | 34.20873447 | 5.4901E-256 | .. |
| pc1 | 0.002626379 | 0.000164589 | 15.95714847 | 2.68017E-57 | .. |
| pc2 | 0.010902031 | 0.00031342 | 34.78407201 | 1.4007E-264 | .. |
| pc3 | 0.000684422 | 0.000581036 | 1.177933449 | 0.238824056 | .. |
| pc4 | 0.002457227 | 0.000780729 | 3.147350729 | 0.00164773 | .. |
| pc5 | 0.013908309 | 0.001060822 | 13.11088181 | 2.92319E-39 | .. |
| pc6 | 0.009014864 | 0.001619714 | 5.565711912 | 2.61302E-08 | .. |
| pc7 | 0.004448805 | 0.001635142 | 2.720744776 | 0.006513871 | .. |
| pc8 | 0.006925603 | 0.001683588 | 4.11359669 | 3.89644E-05 | .. |
| pc9 | -0.014031536 | 0.001813574 | -7.736954064 | 1.02136E-14 | .. |
| pc10 | 0.007170722 | 0.002029129 | 3.533892135 | 0.000409549 | .. |
| physical activity | -0.700964419 | 0.017458732 | -40.14979073 | 0 | .. |
| sedentary | -1.635337949 | 0.025856951 | -63.2455827 | 0 | .. |
| diet | -0.488577697 | 0.017174033 | -28.4486299 | 8.6609E-178 | .. |
| sleep | -0.474134093 | 0.024050977 | -19.71371479 | 1.86159E-86 | .. |
| higher ed | -0.716568849 | 0.016834369 | -42.56582746 | 0 | .. |
| TDI group | 0.352047185 | 0.013169011 | 26.73300171 | 2.9574E-157 | .. |
| depression | 0.656992523 | 0.02762204 | 23.78508336 | 6.1952E-125 | .. |
| hypothyroidism | 0.876999252 | 0.031312865 | 28.0076333 | 2.1706E-172 | .. |
| PCOS | 2.863447383 | 0.181026975 | 15.8177939 | 2.46985E-56 | .. |
| weightgainMeds | 0.919703542 | 0.029415037 | 31.26644165 | 2.9159E-214 | .. |
| smoke_statusFormer | 1.585233725 | 0.033553794 | 47.24454521 | 0 | .. |
| smoke_statusNever | 0.993747086 | 0.031347801 | 31.70069544 | 3.4706E-220 | .. |
| t2dm | 3.547666981 | 0.029741278 | 119.2842813 | 0 | .. |
| cushing | 2.154657594 | 0.391234067 | 5.507336334 | 3.64596E-08 | .. |
| harmonized PRS 3:sedentary | -0.059154241 | 0.024434952 | -2.42088631 | 0.015483297 | 0.030966594 |

**ST18:**
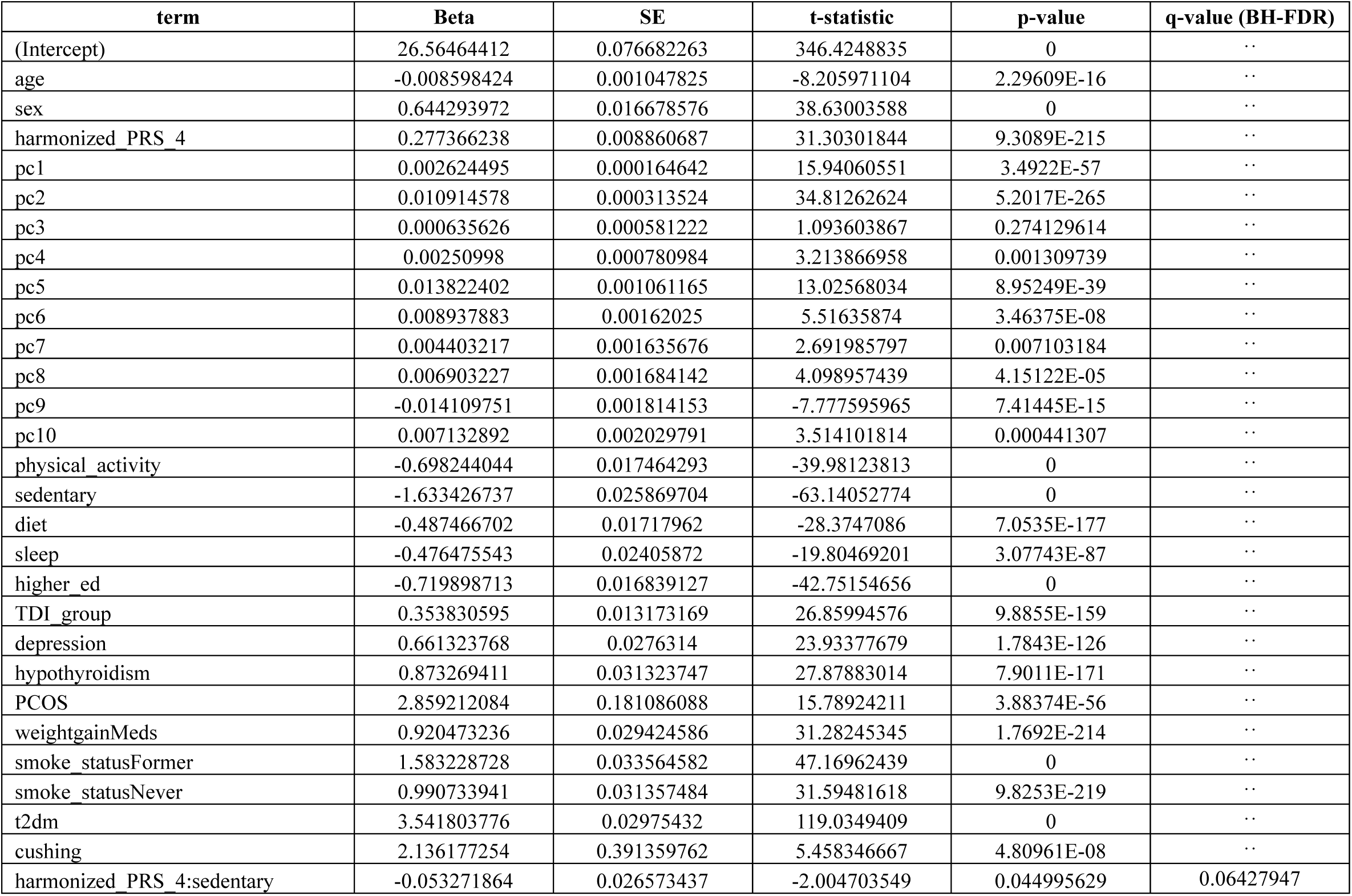
Linear regression. BMI ∼ age + sex + pc’s 1-10 + clinical covariates + Hypoinsulin 2 pPS * sedentary.

| term | Beta | SE | t-statistic | p-value | q-value (BH-FDR) |
| --- | --- | --- | --- | --- | --- |
| (Intercept) | 26.56464412 | 0.076682263 | 346.4248835 | 0 | .. |
| age | -0.008598424 | 0.001047825 | -8.205971104 | 2.29609E-16 | .. |
| sex | 0.644293972 | 0.016678576 | 38.63003588 | 0 | .. |
| harmonized PRS 4 | 0.277366238 | 0.008860687 | 31.30301844 | 9.3089E-215 | .. |
| pc1 | 0.002624495 | 0.000164642 | 15.94060551 | 3.4922E-57 | .. |
| pc2 | 0.010914578 | 0.000313524 | 34.81262624 | 5.2017E-265 | .. |
| pc3 | 0.000635626 | 0.000581222 | 1.093603867 | 0.274129614 | .. |
| pc4 | 0.00250998 | 0.000780984 | 3.213866958 | 0.001309739 | .. |
| pc5 | 0.013822402 | 0.001061165 | 13.02568034 | 8.95249E-39 | .. |
| pc6 | 0.008937883 | 0.00162025 | 5.51635874 | 3.46375E-08 | .. |
| pc7 | 0.004403217 | 0.001635676 | 2.691985797 | 0.007103184 | .. |
| pc8 | 0.006903227 | 0.001684142 | 4.098957439 | 4.15122E-05 | .. |
| pc9 | -0.014109751 | 0.001814153 | -7.777595965 | 7.41445E-15 | .. |
| pc10 | 0.007132892 | 0.002029791 | 3.514101814 | 0.000441307 | .. |
| physical activity | -0.698244044 | 0.017464293 | -39.98123813 | 0 | .. |
| sedentary | -1.633426737 | 0.025869704 | -63.14052774 | 0 | .. |
| diet | -0.487466702 | 0.01717962 | -28.3747086 | 7.0535E-177 | .. |
| sleep | -0.476475543 | 0.02405872 | -19.80469201 | 3.07743E-87 | .. |
| higher ed | -0.719898713 | 0.016839127 | -42.75154656 | 0 | .. |
| TDI group | 0.353830595 | 0.013173169 | 26.85994576 | 9.8855E-159 | .. |
| depression | 0.661323768 | 0.0276314 | 23.93377679 | 1.7843E-126 | .. |
| hypothyroidism | 0.873269411 | 0.031323747 | 27.87883014 | 7.9011E-171 | .. |
| PCOS | 2.859212084 | 0.181086088 | 15.78924211 | 3.88374E-56 | .. |
| weightgainMeds | 0.920473236 | 0.029424586 | 31.28245345 | 1.7692E-214 | .. |
| smoke_statusFormer | 1.583228728 | 0.033564582 | 47.16962439 | 0 | .. |
| smoke_statusNever | 0.990733941 | 0.031357484 | 31.59481618 | 9.8253E-219 | .. |
| t2dm | 3.541803776 | 0.02975432 | 119.0349409 | 0 | .. |
| cushing | 2.136177254 | 0.391359762 | 5.458346667 | 4.80961E-08 | .. |
| harmonized PRS 4:sedentary | -0.053271864 | 0.026573437 | -2.004703549 | 0.044995629 | 0.06427947 |

**ST19:** Linear regression. BMI ∼ age + sex + pc’s 1-10 + clinical covariates + Proinsulin pPS * sedentary.

| term | Beta | SE | t-statistic | p-value | q-value (BH-FDR) |
| --- | --- | --- | --- | --- | --- |
| (Intercept) | 26.57641247 | 0.076802592 | 346.0353588 | 0 | .. |
| age | -0.008763537 | 0.001049468 | -8.350458727 | 6.8277E-17 | .. |
| sex | 0.642573547 | 0.016704985 | 38.46597568 | 0 | .. |
| harmonized PRS 6 | 0.269737203 | 0.029409813 | 9.171673511 | 4.68498E-20 | .. |
| pc1 | 0.002615076 | 0.000164901 | 15.85848109 | 1.29399E-56 | .. |
| pc2 | 0.010918714 | 0.000314016 | 34.77119734 | 2.1887E-264 | .. |
| pc3 | 0.000618159 | 0.000582138 | 1.061877778 | 0.288292014 | .. |
| pc4 | 0.002475012 | 0.000782214 | 3.164111711 | 0.001555723 | .. |
| pc5 | 0.013818526 | 0.001062836 | 13.00156181 | 1.22737E-38 | .. |
| pc6 | 0.009090659 | 0.001622797 | 5.601847661 | 2.12261E-08 | .. |
| pc7 | 0.0044684 | 0.001638255 | 2.727535873 | 0.006381296 | .. |
| pc8 | 0.006961451 | 0.00168679 | 4.127041479 | 3.67557E-05 | .. |
| pc9 | -0.014118234 | 0.001817008 | -7.77004473 | 7.87003E-15 | .. |
| pc10 | 0.007131595 | 0.00203299 | 3.507934777 | 0.000451665 | .. |
| physical activity | -0.697643657 | 0.017491973 | -39.88364495 | 0 | .. |
| sedentary | -1.637436885 | 0.025907323 | -63.2036319 | 0 | .. |
| diet | -0.48629016 | 0.017206818 | -28.26148045 | 1.7338E-175 | .. |
| sleep | -0.476895783 | 0.024096636 | -19.79096896 | 4.03951E-87 | .. |
| higher ed | -0.722687621 | 0.016865467 | -42.85014073 | 0 | .. |
| TDI group | 0.354714413 | 0.013194009 | 26.88450501 | 5.1128E-159 | .. |
| depression | 0.657614499 | 0.027674669 | 23.76232609 | 1.0641E-124 | .. |
| hypothyroidism | 0.876959297 | 0.03137293 | 27.95273831 | 1.0064E-171 | .. |
| PCOS | 2.869270731 | 0.181371655 | 15.81984093 | 2.39091E-56 | .. |
| weightgainMeds | 0.924024475 | 0.029470789 | 31.3539106 | 1.8966E-215 | .. |
| smoke_statusFormer | 1.580799119 | 0.033617447 | 47.0231752 | 0 | .. |
| smoke_statusNever | 0.985935394 | 0.031406519 | 31.39269925 | 5.6317E-216 | .. |
| t2dm | 3.561538298 | 0.029795079 | 119.534446 | 0 | .. |
| cushing | 2.152606894 | 0.391976863 | 5.491668248 | 3.9847E-08 | .. |
| harmonized PRS 6:sedentary | 0.015398024 | 0.088300397 | 0.174382269 | 0.861565163 | 0.861565163 |

**ST20:** Linear regression. BMI ∼ age + sex + pc’s 1-10 + clinical covariates + Immune dysregulation pPS * sedentary.

| term | Beta | SE | t-statistic | p-value | q-value (BH-FDR) |
| --- | --- | --- | --- | --- | --- |
| (Intercept) | 26.57377364 | 0.076774625 | 346.12704 | 0 | .. |
| age | -0.008759858 | 0.001049084 | -8.350008563 | 6.85377E-17 | .. |
| sex | 0.643676424 | 0.016698887 | 38.54606721 | 0 | .. |
| harmonized PRS 7 | 0.282490385 | 0.016324921 | 17.30424179 | 4.70452E-67 | .. |
| pc1 | 0.002621736 | 0.000164842 | 15.90453172 | 6.21328E-57 | .. |
| pc2 | 0.010913747 | 0.000313904 | 34.76780511 | 2.4618E-264 | .. |
| pc3 | 0.00059582 | 0.000581929 | 1.023871052 | 0.305896995 | .. |
| pc4 | 0.002475114 | 0.000781932 | 3.165382851 | 0.001548942 | .. |
| pc5 | 0.013825616 | 0.001062455 | 13.01289716 | 1.05829E-38 | .. |
| pc6 | 0.009101352 | 0.001622216 | 5.610442565 | 2.01984E-08 | .. |
| pc7 | 0.004473831 | 0.001637665 | 2.731835412 | 0.006298621 | .. |
| pc8 | 0.007005242 | 0.001686188 | 4.154485332 | 3.26109E-05 | .. |
| pc9 | -0.014141919 | 0.001816355 | -7.785877437 | 6.94462E-15 | .. |
| pc10 | 0.007067556 | 0.002032262 | 3.477680397 | 0.000505843 | .. |
| physical activity | -0.697798443 | 0.017485552 | -39.9071445 | 0 | .. |
| sedentary | -1.639010961 | 0.025892267 | -63.30117698 | 0 | .. |
| diet | -0.484986676 | 0.01720027 | -28.19645751 | 1.0842E-174 | .. |
| sleep | -0.477330024 | 0.024088017 | -19.81607787 | 2.45532E-87 | .. |
| higher ed | -0.721902716 | 0.016859435 | -42.81891546 | 0 | .. |
| TDI group | 0.354076363 | 0.013189209 | 26.84591405 | 1.4404E-158 | .. |
| depression | 0.65691214 | 0.027664625 | 23.74556496 | 1.5845E-124 | .. |
| hypothyroidism | 0.885327594 | 0.031363216 | 28.22821486 | 4.4311E-175 | .. |
| PCOS | 2.864146729 | 0.181307373 | 15.79718836 | 3.42433E-56 | .. |
| weightgainMeds | 0.9284321 | 0.029461522 | 31.51337873 | 1.276E-217 | .. |
| smoke_statusFormer | 1.584169569 | 0.033605713 | 47.1398888 | 0 | .. |
| smoke_statusNever | 0.98806772 | 0.031395509 | 31.47162594 | 4.7382E-217 | .. |
| t2dm | 3.56031789 | 0.029784485 | 119.5359895 | 0 | .. |
| cushing | 2.134361508 | 0.391837525 | 5.447057449 | 5.12493E-08 | .. |
| harmonized PRS 7:sedentary | -0.098201523 | 0.048672588 | -2.017594013 | 0.043634433 | 0.06427947 |

**ST21:** Linear regression. BMI ∼ age + sex + pc’s 1-10 + clinical covariates + Hyperinsulin 1 pPS * sedentary.

| term | Beta | SE | t-statistic | p-value | q-value (BH-FDR) |
| --- | --- | --- | --- | --- | --- |
| (Intercept) | 26.57127792 | 0.076647103 | 346.670349 | 0 | .. |
| age | -0.008683863 | 0.001047344 | -8.291319378 | 1.12445E-16 | .. |
| sex | 0.641480047 | 0.016671178 | 38.47838741 | 0 | .. |
| harmonized PRS 8 | 0.273560835 | 0.007881973 | 34.70715292 | 2.011E-263 | .. |
| pc1 | 0.002610651 | 0.000164568 | 15.86366404 | 1.19156E-56 | .. |
| pc2 | 0.01093555 | 0.000313383 | 34.89516694 | 2.9557E-266 | .. |
| pc3 | 0.00062411 | 0.000580962 | 1.074270612 | 0.28270226 | .. |
| pc4 | 0.00249815 | 0.000780638 | 3.200138475 | 0.001373756 | .. |
| pc5 | 0.013885085 | 0.001060693 | 13.09058348 | 3.81897E-39 | .. |
| pc6 | 0.009124312 | 0.001619521 | 5.633955752 | 1.76276E-08 | .. |
| pc7 | 0.00442356 | 0.001634949 | 2.705626202 | 0.006817952 | .. |
| pc8 | 0.006974999 | 0.001683385 | 4.143436063 | 3.42232E-05 | .. |
| pc9 | -0.014129092 | 0.00181334 | -7.791750828 | 6.6293E-15 | .. |
| pc10 | 0.00729688 | 0.002028891 | 3.59648672 | 0.000322595 | .. |
| physical activity | -0.700204237 | 0.017456566 | -40.11122386 | 0 | .. |
| sedentary | -1.633723303 | 0.025855161 | -63.18751331 | 0 | .. |
| diet | -0.49151951 | 0.017172604 | -28.6223056 | 6.1422E-180 | .. |
| sleep | -0.474602256 | 0.024048041 | -19.73558937 | 1.20853E-86 | .. |
| higher ed | -0.716030877 | 0.016832484 | -42.53863448 | 0 | .. |
| TDI group | 0.351272639 | 0.013167559 | 26.67712758 | 1.3132E-156 | .. |
| depression | 0.659057097 | 0.027618789 | 23.86263552 | 9.7665E-126 | .. |
| hypothyroidism | 0.874150666 | 0.031309564 | 27.91960489 | 2.5367E-171 | .. |
| PCOS | 2.850905138 | 0.181005588 | 15.75037085 | 7.18313E-56 | .. |
| weightgainMeds | 0.923335671 | 0.02941129 | 31.39391958 | 5.4205E-216 | .. |
| smoke_statusFormer | 1.583891021 | 0.033549754 | 47.21021214 | 0 | .. |
| smoke_statusNever | 0.993373995 | 0.031343995 | 31.69264148 | 4.4773E-220 | .. |
| t2dm | 3.566913644 | 0.029735332 | 119.9554019 | 0 | .. |
| cushing | 2.152116409 | 0.391184895 | 5.501532488 | 3.76805E-08 | .. |
| harmonized PRS 8:sedentary | -0.019158072 | 0.023711962 | -0.807949652 | 0.419120195 | 0.465689106 |

**ST22:** Linear regression. BMI ∼ age + sex + pc’s 1-10 + clinical covariates + Hyperinsulin 2 pPS * sedentary.

| term | Beta | SE | t-statistic | p-value | q-value (BH-FDR) |
| --- | --- | --- | --- | --- | --- |
| (Intercept) | 26.5630557 | 0.076614983 | 346.7083696 | 0 | .. |
| age | -0.008584465 | 0.001046906 | -8.199841698 | 2.41622E-16 | .. |
| sex | 0.641303749 | 0.016664059 | 38.48424568 | 0 | .. |
| harmonized PRS 9 | 0.275510087 | 0.007216053 | 38.18016613 | 0 | .. |
| pc1 | 0.002609315 | 0.000164498 | 15.86231523 | 1.21741E-56 | .. |
| pc2 | 0.010923841 | 0.00031325 | 34.8726443 | 6.4685E-266 | .. |
| pc3 | 0.000579962 | 0.000580715 | 0.9987034 | 0.317939184 | .. |
| pc4 | 0.002536913 | 0.000780311 | 3.251158527 | 0.001149482 | .. |
| pc5 | 0.013866418 | 0.001060243 | 13.0785315 | 4.47496E-39 | .. |
| pc6 | 0.009216565 | 0.001618849 | 5.69328279 | 1.24733E-08 | .. |
| pc7 | 0.004322003 | 0.001634266 | 2.644614013 | 0.008178833 | .. |
| pc8 | 0.006978334 | 0.001682669 | 4.147181386 | 3.36684E-05 | .. |
| pc9 | -0.014084799 | 0.001812574 | -7.770607773 | 7.83513E-15 | .. |
| pc10 | 0.007006563 | 0.002028021 | 3.454876341 | 0.00055062 | .. |
| physical activity | -0.701161577 | 0.017449193 | -40.18303839 | 0 | .. |
| sedentary | -1.633020404 | 0.025844937 | -63.18531288 | 0 | .. |
| diet | -0.488191666 | 0.017164538 | -28.44187672 | 1.0492E-177 | .. |
| sleep | -0.473714397 | 0.024037929 | -19.70695549 | 2.12725E-86 | .. |
| higher ed | -0.715834363 | 0.016825067 | -42.54570691 | 0 | .. |
| TDI group | 0.352391051 | 0.013161742 | 26.77389231 | 9.914E-158 | .. |
| depression | 0.657422265 | 0.027606917 | 23.81367917 | 3.1371E-125 | .. |
| hypothyroidism | 0.86939191 | 0.031296539 | 27.77917064 | 1.2609E-169 | .. |
| PCOS | 2.864382098 | 0.180927822 | 15.8316287 | 1.98286E-56 | .. |
| weightgainMeds | 0.919012407 | 0.02939912 | 31.25986071 | 3.5804E-214 | .. |
| smoke_statusFormer | 1.583070714 | 0.033535278 | 47.20613121 | 0 | .. |
| smoke_statusNever | 0.991966507 | 0.031330128 | 31.66174494 | 1.1888E-219 | .. |
| t2dm | 3.570415606 | 0.029723105 | 120.1225646 | 0 | .. |
| cushing | 2.107213312 | 0.391020238 | 5.389013425 | 7.08977E-08 | .. |
| harmonized PRS 9:sedentary | -0.03628106 | 0.021556025 | -1.683105265 | 0.092355737 | 0.115444671 |

**ST23:** Linear regression. BMI ∼ age + sex + pc’s 1-10 + clinical covariates + Hypothalamic dysregulation pPS * sedentary.

| term | Beta | SE | t-statistic | p-value | q-value (BH-FDR) |
| --- | --- | --- | --- | --- | --- |
| (Intercept) | 26.576018 | 0.076634705 | 346.7882855 | 0 | .. |
| age | -0.008666956 | 0.001047183 | -8.276445451 | 1.27409E-16 | .. |
| sex | 0.641942064 | 0.016668681 | 38.5118697 | 0 | .. |
| harmonized PRS 10 | 0.280037808 | 0.007644721 | 36.63152778 | 3.9199E-293 | .. |
| pc1 | 0.002624469 | 0.000164542 | 15.95010553 | 2.99992E-57 | .. |
| pc2 | 0.010910361 | 0.000313333 | 34.82028377 | 3.9877E-265 | .. |
| pc3 | 0.000658142 | 0.000580872 | 1.133023912 | 0.2572051 | .. |
| pc4 | 0.002417545 | 0.000780514 | 3.097376402 | 0.001952595 | .. |
| pc5 | 0.013844626 | 0.001060531 | 13.05442732 | 6.14169E-39 | .. |
| pc6 | 0.009197461 | 0.001619278 | 5.679976743 | 1.34835E-08 | .. |
| pc7 | 0.004519982 | 0.001634692 | 2.765036322 | 0.005691987 | .. |
| pc8 | 0.006923275 | 0.001683123 | 4.113351668 | 3.90058E-05 | .. |
| pc9 | -0.014228791 | 0.001813064 | -7.847926075 | 4.2433E-15 | .. |
| pc10 | 0.007223547 | 0.002028572 | 3.560902644 | 0.000369638 | .. |
| physical activity | -0.695371635 | 0.017453983 | -39.8402835 | 0 | .. |
| sedentary | -1.639052787 | 0.025845421 | -63.41753052 | 0 | .. |
| diet | -0.495093761 | 0.017171042 | -28.83306486 | 1.4546E-182 | .. |
| sleep | -0.481713599 | 0.024044647 | -20.03413049 | 3.16916E-89 | .. |
| higher ed | -0.72840303 | 0.016829274 | -43.28190454 | 0 | .. |
| TDI group | 0.354148479 | 0.013165212 | 26.90032436 | 3.3426E-159 | .. |
| depression | 0.660591314 | 0.027614571 | 23.92183901 | 2.3741E-126 | .. |
| hypothyroidism | 0.872735141 | 0.03130456 | 27.87884988 | 7.8968E-171 | .. |
| PCOS | 2.869187154 | 0.180976997 | 15.85387758 | 1.39229E-56 | .. |
| weightgainMeds | 0.924583783 | 0.029406823 | 31.44113081 | 1.2339E-216 | .. |
| smoke_statusFormer | 1.579863556 | 0.033544471 | 47.09758483 | 0 | .. |
| smoke_statusNever | 0.993402713 | 0.031339171 | 31.69843671 | 3.7276E-220 | .. |
| t2dm | 3.554834852 | 0.02973082 | 119.5673346 | 0 | .. |
| cushing | 2.169791737 | 0.391124213 | 5.547577122 | 2.89894E-08 | .. |
| harmonized PRS 10:sedentary | -0.077267931 | 0.02291786 | -3.371515984 | 0.00074765 | 0.001869124 |

**ST24:** Linear regression. BMI ∼ age + sex + pc’s 1-10 + clinical covariates + Metabolically healthy pPS * sedentary.

| term | Beta | SE | t-statistic | p-value | q-value (BH-FDR) |
| --- | --- | --- | --- | --- | --- |
| (Intercept) | 26.57792978 | 0.076703294 | 346.5031094 | 0 | .. |
| age | -0.008860994 | 0.001048115 | -8.454218589 | 2.82171E-17 | .. |
| sex | 0.639185992 | 0.016683993 | 38.31133097 | 0 | .. |
| harmonized PRS 11 | 0.285342957 | 0.009795531 | 29.12991208 | 2.7058E-186 | .. |
| pc1 | 0.00260788 | 0.00016469 | 15.83511736 | 1.87597E-56 | .. |
| pc2 | 0.010927771 | 0.000313614 | 34.84470433 | 1.7079E-265 | .. |
| pc3 | 0.00056067 | 0.000581394 | 0.964354723 | 0.334868877 | .. |
| pc4 | 0.002454185 | 0.000781218 | 3.141483663 | 0.001681102 | .. |
| pc5 | 0.013836937 | 0.001061475 | 13.03557273 | 7.86443E-39 | .. |
| pc6 | 0.009031293 | 0.001620717 | 5.572407249 | 2.51454E-08 | .. |
| pc7 | 0.004497393 | 0.001636155 | 2.748757105 | 0.005982521 | .. |
| pc8 | 0.006877298 | 0.001684631 | 4.082375645 | 4.4589E-05 | .. |
| pc9 | -0.01410313 | 0.001814692 | -7.771638071 | 7.77166E-15 | .. |
| pc10 | 0.007081072 | 0.002030386 | 3.487550533 | 0.000487535 | .. |
| physical activity | -0.696539587 | 0.017469523 | -39.87170104 | 0 | .. |
| sedentary | -1.639123173 | 0.025868167 | -63.3644882 | 0 | .. |
| diet | -0.486563409 | 0.017184452 | -28.31416447 | 3.9143E-176 | .. |
| sleep | -0.475828794 | 0.024065753 | -19.77202979 | 5.87815E-87 | .. |
| higher ed | -0.722008011 | 0.016843892 | -42.86467754 | 0 | .. |
| TDI group | 0.354281459 | 0.013177008 | 26.88633566 | 4.8675E-159 | .. |
| depression | 0.654761064 | 0.027639202 | 23.68957912 | 5.9777E-124 | .. |
| hypothyroidism | 0.874854447 | 0.031332513 | 27.92161721 | 2.3982E-171 | .. |
| PCOS | 2.878009161 | 0.181139397 | 15.88836671 | 8.04018E-57 | .. |
| weightgainMeds | 0.928082813 | 0.029433376 | 31.53164639 | 7.1831E-218 | .. |
| smoke_statusFormer | 1.582551148 | 0.033574402 | 47.13564709 | 0 | .. |
| smoke_statusNever | 0.989484049 | 0.03136669 | 31.54569583 | 4.6164E-218 | .. |
| t2dm | 3.594698091 | 0.029778153 | 120.7159523 | 0 | .. |
| cushing | 2.136589843 | 0.391474374 | 5.457802567 | 4.82436E-08 | .. |
| harmonized PRS 11:sedentary | -0.12673508 | 0.029567075 | -4.286358328 | 1.81681E-05 | 0.000181681 |

**ST25:** Linear regression. BMI ∼ age + sex + pc’s 1-10 + clinical covariates + Metabolically unhealthy pPS * diet.

| term | Beta | SE | t-statistic | p-value | q-value (BH-FDR) |
| --- | --- | --- | --- | --- | --- |
| (Intercept) | 26.52603459 | 0.076268966 | 347.7959135 | 0 | .. |
| age | -0.008063594 | 0.001042182 | -7.737220196 | 1.01922E-14 | .. |
| sex | 0.641226413 | 0.0165881 | 38.65580733 | 0 | .. |
| harmonized PRS 1 | 0.282252904 | 0.005105377 | 55.28541715 | 0 | .. |
| pc1 | 0.002658284 | 0.000163749 | 16.23385256 | 3.0774E-59 | .. |
| pc2 | 0.010917539 | 0.000311821 | 35.01222604 | 5.0039E-268 | .. |
| pc3 | 0.000727575 | 0.00057807 | 1.25862726 | 0.208165957 | .. |
| pc4 | 0.002535089 | 0.000776746 | 3.263730148 | 0.001099679 | .. |
| pc5 | 0.013835263 | 0.001055407 | 13.10893878 | 2.999E-39 | .. |
| pc6 | 0.009124186 | 0.001611452 | 5.662090533 | 1.49673E-08 | .. |
| pc7 | 0.004814728 | 0.001626811 | 2.95961144 | 0.003080509 | .. |
| pc8 | 0.006959671 | 0.001674999 | 4.155030225 | 3.25333E-05 | .. |
| pc9 | -0.014006105 | 0.001804306 | -7.762599307 | 8.34618E-15 | .. |
| pc10 | 0.007236188 | 0.002018786 | 3.584424733 | 0.000337875 | .. |
| physical activity | -0.69498923 | 0.017369595 | -40.01182712 | 0 | .. |
| sedentary | -1.608914075 | 0.025724935 | -62.54297964 | 0 | .. |
| diet | -0.491819443 | 0.017086547 | -28.78401615 | 5.9633E-182 | .. |
| sleep | -0.477047119 | 0.023928175 | -19.93662741 | 2.2296E-88 | .. |
| higher ed | -0.705758213 | 0.016749733 | -42.13549082 | 0 | .. |
| TDI_group | 0.350895184 | 0.013101794 | 26.78222477 | 7.933E-158 | .. |
| depression | 0.655649503 | 0.027481137 | 23.85816468 | 1.0866E-125 | .. |
| hypothyroidism | 0.863074195 | 0.031153946 | 27.70352697 | 1.0254E-168 | .. |
| PCOS | 2.814784175 | 0.18010544 | 15.62853502 | 4.88831E-55 | .. |
| weightgainMeds | 0.90939801 | 0.029265565 | 31.07399423 | 1.1597E-211 | .. |
| smoke_statusFormer | 1.594880912 | 0.033383596 | 47.77438899 | 0 | .. |
| smoke_statusNever | 1.008004527 | 0.031189161 | 32.319065 | 9.2125E-229 | .. |
| t2dm | 3.490997157 | 0.029605721 | 117.9163024 | 0 | .. |
| cushing | 2.174640554 | 0.38923606 | 5.586945247 | 2.31294E-08 | .. |
| harmonized PRS 1:diet | -0.030132092 | 0.008516749 | -3.53798064 | 0.000403259 | 0.002016297 |

**ST26:**
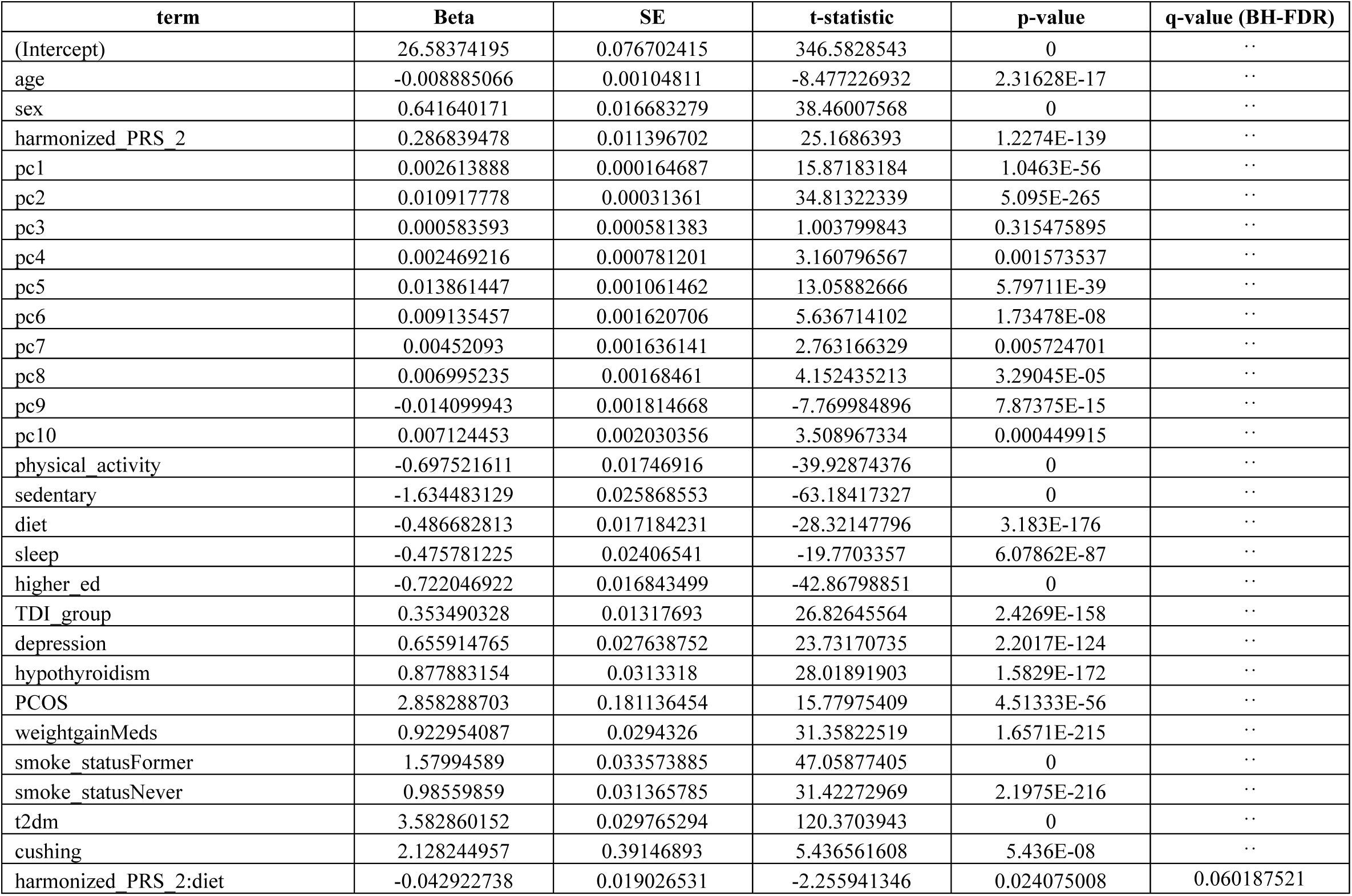
Linear regression. BMI ∼ age + sex + pc’s 1-10 + clinical covariates + Strong beta cell pPS * diet.

| term | Beta | SE | t-statistic | p-value | q-value (BH-FDR) |
| --- | --- | --- | --- | --- | --- |
| (Intercept) | 26.58374195 | 0.076702415 | 346.5828543 | 0 | .. |
| age | -0.008885066 | 0.00104811 | -8.477226932 | 2.31628E-17 | .. |
| sex | 0.641640171 | 0.016683279 | 38.46007568 | 0 | .. |
| harmonized PRS 2 | 0.286839478 | 0.011396702 | 25.1686393 | 1.2274E-139 | .. |
| pc1 | 0.002613888 | 0.000164687 | 15.87183184 | 1.0463E-56 | .. |
| pc2 | 0.010917778 | 0.00031361 | 34.81322339 | 5.095E-265 | .. |
| pc3 | 0.000583593 | 0.000581383 | 1.003799843 | 0.315475895 | .. |
| pc4 | 0.002469216 | 0.000781201 | 3.160796567 | 0.001573537 | .. |
| pc5 | 0.013861447 | 0.001061462 | 13.05882666 | 5.79711E-39 | .. |
| pc6 | 0.009135457 | 0.001620706 | 5.636714102 | 1.73478E-08 | .. |
| pc7 | 0.00452093 | 0.001636141 | 2.763166329 | 0.005724701 | .. |
| pc8 | 0.006995235 | 0.00168461 | 4.152435213 | 3.29045E-05 | .. |
| pc9 | -0.014099943 | 0.001814668 | -7.769984896 | 7.87375E-15 | .. |
| pc10 | 0.007124453 | 0.002030356 | 3.508967334 | 0.000449915 | .. |
| physical activity | -0.697521611 | 0.01746916 | -39.92874376 | 0 | .. |
| sedentary | -1.634483129 | 0.025868553 | -63.18417327 | 0 | .. |
| diet | -0.486682813 | 0.017184231 | -28.32147796 | 3.183E-176 | .. |
| sleep | -0.475781225 | 0.02406541 | -19.7703357 | 6.07862E-87 | .. |
| higher ed | -0.722046922 | 0.016843499 | -42.86798851 | 0 | .. |
| TDI group | 0.353490328 | 0.01317693 | 26.82645564 | 2.4269E-158 | .. |
| depression | 0.655914765 | 0.027638752 | 23.73170735 | 2.2017E-124 | .. |
| hypothyroidism | 0.877883154 | 0.0313318 | 28.01891903 | 1.5829E-172 | .. |
| PCOS | 2.858288703 | 0.181136454 | 15.77975409 | 4.51333E-56 | .. |
| weightgainMeds | 0.922954087 | 0.0294326 | 31.35822519 | 1.6571E-215 | .. |
| smoke_statusFormer | 1.57994589 | 0.033573885 | 47.05877405 | 0 | .. |
| smoke_statusNever | 0.98559859 | 0.031365785 | 31.42272969 | 2.1975E-216 | .. |
| t2dm | 3.582860152 | 0.029765294 | 120.3703943 | 0 | .. |
| cushing | 2.128244957 | 0.39146893 | 5.436561608 | 5.436E-08 | .. |
| harmonized PRS 2:diet | -0.042922738 | 0.019026531 | -2.255941346 | 0.024075008 | 0.060187521 |

**ST27:** Linear regression. BMI ∼ age + sex + pc’s 1-10 + clinical covariates + Hypoinsulin 1 pPS * diet.

| term | Beta | SE | t-statistic | p-value | q-value (BH-FDR) |
| --- | --- | --- | --- | --- | --- |
| (Intercept) | 26.55533186 | 0.076658947 | 346.408773 | 0 | .. |
| age | -0.008441987 | 0.001047517 | -8.059043299 | 7.71677E-16 | .. |
| sex | 0.641465813 | 0.016673331 | 38.47256616 | 0 | .. |
| harmonized PRS 3 | 0.283262099 | 0.009578854 | 29.57160775 | 6.4769E-192 | .. |
| pc1 | 0.002624696 | 0.000164588 | 15.94703886 | 3.15076E-57 | .. |
| pc2 | 0.010902947 | 0.000313421 | 34.78693362 | 1.2683E-264 | .. |
| pc3 | 0.00068123 | 0.000581036 | 1.172439593 | 0.241021525 | .. |
| pc4 | 0.0024561 | 0.00078073 | 3.145900471 | 0.001655922 | .. |
| pc5 | 0.013910917 | 0.001060828 | 13.11326477 | 2.83281E-39 | .. |
| pc6 | 0.009014143 | 0.001619719 | 5.565251578 | 2.61993E-08 | .. |
| pc7 | 0.004448484 | 0.001635147 | 2.720540893 | 0.00651789 | .. |
| pc8 | 0.006935797 | 0.001683593 | 4.11963922 | 3.79566E-05 | .. |
| pc9 | -0.014011193 | 0.001813563 | -7.725780705 | 1.11505E-14 | .. |
| pc10 | 0.007146474 | 0.002029157 | 3.521892595 | 0.00042854 | .. |
| physical activity | -0.700963845 | 0.017458785 | -40.14963588 | 0 | .. |
| sedentary | -1.634196826 | 0.025852588 | -63.21211827 | 0 | .. |
| diet | -0.488523464 | 0.017174114 | -28.44533647 | 9.5103E-178 | .. |
| sleep | -0.474167302 | 0.02405109 | -19.71500232 | 1.81487E-86 | .. |
| higher ed | -0.716521835 | 0.016834387 | -42.56298893 | 0 | .. |
| TDI group | 0.352153737 | 0.013169115 | 26.74088192 | 2.396E-157 | .. |
| depression | 0.656827006 | 0.027622116 | 23.77902555 | 7.1552E-125 | .. |
| hypothyroidism | 0.877185844 | 0.031312894 | 28.01356647 | 1.8386E-172 | .. |
| PCOS | 2.864389705 | 0.181027419 | 15.82296054 | 2.27541E-56 | .. |
| weightgainMeds | 0.919756054 | 0.029415098 | 31.2681622 | 2.7635E-214 | .. |
| smoke_statusFormer | 1.585694151 | 0.033554249 | 47.25762544 | 0 | .. |
| smoke_statusNever | 0.994198715 | 0.031348108 | 31.71479171 | 2.2219E-220 | .. |
| t2dm | 3.547904291 | 0.029741021 | 119.2932919 | 0 | .. |
| cushing | 2.158259527 | 0.391232694 | 5.516562288 | 3.45974E-08 | .. |
| harmonized PRS 3:diet | -0.032824907 | 0.015961447 | -2.056511973 | 0.03973404 | 0.066223401 |

**ST28:** Linear regression. BMI ∼ age + sex + pc’s 1-10 + clinical covariates + Hypoinsulin 2 pPS * diet.

| term | Beta | SE | t-statistic | p-value | q-value (BH-FDR) |
| --- | --- | --- | --- | --- | --- |
| (Intercept) | 26.56437713 | 0.076682277 | 346.4213385 | 0 | .. |
| age | -0.008594876 | 0.001047829 | -8.20255651 | 2.36227E-16 | .. |
| sex | 0.644308126 | 0.016678602 | 38.63082345 | 0 | .. |
| harmonized PRS 4 | 0.282639488 | 0.010449699 | 27.04761887 | 6.3142E-161 | .. |
| pc1 | 0.002623335 | 0.000164642 | 15.93356765 | 3.90805E-57 | .. |
| pc2 | 0.010916903 | 0.000313522 | 34.82016769 | 4.0038E-265 | .. |
| pc3 | 0.000635762 | 0.000581222 | 1.09383551 | 0.274027989 | .. |
| pc4 | 0.002507609 | 0.000780989 | 3.210812495 | 0.001323739 | .. |
| pc5 | 0.013816755 | 0.001061167 | 13.0203391 | 9.60091E-39 | .. |
| pc6 | 0.008949829 | 0.001620253 | 5.523723332 | 3.32159E-08 | .. |
| pc7 | 0.004403272 | 0.001635678 | 2.692015505 | 0.007102551 | .. |
| pc8 | 0.006902919 | 0.001684145 | 4.098767275 | 4.15463E-05 | .. |
| pc9 | -0.014122232 | 0.001814153 | -7.784477535 | 7.02193E-15 | .. |
| pc10 | 0.007128619 | 0.002029796 | 3.511988397 | 0.000444832 | .. |
| physical activity | -0.698088605 | 0.017464422 | -39.97204235 | 0 | .. |
| sedentary | -1.632355236 | 0.025861898 | -63.11815353 | 0 | .. |
| diet | -0.487297618 | 0.017179537 | -28.36500296 | 9.2858E-177 | .. |
| sleep | -0.476408233 | 0.024058737 | -19.80188051 | 3.25386E-87 | .. |
| higher ed | -0.719867341 | 0.016839125 | -42.74968713 | 0 | .. |
| TDI group | 0.353777159 | 0.013173267 | 26.85568863 | 1.1082E-158 | .. |
| depression | 0.661331839 | 0.027631437 | 23.93403716 | 1.7732E-126 | .. |
| hypothyroidism | 0.873567861 | 0.031323464 | 27.88860959 | 6.0174E-171 | .. |
| PCOS | 2.859611053 | 0.181086392 | 15.79141882 | 3.75211E-56 | .. |
| weightgainMeds | 0.920593809 | 0.029424686 | 31.28644483 | 1.5619E-214 | .. |
| smoke_statusFormer | 1.583242397 | 0.033564626 | 47.16997024 | 0 | .. |
| smoke_statusNever | 0.990741629 | 0.031357526 | 31.5950189 | 9.7627E-219 | .. |
| t2dm | 3.542033617 | 0.0297542 | 119.0431465 | 0 | .. |
| cushing | 2.136631682 | 0.391360097 | 5.459503156 | 4.77839E-08 | .. |
| harmonized PRS 4:diet | -0.031003709 | 0.017386273 | -1.783229173 | 0.074549997 | 0.093187497 |

**ST29:** Linear regression. BMI ∼ age + sex + pc’s 1-10 + clinical covariates + Proinsulin pPS * diet.

| term | Beta | SE | t-statistic | p-value | q-value (BH-FDR) |
| --- | --- | --- | --- | --- | --- |
| (Intercept) | 26.57652922 | 0.076801892 | 346.0400306 | 0 | .. |
| age | -0.008764283 | 0.001049461 | -8.351219109 | 6.78389E-17 | .. |
| sex | 0.642598416 | 0.016704946 | 38.46755417 | 0 | .. |
| harmonized PRS 6 | 0.297547822 | 0.034621599 | 8.594283153 | 8.41709E-18 | .. |
| pc1 | 0.002614682 | 0.000164901 | 15.85610902 | 1.34375E-56 | .. |
| pc2 | 0.010918309 | 0.000314015 | 34.77000931 | 2.2807E-264 | .. |
| pc3 | 0.00061824 | 0.000582135 | 1.062021076 | 0.288226956 | .. |
| pc4 | 0.002475792 | 0.000782211 | 3.165119965 | 0.001550342 | .. |
| pc5 | 0.013815168 | 0.001062837 | 12.99839394 | 1.27925E-38 | .. |
| pc6 | 0.009088834 | 0.001622793 | 5.600735116 | 2.13627E-08 | .. |
| pc7 | 0.004464809 | 0.001638253 | 2.725346902 | 0.006423761 | .. |
| pc8 | 0.006958593 | 0.001686787 | 4.125353798 | 3.70263E-05 | .. |
| pc9 | -0.014110056 | 0.001817014 | -7.765516755 | 8.15632E-15 | .. |
| pc10 | 0.007139576 | 0.002032991 | 3.511857318 | 0.000445051 | .. |
| physical activity | -0.697618285 | 0.017491939 | -39.88227289 | 0 | .. |
| sedentary | -1.63761011 | 0.025902132 | -63.22298622 | 0 | .. |
| diet | -0.486211462 | 0.01720689 | -28.25678869 | 1.9793E-175 | .. |
| sleep | -0.476883508 | 0.024096571 | -19.79051321 | 4.07616E-87 | .. |
| higher ed | -0.722749813 | 0.016865371 | -42.854072 | 0 | .. |
| TDI group | 0.354718438 | 0.013193904 | 26.88502499 | 5.0419E-159 | .. |
| depression | 0.657532653 | 0.027674668 | 23.75936917 | 1.1416E-124 | .. |
| hypothyroidism | 0.876964575 | 0.031372835 | 27.95299068 | 9.9936E-172 | .. |
| PCOS | 2.870015105 | 0.181372155 | 15.8239014 | 2.24168E-56 | .. |
| weightgainMeds | 0.924153694 | 0.02947085 | 31.35823036 | 1.6569E-215 | .. |
| smoke_statusFormer | 1.580708094 | 0.0336174 | 47.02053359 | 0 | .. |
| smoke_statusNever | 0.985892067 | 0.031406415 | 31.39142282 | 5.8615E-216 | .. |
| t2dm | 3.5613869 | 0.02979525 | 119.5286793 | 0 | .. |
| cushing | 2.152489157 | 0.391975342 | 5.491389187 | 3.99101E-08 | .. |
| harmonized PRS 6:diet | -0.07281682 | 0.057823046 | -1.259304466 | 0.207921342 | 0.231023714 |

**ST30:** Linear regression. BMI ∼ age + sex + pc’s 1-10 + clinical covariates + Immune dysregulation pPS * diet.

| term | Beta | SE | t-statistic | p-value | q-value (BH-FDR) |
| --- | --- | --- | --- | --- | --- |
| (Intercept) | 26.5739662 | 0.076775407 | 346.1260204 | 0 | .. |
| age | -0.008766828 | 0.001049096 | -8.356554652 | 6.48419E-17 | .. |
| sex | 0.643726156 | 0.016698967 | 38.54886088 | 0 | .. |
| harmonized PRS 7 | 0.281598449 | 0.019201746 | 14.66525247 | 1.11802E-48 | .. |
| pc1 | 0.002620956 | 0.000164843 | 15.89969495 | 6.71163E-57 | .. |
| pc2 | 0.010914455 | 0.000313906 | 34.76986057 | 2.2925E-264 | .. |
| pc3 | 0.00059338 | 0.000581933 | 1.019670692 | 0.307885473 | .. |
| pc4 | 0.002474607 | 0.000781936 | 3.164717663 | 0.001552487 | .. |
| pc5 | 0.013826947 | 0.00106246 | 13.01408208 | 1.04201E-38 | .. |
| pc6 | 0.009093808 | 0.001622222 | 5.605774488 | 2.07504E-08 | .. |
| pc7 | 0.004472819 | 0.001637674 | 2.731202662 | 0.006310727 | .. |
| pc8 | 0.00699586 | 0.001686197 | 4.148897723 | 3.3417E-05 | .. |
| pc9 | -0.014142162 | 0.001816368 | -7.785957772 | 6.94021E-15 | .. |
| pc10 | 0.007073599 | 0.002032273 | 3.480634968 | 0.000500296 | .. |
| physical activity | -0.697831059 | 0.017485638 | -39.90881229 | 0 | .. |
| sedentary | -1.638718245 | 0.025892181 | -63.29008123 | 0 | .. |
| diet | -0.485062339 | 0.017200356 | -28.20071481 | 9.6167E-175 | .. |
| sleep | -0.477259419 | 0.024088116 | -19.81306582 | 2.60651E-87 | .. |
| higher ed | -0.721874386 | 0.016859546 | -42.81695393 | 0 | .. |
| TDI group | 0.354135672 | 0.01318924 | 26.85034682 | 1.2789E-158 | .. |
| depression | 0.656967983 | 0.027664883 | 23.74736149 | 1.5184E-124 | .. |
| hypothyroidism | 0.885100167 | 0.031363577 | 28.22063877 | 5.486E-175 | .. |
| PCOS | 2.865231622 | 0.181307369 | 15.80317246 | 3.11446E-56 | .. |
| weightgainMeds | 0.928642634 | 0.029461653 | 31.52038483 | 1.0237E-217 | .. |
| smoke_statusFormer | 1.584207236 | 0.033606019 | 47.14058035 | 0 | .. |
| smoke_statusNever | 0.988062251 | 0.031395684 | 31.47127656 | 4.7905E-217 | .. |
| t2dm | 3.56020756 | 0.029784596 | 119.5318397 | 0 | .. |
| cushing | 2.132044958 | 0.391837906 | 5.441140132 | 5.29811E-08 | .. |
| harmonized PRS 7:diet | -0.028319681 | 0.032066243 | -0.883161784 | 0.377149561 | 0.377149561 |

**ST31:** Linear regression. BMI ∼ age + sex + pc’s 1-10 + clinical covariates + Hyperinsulin 1 pPS * diet.

| term | Beta | SE | t-statistic | p-value | q-value (BH-FDR) |
| --- | --- | --- | --- | --- | --- |
| (Intercept) | 26.57134852 | 0.076646515 | 346.6739312 | 0 | .. |
| age | -0.008687898 | 0.001047339 | -8.295208963 | 1.08827E-16 | .. |
| sex | 0.641444055 | 0.016671082 | 38.47645069 | 0 | .. |
| harmonized PRS 8 | 0.283114481 | 0.009269639 | 30.5421262 | 1.4658E-204 | .. |
| pc1 | 0.002610597 | 0.000164567 | 15.86343572 | 1.1959E-56 | .. |
| pc2 | 0.01093587 | 0.00031338 | 34.89648002 | 2.8238E-266 | .. |
| pc3 | 0.000618423 | 0.000580964 | 1.064477574 | 0.287113249 | .. |
| pc4 | 0.00249508 | 0.000780633 | 3.196225853 | 0.001392522 | .. |
| pc5 | 0.013887925 | 0.001060687 | 13.0933303 | 3.68338E-39 | .. |
| pc6 | 0.009131809 | 0.001619513 | 5.638613882 | 1.71575E-08 | .. |
| pc7 | 0.004419595 | 0.001634939 | 2.703217843 | 0.006867552 | .. |
| pc8 | 0.006986665 | 0.001683383 | 4.150371749 | 3.32025E-05 | .. |
| pc9 | -0.01412722 | 0.001813328 | -7.790770299 | 6.68094E-15 | .. |
| pc10 | 0.007298538 | 0.002028878 | 3.597327403 | 0.000321554 | .. |
| physical activity | -0.700277673 | 0.017456497 | -40.11558918 | 0 | .. |
| sedentary | -1.633286194 | 0.025849387 | -63.1847163 | 0 | .. |
| diet | -0.49143608 | 0.01717255 | -28.61753634 | 7.0391E-180 | .. |
| sleep | -0.474647483 | 0.024047895 | -19.7375893 | 1.1617E-86 | .. |
| higher ed | -0.716141407 | 0.016832456 | -42.54527071 | 0 | .. |
| TDI group | 0.351303871 | 0.013167463 | 26.67969357 | 1.2264E-156 | .. |
| depression | 0.65897679 | 0.027618615 | 23.8598783 | 1.0431E-125 | .. |
| hypothyroidism | 0.874424089 | 0.031309615 | 27.92829246 | 1.9909E-171 | .. |
| PCOS | 2.850133664 | 0.18100483 | 15.7461746 | 7.67547E-56 | .. |
| weightgainMeds | 0.923240303 | 0.029411118 | 31.39086021 | 5.9657E-216 | .. |
| smoke_statusFormer | 1.5843606 | 0.033549933 | 47.22395692 | 0 | .. |
| smoke_statusNever | 0.993688298 | 0.031343825 | 31.70284089 | 3.2429E-220 | .. |
| t2dm | 3.566842046 | 0.029735167 | 119.9536587 | 0 | .. |
| cushing | 2.15318932 | 0.391182824 | 5.504304348 | 3.70925E-08 | .. |
| harmonized PRS 8:diet | -0.032697713 | 0.015514809 | -2.107516334 | 0.035073659 | 0.066223401 |

**ST32:**
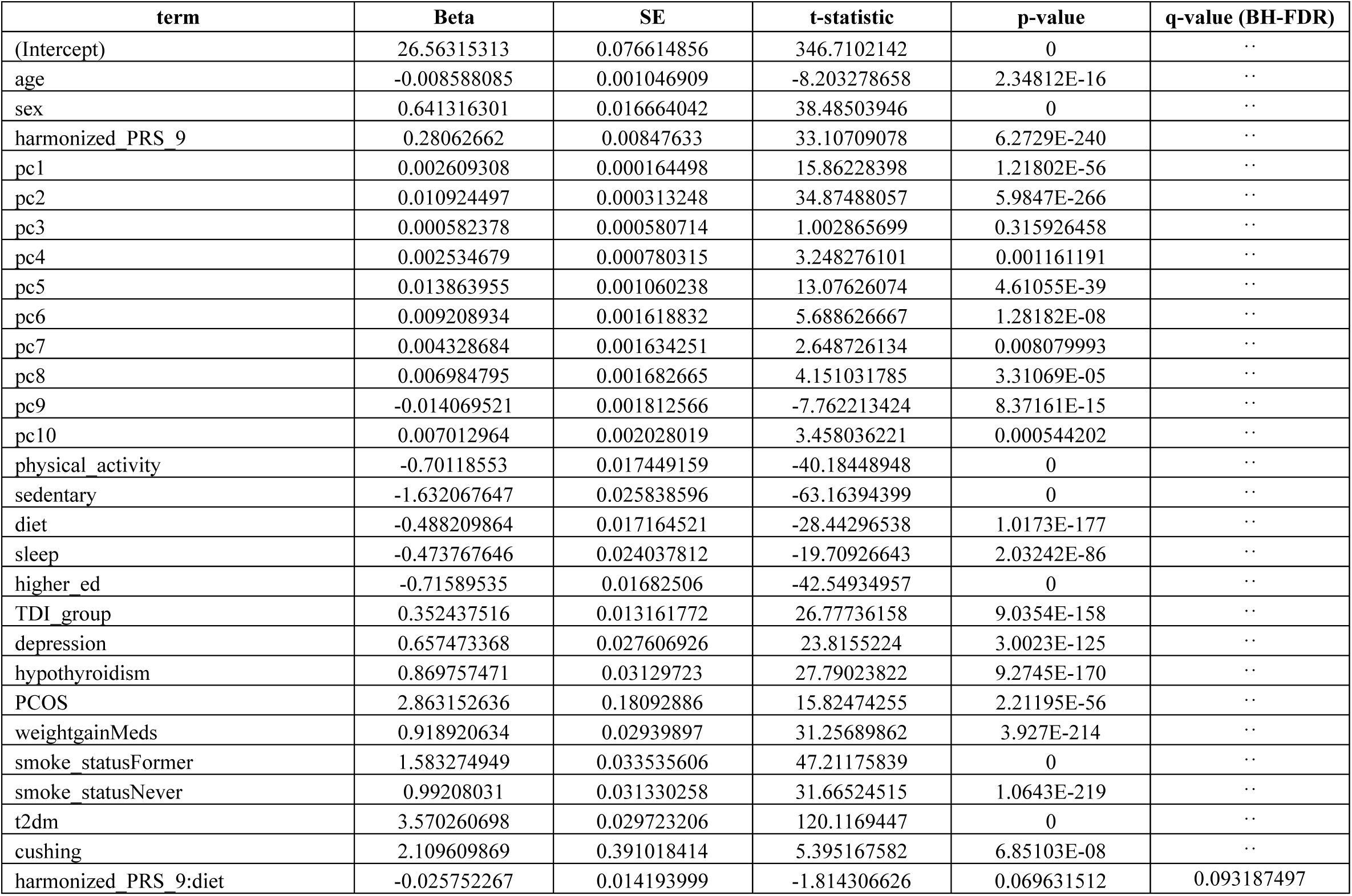
Linear regression. BMI ∼ age + sex + pc’s 1-10 + clinical covariates + Hyperinsulin 2 pPS * diet.

| term | Beta | SE | t-statistic | p-value | q-value (BH-FDR) |
| --- | --- | --- | --- | --- | --- |
| (Intercept) | 26.56315313 | 0.076614856 | 346.7102142 | 0 | .. |
| age | -0.008588085 | 0.001046909 | -8.203278658 | 2.34812E-16 | .. |
| sex | 0.641316301 | 0.016664042 | 38.48503946 | 0 | .. |
| harmonized PRS 9 | 0.28062662 | 0.00847633 | 33.10709078 | 6.2729E-240 | .. |
| pc1 | 0.002609308 | 0.000164498 | 15.86228398 | 1.21802E-56 | .. |
| pc2 | 0.010924497 | 0.000313248 | 34.87488057 | 5.9847E-266 | .. |
| pc3 | 0.000582378 | 0.000580714 | 1.002865699 | 0.315926458 | .. |
| pc4 | 0.002534679 | 0.000780315 | 3.248276101 | 0.001161191 | .. |
| pc5 | 0.013863955 | 0.001060238 | 13.07626074 | 4.61055E-39 | .. |
| pc6 | 0.009208934 | 0.001618832 | 5.688626667 | 1.28182E-08 | .. |
| pc7 | 0.004328684 | 0.001634251 | 2.648726134 | 0.008079993 | .. |
| pc8 | 0.006984795 | 0.001682665 | 4.151031785 | 3.31069E-05 | .. |
| pc9 | -0.014069521 | 0.001812566 | -7.762213424 | 8.37161E-15 | .. |
| pc10 | 0.007012964 | 0.002028019 | 3.458036221 | 0.000544202 | .. |
| physical activity | -0.70118553 | 0.017449159 | -40.18448948 | 0 | .. |
| sedentary | -1.632067647 | 0.025838596 | -63.16394399 | 0 | .. |
| diet | -0.488209864 | 0.017164521 | -28.44296538 | 1.0173E-177 | .. |
| sleep | -0.473767646 | 0.024037812 | -19.70926643 | 2.03242E-86 | .. |
| higher ed | -0.71589535 | 0.01682506 | -42.54934957 | 0 | .. |
| TDI group | 0.352437516 | 0.013161772 | 26.77736158 | 9.0354E-158 | .. |
| depression | 0.657473368 | 0.027606926 | 23.8155224 | 3.0023E-125 | .. |
| hypothyroidism | 0.869757471 | 0.03129723 | 27.79023822 | 9.2745E-170 | .. |
| PCOS | 2.863152636 | 0.18092886 | 15.82474255 | 2.21195E-56 | .. |
| weightgainMeds | 0.918920634 | 0.02939897 | 31.25689862 | 3.927E-214 | .. |
| smoke_statusFormer | 1.583274949 | 0.033535606 | 47.21175839 | 0 | .. |
| smoke_statusNever | 0.99208031 | 0.031330258 | 31.66524515 | 1.0643E-219 | .. |
| t2dm | 3.570260698 | 0.029723206 | 120.1169447 | 0 | .. |
| cushing | 2.109609869 | 0.391018414 | 5.395167582 | 6.85103E-08 | .. |
| harmonized PRS 9:diet | -0.025752267 | 0.014193999 | -1.814306626 | 0.069631512 | 0.093187497 |

**ST33:**
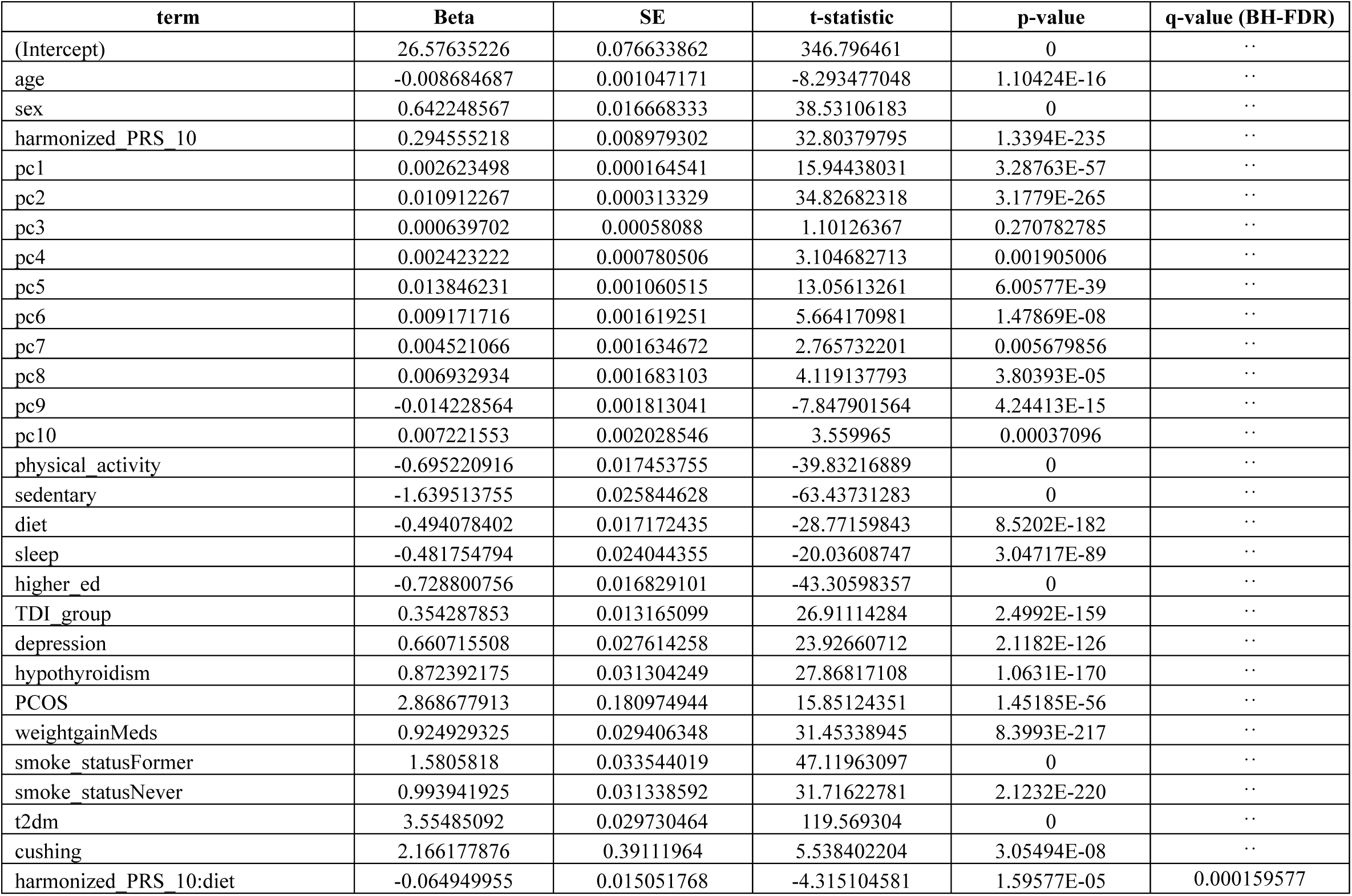
Linear regression. BMI ∼ age + sex + pc’s 1-10 + clinical covariates + Hypothalamic dysregulation pPS * diet.

**ST34:** Linear regression. BMI ∼ age + sex + pc’s 1-10 + clinical covariates + Metabolically healthy pPS * diet.

| term | Beta | SE | t-statistic | p-value | q-value (BH-FDR) |
| --- | --- | --- | --- | --- | --- |
| (Intercept) | 26.5779617 | 0.076704627 | 346.4975045 | 0 | .. |
| age | -0.008862794 | 0.001048135 | -8.455775198 | 2.78433E-17 | .. |
| sex | 0.639137505 | 0.01668431 | 38.30769695 | 0 | .. |
| harmonized PRS 11 | 0.291173633 | 0.011519731 | 25.27608008 | 8.179E-141 | .. |
| pc1 | 0.0026069 | 0.000164693 | 15.8288328 | 2.07287E-56 | .. |
| pc2 | 0.010929779 | 0.00031362 | 34.85044037 | 1.3993E-265 | .. |
| pc3 | 0.000565893 | 0.000581402 | 0.973324689 | 0.330392726 | .. |
| pc4 | 0.002442582 | 0.000781225 | 3.126604539 | 0.001768542 | .. |
| pc5 | 0.013834764 | 0.001061493 | 13.03331091 | 8.10099E-39 | .. |
| pc6 | 0.009028038 | 0.001620744 | 5.5703043 | 2.54508E-08 | .. |
| pc7 | 0.004505076 | 0.001636182 | 2.753406605 | 0.005898208 | .. |
| pc8 | 0.006891622 | 0.001684667 | 4.090791122 | 4.30014E-05 | .. |
| pc9 | -0.014121411 | 0.001814713 | -7.781621196 | 7.18231E-15 | .. |
| pc10 | 0.007079081 | 0.00203042 | 3.486511408 | 0.000489433 | .. |
| physical activity | -0.696537662 | 0.017469828 | -39.87089504 | 0 | .. |
| sedentary | -1.638948286 | 0.025868538 | -63.35681842 | 0 | .. |
| diet | -0.486560622 | 0.017184746 | -28.31351881 | 3.9865E-176 | .. |
| sleep | -0.476000499 | 0.024066139 | -19.7788477 | 5.13584E-87 | .. |
| higher ed | -0.722437597 | 0.016844005 | -42.88989412 | 0 | .. |
| TDI group | 0.354295573 | 0.013177252 | 26.8869079 | 4.7933E-159 | .. |
| depression | 0.654690494 | 0.02763969 | 23.68660735 | 6.4137E-124 | .. |
| hypothyroidism | 0.875215599 | 0.031332906 | 27.93279394 | 1.7559E-171 | .. |
| PCOS | 2.877522628 | 0.181142342 | 15.88542244 | 8.42642E-57 | .. |
| weightgainMeds | 0.927908757 | 0.029433969 | 31.52509834 | 8.8261E-218 | .. |
| smoke_statusFormer | 1.582942667 | 0.03357497 | 47.14651054 | 0 | .. |
| smoke_statusNever | 0.989998433 | 0.031367132 | 31.56164945 | 2.7937E-218 | .. |
| t2dm | 3.594018093 | 0.029778119 | 120.6932541 | 0 | .. |
| cushing | 2.137515805 | 0.391482458 | 5.460055131 | 4.76356E-08 | .. |
| harmonized PRS 11:diet | -0.055337328 | 0.019283167 | -2.869721884 | 0.004108611 | 0.013695371 |

**ST35:** Linear regression. BMI ∼ age + sex + pc’s 1-10 + clinical covariates + Metabolically unhealthy pPS * sleep.

| term | Beta | SE | t-statistic | p-value | q-value (BH-FDR) |
| --- | --- | --- | --- | --- | --- |
| (Intercept) | 26.52536192 | 0.076270537 | 347.7799299 | 0 | .. |
| age | -0.008057585 | 0.001042195 | -7.731360246 | 1.06725E-14 | .. |
| sex | 0.64129223 | 0.016588453 | 38.65895264 | 0 | .. |
| harmonized PRS 1 | 0.294151126 | 0.011351657 | 25.91261527 | 6.9453E-148 | .. |
| pc1 | 0.002658297 | 0.000163752 | 16.233718 | 3.08415E-59 | .. |
| pc2 | 0.010919134 | 0.000311825 | 35.01683909 | 4.2598E-268 | .. |
| pc3 | 0.000727112 | 0.000578078 | 1.257809443 | 0.208461641 | .. |
| pc4 | 0.00253231 | 0.000776758 | 3.260102198 | 0.001113843 | .. |
| pc5 | 0.013829752 | 0.001055421 | 13.10353604 | 3.22025E-39 | .. |
| pc6 | 0.009125121 | 0.001611473 | 5.66259776 | 1.49232E-08 | .. |
| pc7 | 0.004815435 | 0.001626834 | 2.960004204 | 0.003076584 | .. |
| pc8 | 0.006971708 | 0.001675025 | 4.162151109 | 3.1535E-05 | .. |
| pc9 | -0.014016912 | 0.001804332 | -7.768476832 | 7.96803E-15 | .. |
| pc10 | 0.007252346 | 0.002018805 | 3.592395183 | 0.000327704 | .. |
| physical activity | -0.695037757 | 0.017369829 | -40.01408099 | 0 | .. |
| sedentary | -1.608597357 | 0.025725024 | -62.53045002 | 0 | .. |
| diet | -0.492022872 | 0.017086687 | -28.79568589 | 4.2638E-182 | .. |
| sleep | -0.476355753 | 0.023930103 | -19.90613065 | 4.09634E-88 | .. |
| higher ed | -0.705656549 | 0.016749983 | -42.12879146 | 0 | .. |
| TDI group | 0.350972074 | 0.013101941 | 26.78779153 | 6.8351E-158 | .. |
| depression | 0.655488813 | 0.027481494 | 23.85200772 | 1.2585E-125 | .. |
| hypothyroidism | 0.863139933 | 0.031154366 | 27.7052641 | 9.7732E-169 | .. |
| PCOS | 2.812533269 | 0.180108046 | 15.61581134 | 5.96714E-55 | .. |
| weightgainMeds | 0.909055271 | 0.029266255 | 31.06155129 | 1.7057E-211 | .. |
| smoke_statusFormer | 1.594285373 | 0.033383453 | 47.75675434 | 0 | .. |
| smoke_statusNever | 1.007464716 | 0.031189092 | 32.30182869 | 1.6056E-228 | .. |
| t2dm | 3.490981239 | 0.029606253 | 117.9136458 | 0 | .. |
| cushing | 2.174272645 | 0.389241083 | 5.585927954 | 2.32652E-08 | .. |
| harmonized PRS 1:sleep | -0.026084769 | 0.012164669 | -2.144305642 | 0.032009191 | 0.045727416 |

**ST36:**
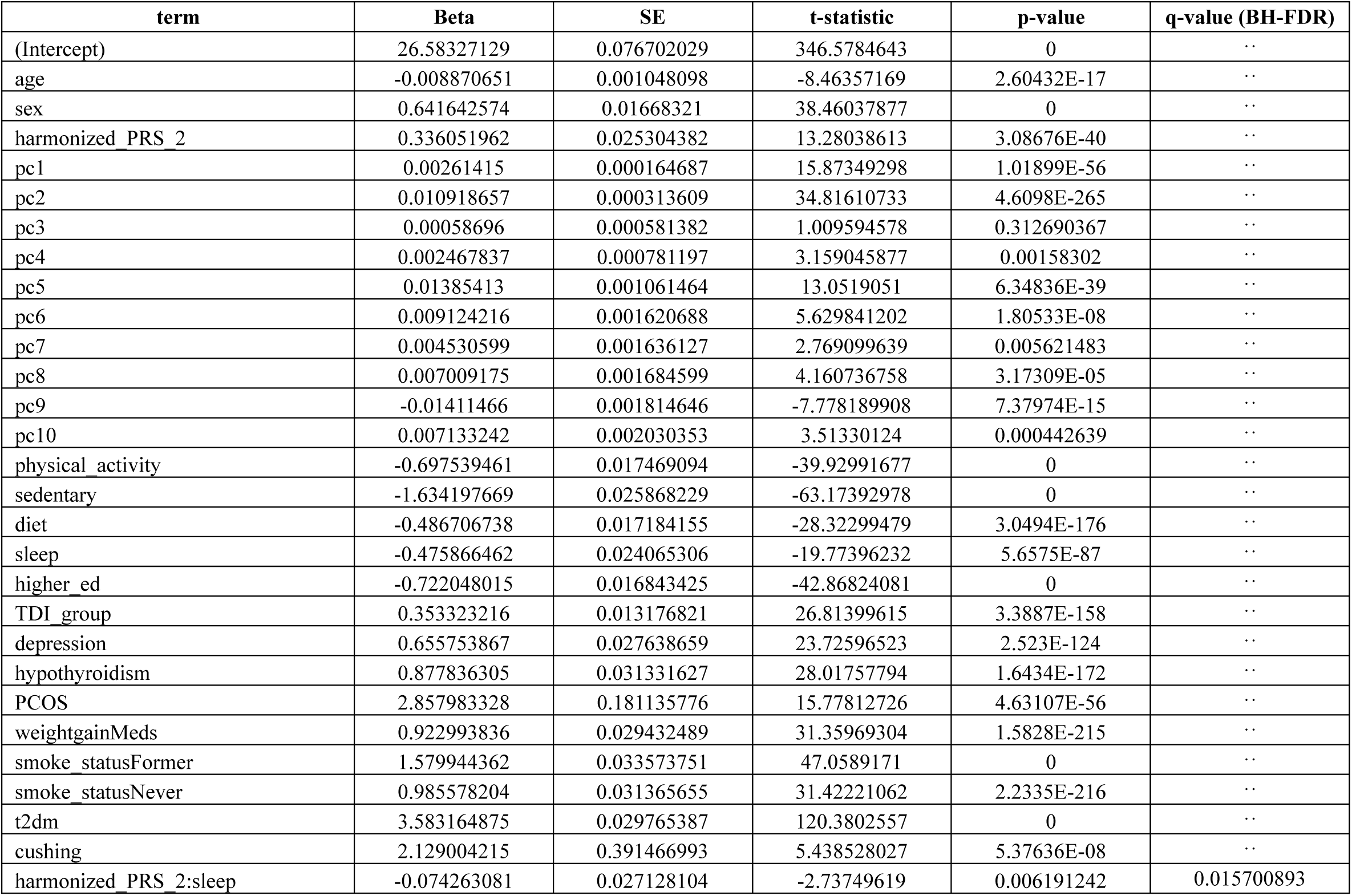
Linear regression. BMI ∼ age + sex + pc’s 1-10 + clinical covariates + Strong beta cell pPS * sleep.

| term | Beta | SE | t-statistic | p-value | q-value (BH-FDR) |
| --- | --- | --- | --- | --- | --- |
| (Intercept) | 26.58327129 | 0.076702029 | 346.5784643 | 0 | .. |
| age | -0.008870651 | 0.001048098 | -8.46357169 | 2.60432E-17 | .. |
| sex | 0.641642574 | 0.01668321 | 38.46037877 | 0 | .. |
| harmonized PRS 2 | 0.336051962 | 0.025304382 | 13.28038613 | 3.08676E-40 | .. |
| pc1 | 0.00261415 | 0.000164687 | 15.87349298 | 1.01899E-56 | .. |
| pc2 | 0.010918657 | 0.000313609 | 34.81610733 | 4.6098E-265 | .. |
| pc3 | 0.00058696 | 0.000581382 | 1.009594578 | 0.312690367 | .. |
| pc4 | 0.002467837 | 0.000781197 | 3.159045877 | 0.00158302 | .. |
| pc5 | 0.01385413 | 0.001061464 | 13.0519051 | 6.34836E-39 | .. |
| pc6 | 0.009124216 | 0.001620688 | 5.629841202 | 1.80533E-08 | .. |
| pc7 | 0.004530599 | 0.001636127 | 2.769099639 | 0.005621483 | .. |
| pc8 | 0.007009175 | 0.001684599 | 4.160736758 | 3.17309E-05 | .. |
| pc9 | -0.01411466 | 0.001814646 | -7.778189908 | 7.37974E-15 | .. |
| pc10 | 0.007133242 | 0.002030353 | 3.51330124 | 0.000442639 | .. |
| physical activity | -0.697539461 | 0.017469094 | -39.92991677 | 0 | .. |
| sedentary | -1.634197669 | 0.025868229 | -63.17392978 | 0 | .. |
| diet | -0.486706738 | 0.017184155 | -28.32299479 | 3.0494E-176 | .. |
| sleep | -0.475866462 | 0.024065306 | -19.77396232 | 5.6575E-87 | .. |
| higher ed | -0.722048015 | 0.016843425 | -42.86824081 | 0 | .. |
| TDI group | 0.353323216 | 0.013176821 | 26.81399615 | 3.3887E-158 | .. |
| depression | 0.655753867 | 0.027638659 | 23.72596523 | 2.523E-124 | .. |
| hypothyroidism | 0.877836305 | 0.031331627 | 28.01757794 | 1.6434E-172 | .. |
| PCOS | 2.857983328 | 0.181135776 | 15.77812726 | 4.63107E-56 | .. |
| weightgainMeds | 0.922993836 | 0.029432489 | 31.35969304 | 1.5828E-215 | .. |
| smoke_statusFormer | 1.579944362 | 0.033573751 | 47.0589171 | 0 | .. |
| smoke_statusNever | 0.985578204 | 0.031365655 | 31.42221062 | 2.2335E-216 | .. |
| t2dm | 3.583164875 | 0.029765387 | 120.3802557 | 0 | .. |
| cushing | 2.129004215 | 0.391466993 | 5.438528027 | 5.37636E-08 | .. |
| harmonized PRS 2:sleep | -0.074263081 | 0.027128104 | -2.73749619 | 0.006191242 | 0.015700893 |

**ST37:** Linear regression. BMI ∼ age + sex + pc’s 1-10 + clinical covariates + Hypoinsulin 1 pPS * sleep.

| term | Beta | SE | t-statistic | p-value | q-value (BH-FDR) |
| --- | --- | --- | --- | --- | --- |
| (Intercept) | 26.5547325 | 0.076659771 | 346.3972321 | 0 | .. |
| age | -0.008434734 | 0.001047519 | -8.052103443 | 8.16719E-16 | .. |
| sex | 0.641374719 | 0.016673227 | 38.46734141 | 0 | .. |
| harmonized PRS 3 | 0.314026634 | 0.021186146 | 14.82226299 | 1.09453E-49 | .. |
| pc1 | 0.00262445 | 0.000164588 | 15.94555089 | 3.22665E-57 | .. |
| pc2 | 0.010902761 | 0.000313421 | 34.7863619 | 1.2937E-264 | .. |
| pc3 | 0.000681648 | 0.000581036 | 1.173159947 | 0.240732585 | .. |
| pc4 | 0.00245349 | 0.00078073 | 3.142560332 | 0.001674932 | .. |
| pc5 | 0.013906761 | 0.001060823 | 13.10940438 | 2.98066E-39 | .. |
| pc6 | 0.00901701 | 0.001619718 | 5.567025643 | 2.59341E-08 | .. |
| pc7 | 0.004446278 | 0.001635145 | 2.719194698 | 0.006544477 | .. |
| pc8 | 0.006924614 | 0.001683593 | 4.112998073 | 3.90656E-05 | .. |
| pc9 | -0.014014215 | 0.001813562 | -7.727453298 | 1.1005E-14 | .. |
| pc10 | 0.007167249 | 0.002029132 | 3.532174749 | 0.000412218 | .. |
| physical activity | -0.70095507 | 0.017458764 | -40.14917966 | 0 | .. |
| sedentary | -1.634309853 | 0.02585264 | -63.21636226 | 0 | .. |
| diet | -0.488678716 | 0.017174106 | -28.45439076 | 7.3533E-178 | .. |
| sleep | -0.473450355 | 0.024052535 | -19.68401092 | 3.34451E-86 | .. |
| higher ed | -0.71651706 | 0.016834372 | -42.56274248 | 0 | .. |
| TDI group | 0.352113353 | 0.013169054 | 26.73793852 | 2.592E-157 | .. |
| depression | 0.656901241 | 0.027622071 | 23.78175177 | 6.7061E-125 | .. |
| hypothyroidism | 0.877023774 | 0.03131292 | 28.00836738 | 2.1265E-172 | .. |
| PCOS | 2.863950159 | 0.181027194 | 15.8205521 | 2.36408E-56 | .. |
| weightgainMeds | 0.919613265 | 0.02941519 | 31.26320982 | 3.2253E-214 | .. |
| smoke_statusFormer | 1.585200144 | 0.0335539 | 47.24339474 | 0 | .. |
| smoke_statusNever | 0.993821011 | 0.031347816 | 31.703038 | 3.2227E-220 | .. |
| t2dm | 3.548154293 | 0.029740955 | 119.3019609 | 0 | .. |
| cushing | 2.158284122 | 0.391232428 | 5.516628905 | 3.45843E-08 | .. |
| harmonized PRS 3:sleep | -0.048988533 | 0.022723246 | -2.155877425 | 0.031094015 | 0.045727416 |

**ST38:** Linear regression. BMI ∼ age + sex + pc’s 1-10 + clinical covariates + Hypoinsulin 2 pPS * sleep.

| term | Beta | SE | t-statistic | p-value | q-value (BH-FDR) |
| --- | --- | --- | --- | --- | --- |
| (Intercept) | 26.56351855 | 0.076680611 | 346.4176694 | 0 | .. |
| age | -0.008588887 | 0.001047803 | -8.197045676 | 2.47305E-16 | .. |
| sex | 0.644331302 | 0.016678179 | 38.63319185 | 0 | .. |
| harmonized PRS 4 | 0.364653885 | 0.023146985 | 15.7538391 | 6.80005E-56 | .. |
| pc1 | 0.002624631 | 0.000164638 | 15.94184247 | 3.4238E-57 | .. |
| pc2 | 0.010914428 | 0.000313515 | 34.81311319 | 5.1146E-265 | .. |
| pc3 | 0.000642971 | 0.00058121 | 1.106262422 | 0.268613822 | .. |
| pc4 | 0.002526057 | 0.000780971 | 3.234506183 | 0.001218662 | .. |
| pc5 | 0.013821761 | 0.001061139 | 13.02539843 | 8.9856E-39 | .. |
| pc6 | 0.008943245 | 0.001620209 | 5.519810358 | 3.3964E-08 | .. |
| pc7 | 0.004413741 | 0.001635639 | 2.69848151 | 0.006966043 | .. |
| pc8 | 0.006899165 | 0.001684098 | 4.096652707 | 4.19275E-05 | .. |
| pc9 | -0.014091161 | 0.001814116 | -7.767508578 | 8.02914E-15 | .. |
| pc10 | 0.007136278 | 0.002029742 | 3.515854014 | 0.000438405 | .. |
| physical activity | -0.698139376 | 0.017463869 | -39.97621484 | 0 | .. |
| sedentary | -1.632330522 | 0.025860962 | -63.11948172 | 0 | .. |
| diet | -0.487360677 | 0.017179089 | -28.36941377 | 8.1951E-177 | .. |
| sleep | -0.475958881 | 0.024058363 | -19.78351071 | 4.68276E-87 | .. |
| higher ed | -0.719758627 | 0.016838686 | -42.74434713 | 0 | .. |
| TDI group | 0.353735468 | 0.013172877 | 26.85331989 | 1.1809E-158 | .. |
| depression | 0.66126913 | 0.027630739 | 23.93237205 | 1.8453E-126 | .. |
| hypothyroidism | 0.873751959 | 0.031322704 | 27.89516379 | 5.0132E-171 | .. |
| PCOS | 2.85753884 | 0.181082224 | 15.78033877 | 4.47175E-56 | .. |
| weightgainMeds | 0.920308324 | 0.02942391 | 31.27756685 | 2.0607E-214 | .. |
| smoke_statusFormer | 1.583396597 | 0.033563789 | 47.17574024 | 0 | .. |
| smoke_statusNever | 0.990923486 | 0.031356756 | 31.6015945 | 7.9348E-219 | .. |
| t2dm | 3.541517301 | 0.029753655 | 119.0279734 | 0 | .. |
| cushing | 2.135201659 | 0.391350342 | 5.455985158 | 4.87397E-08 | .. |
| harmonized PRS 4:sleep | -0.107153865 | 0.024816654 | -4.317820726 | 1.57627E-05 | 0.000157627 |

**ST39:** Linear regression. BMI ∼ age + sex + pc’s 1-10 + clinical covariates + Proinsulin pPS * sleep.

| term | Beta | SE | t-statistic | p-value | q-value (BH-FDR) |
| --- | --- | --- | --- | --- | --- |
| (Intercept) | 26.57642965 | 0.076802297 | 346.0369107 | 0 | .. |
| age | -0.008763661 | 0.001049466 | -8.35059019 | 6.8201E-17 | .. |
| sex | 0.642565324 | 0.016705094 | 38.46523216 | 0 | .. |
| harmonized PRS 6 | 0.259049364 | 0.077106353 | 3.359637104 | 0.000780545 | .. |
| pc1 | 0.00261507 | 0.000164901 | 15.85844773 | 1.29468E-56 | .. |
| pc2 | 0.010918758 | 0.000314016 | 34.77129166 | 2.1815E-264 | .. |
| pc3 | 0.000617859 | 0.000582137 | 1.061363426 | 0.28852561 | .. |
| pc4 | 0.00247547 | 0.000782214 | 3.164696587 | 0.0015526 | .. |
| pc5 | 0.013818603 | 0.001062836 | 13.00163325 | 1.22622E-38 | .. |
| pc6 | 0.009090221 | 0.0016228 | 5.601565504 | 2.12606E-08 | .. |
| pc7 | 0.004468794 | 0.001638255 | 2.727776075 | 0.006376652 | .. |
| pc8 | 0.006961978 | 0.001686791 | 4.127351889 | 3.67062E-05 | .. |
| pc9 | -0.014119681 | 0.001817015 | -7.770810957 | 7.82258E-15 | .. |
| pc10 | 0.007132078 | 0.002032988 | 3.508175673 | 0.000451256 | .. |
| physical activity | -0.697641291 | 0.017491987 | -39.88347787 | 0 | .. |
| sedentary | -1.637528824 | 0.025902116 | -63.21988709 | 0 | .. |
| diet | -0.486295728 | 0.017206831 | -28.26178301 | 1.7191E-175 | .. |
| sleep | -0.476878812 | 0.024096773 | -19.79015285 | 4.10537E-87 | .. |
| higher ed | -0.722694511 | 0.016865375 | -42.85078241 | 0 | .. |
| TDI group | 0.35470313 | 0.01319395 | 26.88377106 | 5.2146E-159 | .. |
| depression | 0.657614939 | 0.02767467 | 23.76234083 | 1.0638E-124 | .. |
| hypothyroidism | 0.876953805 | 0.031372914 | 27.95257727 | 1.011E-171 | .. |
| PCOS | 2.869317657 | 0.181371856 | 15.82008217 | 2.38177E-56 | .. |
| weightgainMeds | 0.924030276 | 0.029470764 | 31.3541333 | 1.8835E-215 | .. |
| smoke_statusFormer | 1.580805645 | 0.033617518 | 47.02327025 | 0 | .. |
| smoke_statusNever | 0.98593538 | 0.031406519 | 31.39269802 | 5.632E-216 | .. |
| t2dm | 3.561527747 | 0.029795165 | 119.5337487 | 0 | .. |
| cushing | 2.152758264 | 0.391976349 | 5.492061625 | 3.97584E-08 | .. |
| harmonized PRS 6:sleep | 0.01423703 | 0.082634166 | 0.172289871 | 0.863209761 | 0.863209761 |

**ST40:** Linear regression. BMI ∼ age + sex + pc’s 1-10 + clinical covariates + Immune dysregulation pPS * sleep.

| term | Beta | SE | t-statistic | p-value | q-value (BH-FDR) |
| --- | --- | --- | --- | --- | --- |
| (Intercept) | 26.57453465 | 0.076774727 | 346.1364929 | 0 | .. |
| age | -0.008769384 | 0.001049079 | -8.359126341 | 6.34444E-17 | .. |
| sex | 0.643812724 | 0.016698757 | 38.55452921 | 0 | .. |
| harmonized PRS 7 | 0.379975804 | 0.042588138 | 8.922104249 | 4.59948E-19 | .. |
| pc1 | 0.002622376 | 0.000164842 | 15.90846801 | 5.83505E-57 | .. |
| pc2 | 0.01091648 | 0.000313902 | 34.77673697 | 1.8063E-264 | .. |
| pc3 | 0.000594925 | 0.000581925 | 1.022340465 | 0.306620596 | .. |
| pc4 | 0.002479099 | 0.00078193 | 3.170488998 | 0.001521976 | .. |
| pc5 | 0.013821076 | 0.001062451 | 13.00866892 | 1.11846E-38 | .. |
| pc6 | 0.009097212 | 0.001622204 | 5.607933344 | 2.04933E-08 | .. |
| pc7 | 0.004471053 | 0.001637656 | 2.730154436 | 0.006330829 | .. |
| pc8 | 0.006990094 | 0.001686178 | 4.145524664 | 3.39128E-05 | .. |
| pc9 | -0.014141887 | 0.001816345 | -7.785904797 | 6.94312E-15 | .. |
| pc10 | 0.007051681 | 0.002032263 | 3.469867027 | 0.000520788 | .. |
| physical activity | -0.697740093 | 0.017485482 | -39.90396738 | 0 | .. |
| sedentary | -1.638893328 | 0.025891881 | -63.29757624 | 0 | .. |
| diet | -0.485058955 | 0.017200154 | -28.2008503 | 9.5801E-175 | .. |
| sleep | -0.477614243 | 0.024088225 | -19.8277062 | 1.94931E-87 | .. |
| higher ed | -0.721853901 | 0.016859312 | -42.81633308 | 0 | .. |
| TDI group | 0.3541257 | 0.013189094 | 26.84988887 | 1.2947E-158 | .. |
| depression | 0.656916656 | 0.027664471 | 23.74586017 | 1.5735E-124 | .. |
| hypothyroidism | 0.885541543 | 0.031363222 | 28.23503088 | 3.6564E-175 | .. |
| PCOS | 2.862864664 | 0.181307693 | 15.79008927 | 3.83197E-56 | .. |
| weightgainMeds | 0.928522363 | 0.029461269 | 31.51671309 | 1.1489E-217 | .. |
| smoke_statusFormer | 1.584098739 | 0.03360552 | 47.13805205 | 0 | .. |
| smoke_statusNever | 0.987958815 | 0.031395347 | 31.4683198 | 5.2565E-217 | .. |
| t2dm | 3.560166389 | 0.029784274 | 119.5317491 | 0 | .. |
| cushing | 2.132855818 | 0.39183374 | 5.443267391 | 5.23521E-08 | .. |
| harmonized PRS 7:sleep | -0.124802244 | 0.045668401 | -2.732792069 | 0.006280357 | 0.015700893 |

**ST41:** Linear regression. BMI ∼ age + sex + pc’s 1-10 + clinical covariates + Hyperinsulin 1 pPS * sleep.

| term | Beta | SE | t-statistic | p-value | q-value (BH-FDR) |
| --- | --- | --- | --- | --- | --- |
| (Intercept) | 26.57063509 | 0.076647075 | 346.6620899 | 0 | .. |
| age | -0.008681477 | 0.00104734 | -8.289073035 | 1.14589E-16 | .. |
| sex | 0.641473597 | 0.016671102 | 38.47817673 | 0 | .. |
| harmonized PRS 8 | 0.307130486 | 0.020619932 | 14.89483523 | 3.70814E-50 | .. |
| pc1 | 0.002611411 | 0.000164568 | 15.86830483 | 1.10673E-56 | .. |
| pc2 | 0.010937265 | 0.000313381 | 34.90080518 | 2.4293E-266 | .. |
| pc3 | 0.000620586 | 0.000580962 | 1.068204002 | 0.285429343 | .. |
| pc4 | 0.002496187 | 0.000780634 | 3.197641074 | 0.001385707 | .. |
| pc5 | 0.013884771 | 0.001060688 | 13.09034851 | 3.83079E-39 | .. |
| pc6 | 0.009122937 | 0.001619514 | 5.633133503 | 1.77119E-08 | .. |
| pc7 | 0.004431044 | 0.001634941 | 2.710217356 | 0.006724288 | .. |
| pc8 | 0.006968941 | 0.001683381 | 4.139847345 | 3.4763E-05 | .. |
| pc9 | -0.014136945 | 0.001813338 | -7.796091297 | 6.40537E-15 | .. |
| pc10 | 0.007295793 | 0.002028881 | 3.595969354 | 0.000323237 | .. |
| physical activity | -0.70011916 | 0.017456507 | -40.10648721 | 0 | .. |
| sedentary | -1.633404794 | 0.025849506 | -63.18901396 | 0 | .. |
| diet | -0.491592437 | 0.017172536 | -28.62666504 | 5.4226E-180 | .. |
| sleep | -0.474324689 | 0.024048299 | -19.72383504 | 1.52442E-86 | .. |
| higher ed | -0.715889813 | 0.016832416 | -42.53042587 | 0 | .. |
| TDI group | 0.351317664 | 0.013167491 | 26.68068311 | 1.1944E-156 | .. |
| depression | 0.659173585 | 0.027618751 | 23.86688622 | 8.8245E-126 | .. |
| hypothyroidism | 0.873881825 | 0.031309275 | 27.91127595 | 3.1997E-171 | .. |
| PCOS | 2.849971725 | 0.181005452 | 15.74522587 | 7.79136E-56 | .. |
| weightgainMeds | 0.923461043 | 0.02941125 | 31.39822513 | 4.7366E-216 | .. |
| smoke_statusFormer | 1.5840115 | 0.033549477 | 47.21419319 | 0 | .. |
| smoke_statusNever | 0.993453545 | 0.031343684 | 31.69549389 | 4.0911E-220 | .. |
| t2dm | 3.566987927 | 0.029735223 | 119.9583393 | 0 | .. |
| cushing | 2.151299909 | 0.391182985 | 5.499472089 | 3.81234E-08 | .. |
| harmonized PRS 8:sleep | -0.04101443 | 0.022105314 | -1.855410422 | 0.063538602 | 0.079423253 |

**ST42:** Linear regression. BMI ∼ age + sex + pc’s 1-10 + clinical covariates + Hyperinsulin 2 pPS * sleep.

| term | Beta | SE | t-statistic | p-value | q-value (BH-FDR) |
| --- | --- | --- | --- | --- | --- |
| (Intercept) | 26.56295189 | 0.076615811 | 346.7032661 | 0 | .. |
| age | -0.008581838 | 0.00104691 | -8.197303497 | 2.46776E-16 | .. |
| sex | 0.641298079 | 0.016664138 | 38.48372427 | 0 | .. |
| harmonized PRS 9 | 0.287846183 | 0.018828834 | 15.28752059 | 9.68747E-53 | .. |
| pc1 | 0.002610168 | 0.000164499 | 15.86734324 | 1.1238E-56 | .. |
| pc2 | 0.010926624 | 0.000313252 | 34.88131207 | 4.7855E-266 | .. |
| pc3 | 0.000580459 | 0.000580717 | 0.999555551 | 0.317526434 | .. |
| pc4 | 0.002540882 | 0.000780308 | 3.256255075 | 0.001129046 | .. |
| pc5 | 0.013860646 | 0.001060242 | 13.07309854 | 4.80621E-39 | .. |
| pc6 | 0.009205838 | 0.001618838 | 5.686695692 | 1.29639E-08 | .. |
| pc7 | 0.004332755 | 0.001634256 | 2.651210112 | 0.008020806 | .. |
| pc8 | 0.006980485 | 0.001682675 | 4.148444954 | 3.34832E-05 | .. |
| pc9 | -0.014076155 | 0.001812571 | -7.765852038 | 8.13477E-15 | .. |
| pc10 | 0.007017705 | 0.002028043 | 3.46033376 | 0.00053958 | .. |
| physical activity | -0.701181172 | 0.017449257 | -40.18401414 | 0 | .. |
| sedentary | -1.63200071 | 0.025838772 | -63.16092454 | 0 | .. |
| diet | -0.488196348 | 0.017164597 | -28.44205164 | 1.044E-177 | .. |
| sleep | -0.473692935 | 0.024038498 | -19.70559647 | 2.18507E-86 | .. |
| higher ed | -0.715864584 | 0.016825115 | -42.54738008 | 0 | .. |
| TDI group | 0.352390748 | 0.01316179 | 26.77377165 | 9.9461E-158 | .. |
| depression | 0.657313426 | 0.027607133 | 23.80955038 | 3.4611E-125 | .. |
| hypothyroidism | 0.869319099 | 0.031296692 | 27.77670876 | 1.35E-169 | .. |
| PCOS | 2.86440974 | 0.180928417 | 15.8317294 | 1.97969E-56 | .. |
| weightgainMeds | 0.918797892 | 0.029399124 | 31.25255976 | 4.496E-214 | .. |
| smoke_statusFormer | 1.583039992 | 0.033535405 | 47.20503511 | 0 | .. |
| smoke_statusNever | 0.991923547 | 0.031330222 | 31.66027848 | 1.2451E-219 | .. |
| t2dm | 3.570432777 | 0.029723206 | 120.1227353 | 0 | .. |
| cushing | 2.109278103 | 0.391019905 | 5.394298542 | 6.88426E-08 | .. |
| harmonized PRS 9:sleep | -0.018859973 | 0.020190444 | -0.934103911 | 0.350251033 | 0.389167814 |

**ST43:** Linear regression. BMI ∼ age + sex + pc’s 1-10 + clinical covariates + Hypothalamic dysregulation pPS * sleep.

| term | Beta | SE | t-statistic | p-value | q-value (BH-FDR) |
| --- | --- | --- | --- | --- | --- |
| (Intercept) | 26.5766975 | 0.076636294 | 346.7899629 | 0 | .. |
| age | -0.00867429 | 0.001047191 | -8.283386112 | 1.20197E-16 | .. |
| sex | 0.642126861 | 0.016668712 | 38.52288399 | 0 | .. |
| harmonized PRS 10 | 0.313722128 | 0.019876901 | 15.7832512 | 4.27025E-56 | .. |
| pc1 | 0.002624848 | 0.000164544 | 15.95223756 | 2.89931E-57 | .. |
| pc2 | 0.010913299 | 0.000313337 | 34.82929953 | 2.916E-265 | .. |
| pc3 | 0.00065849 | 0.000580878 | 1.13361109 | 0.256958608 | .. |
| pc4 | 0.002415245 | 0.000780522 | 3.094395309 | 0.001972323 | .. |
| pc5 | 0.013852472 | 0.001060537 | 13.06174634 | 5.5791E-39 | .. |
| pc6 | 0.009176113 | 0.001619286 | 5.666763555 | 1.4565E-08 | .. |
| pc7 | 0.004528247 | 0.001634711 | 2.770060424 | 0.005604928 | .. |
| pc8 | 0.006918928 | 0.001683142 | 4.110722009 | 3.94527E-05 | .. |
| pc9 | -0.014255046 | 0.001813085 | -7.862316704 | 3.78311E-15 | .. |
| pc10 | 0.007197456 | 0.002028599 | 3.547992658 | 0.000388237 | .. |
| physical activity | -0.695257676 | 0.017454135 | -39.83340778 | 0 | .. |
| sedentary | -1.639727168 | 0.025845258 | -63.44402437 | 0 | .. |
| diet | -0.495178141 | 0.017171259 | -28.83761391 | 1.276E-182 | .. |
| sleep | -0.481993899 | 0.024045721 | -20.04489269 | 2.55371E-89 | .. |
| higher ed | -0.728473299 | 0.016829422 | -43.28569878 | 0 | .. |
| TDI group | 0.35410698 | 0.013165354 | 26.89688215 | 3.6666E-159 | .. |
| depression | 0.660703076 | 0.027614893 | 23.92560711 | 2.1695E-126 | .. |
| hypothyroidism | 0.872461005 | 0.031305004 | 27.86969824 | 1.0188E-170 | .. |
| PCOS | 2.87007294 | 0.180978786 | 15.85861527 | 1.29123E-56 | .. |
| weightgainMeds | 0.925025495 | 0.029407063 | 31.45589572 | 7.764E-217 | .. |
| smoke_statusFormer | 1.580043911 | 0.03354473 | 47.10259677 | 0 | .. |
| smoke_statusNever | 0.993588147 | 0.031339402 | 31.70411907 | 3.1144E-220 | .. |
| t2dm | 3.554317642 | 0.029731666 | 119.5465342 | 0 | .. |
| cushing | 2.17025017 | 0.391128828 | 5.548683738 | 2.88066E-08 | .. |
| harmonized PRS 10:sleep | -0.048673097 | 0.021326654 | -2.282265953 | 0.022474327 | 0.044948654 |

**ST44:** Linear regression. BMI ∼ age + sex + pc’s 1-10 + clinical covariates + Metabolically healthy pPS * sleep.

| term | Beta | SE | t-statistic | p-value | q-value (BH-FDR) |
| --- | --- | --- | --- | --- | --- |
| (Intercept) | 26.57772797 | 0.076704438 | 346.4953075 | 0 | .. |
| age | -0.008856191 | 0.00104813 | -8.44951345 | 2.93775E-17 | .. |
| sex | 0.639181599 | 0.016684257 | 38.31046169 | 0 | .. |
| harmonized PRS 11 | 0.34310173 | 0.02556205 | 13.42230869 | 4.59719E-41 | .. |
| pc1 | 0.002609463 | 0.000164693 | 15.84444681 | 1.61751E-56 | .. |
| pc2 | 0.010929715 | 0.000313619 | 34.85029639 | 1.4063E-265 | .. |
| pc3 | 0.000561544 | 0.000581403 | 0.965841535 | 0.334124245 | .. |
| pc4 | 0.002440754 | 0.000781224 | 3.12427017 | 0.001782634 | .. |
| pc5 | 0.013829014 | 0.001061491 | 13.02791468 | 8.69434E-39 | .. |
| pc6 | 0.00902058 | 0.001620746 | 5.565695057 | 2.61328E-08 | .. |
| pc7 | 0.004508635 | 0.001636182 | 2.755583819 | 0.005859096 | .. |
| pc8 | 0.006873519 | 0.001684657 | 4.080069366 | 4.50337E-05 | .. |
| pc9 | -0.014139492 | 0.001814707 | -7.79160952 | 6.63672E-15 | .. |
| pc10 | 0.007057902 | 0.002030421 | 3.476077805 | 0.000508875 | .. |
| physical activity | -0.696443449 | 0.017469776 | -39.86561969 | 0 | .. |
| sedentary | -1.638725102 | 0.025868492 | -63.3483035 | 0 | .. |
| diet | -0.486536085 | 0.017184713 | -28.31214527 | 4.1443E-176 | .. |
| sleep | -0.476302743 | 0.024066378 | -19.79120972 | 4.02029E-87 | .. |
| higher ed | -0.722239709 | 0.016843999 | -42.87816084 | 0 | .. |
| TDI group | 0.354280569 | 0.013177221 | 26.88583313 | 4.9337E-159 | .. |
| depression | 0.654627267 | 0.02763969 | 23.68432019 | 6.7707E-124 | .. |
| hypothyroidism | 0.875661303 | 0.031333178 | 27.94677549 | 1.1887E-171 | .. |
| PCOS | 2.877052185 | 0.18114211 | 15.88284569 | 8.77958E-57 | .. |
| weightgainMeds | 0.928177507 | 0.029433826 | 31.53438172 | 6.5908E-218 | .. |
| smoke_statusFormer | 1.582850257 | 0.033574884 | 47.14387918 | 0 | .. |
| smoke_statusNever | 0.989895235 | 0.031367053 | 31.55843892 | 3.0909E-218 | .. |
| t2dm | 3.594620032 | 0.029778957 | 120.7100698 | 0 | .. |
| cushing | 2.134458911 | 0.391479872 | 5.452282648 | 4.97657E-08 | .. |
| harmonized PRS 11:sleep | -0.082411413 | 0.027409183 | -3.006708093 | 0.002641147 | 0.013205734 |

